# Understanding the genetic complexity of puberty timing across the allele frequency spectrum

**DOI:** 10.1101/2023.06.14.23291322

**Authors:** Katherine A Kentistou, Lena R Kaisinger, Stasa Stankovic, Marc Vaudel, Edson M de Oliveira, Andrea Messina, Robin G Walters, Xiaoxi Liu, Alexander S Busch, Hannes Helgason, Deborah J Thompson, Federico Santon, Konstantin M Petricek, Yassine Zouaghi, Isabel Huang-Doran, Daniel F Gudbjartsson, Eirik Bratland, Kuang Lin, Eugene J Gardner, Yajie Zhao, Raina Jia, Chikashi Terao, Margie Riggan, Manjeet K Bolla, Mojgan Yazdanpanah, Nahid Yazdanpanah, Jonath P Bradfield, Linda Broer, Archie Campbell, Daniel I Chasman, Diana L Cousminer, Nora Franceschini, Lude H Franke, Giorgia Girotto, Chunyan He, Marjo-Riitta Järvelin, Peter K Joshi, Yoichiro Kamatani, Robert Karlsson, Jian’an Luan, Kathryn L Lunetta, Reedik Mägi, Massimo Mangino, Sarah E Medland, Christa Meisinger, Raymond Noordam, Teresa Nutile, Maria Pina Concas, Ozren Polašek, Eleonora Porcu, Susan M Ring, Cinzia Sala, Albert V Smith, Toshiko Tanaka, Peter J van der Most, Veronique Vitart, Carol A Wang, Gonneke Willemsen, Marek Zygmunt, Thomas U Ahearn, Irene L Andrulis, Hoda Anton-Culver, Antonis C Antoniou, Paul L Auer, Catriona LK Barnes, Matthias W Beckmann, Amy Berrington, Natalia V Bogdanova, Stig E Bojesen, Hermann Brenner, Julie E Buring, Federico Canzian, Jenny Chang-Claude, Fergus J Couch, Angela Cox, Laura Crisponi, Kamila Czene, Mary B Daly, Ellen W Demerath, Joe Dennis, Peter Devilee, Immaculata De Vivo, Thilo Dörk, Alison M Dunning, Miriam Dwek, Johan G Eriksson, Peter A Fasching, Lindsay Fernandez-Rhodes, Liana Ferreli, Olivia Fletcher, Manuela Gago-Dominguez, Montserrat García-Closas, José A García-Sáenz, Anna González-Neira, Harald Grallert, Pascal Guénel, Christopher A Haiman, Per Hall, Ute Hamann, Hakon Hakonarson, Roger J Hart, Martha Hickey, Maartje J Hooning, Reiner Hoppe, John L Hopper, Jouke-Jan Hottenga, Frank B Hu, Hanna Hübner, David J Hunter, ABCTB Investigators, Helena Jernström, Esther M John, David Karasik, Elza K Khusnutdinova, Vessela N Kristensen, James V Lacey, Diether Lambrechts, Lenore J Launer, Penelope A Lind, Annika Lindblom, Patrik KE Magnusson, Arto Mannermaa, Mark I McCarthy, Thomas Meitinger, Cristina Menni, Kyriaki Michailidou, Iona Y Millwood, Roger L Milne, Grant W Montgomery, Heli Nevanlinna, Ilja M Nolte, Dale R Nyholt, Nadia Obi, Katie M O’Brien, Kenneth Offit, Albertine J Oldehinkel, Sisse R Ostrowski, Aarno Palotie, Ole B Pedersen, Annette Peters, Giulia Pianigiani, Dijana Plaseska-Karanfilska, Anneli Pouta, Alfred Pozarickij, Paolo Radice, Gad Rennert, Frits R Rosendaal, Daniela Ruggiero, Emmanouil Saloustros, Dale P Sandler, Sabine Schipf, Carsten O Schmidt, Marjanka K Schmidt, Kerrin Small, Beatrice Spedicati, Meir Stampfer, Jennifer Stone, Rulla M Tamimi, Lauren R Teras, Emmi Tikkanen, Constance Turman, Celine M Vachon, Qin Wang, Robert Winqvist, Alicja Wolk, Babette S Zemel, Wei Zheng, Ko W van Dijk, Behrooz Z Alizadeh, Stefania Bandinelli, Eric Boerwinkle, Dorret I Boomsma, Marina Ciullo, Georgia Chenevix-Trench, Francesco Cucca, Tõnu Esko, Christian Gieger, Struan FA Grant, Vilmundur Gudnason, Caroline Hayward, Ivana Kolčić, Peter Kraft, Deborah A Lawlor, Nicholas G Martin, Ellen A Nøhr, Nancy L Pedersen, Craig E Pennell, Paul M Ridker, Antonietta Robino, Harold Snieder, Ulla Sovio, Tim D Spector, Doris Stöckl, Cathie Sudlow, Nic J Timpson, Daniela Toniolo, André Uitterlinden, Sheila Ulivi, Henry Völzke, Nicholas J Wareham, Elisabeth Widen, James F Wilson, The Lifelines Cohort Study, The Danish Blood Donor study, The Ovarian Cancer Association Consortium, The Breast Cancer Association Consortium, The Biobank Japan Project, The China Kadoorie Biobank Collaborative Group, Paul DP Pharoah, Liming Li, Douglas F Easton, Pål Njølstad, Patrick Sulem, Joanne M Murabito, Anna Murray, Despoina Manousaki, Anders Juul, Christian Erikstrup, Kari Stefansson, Momoko Horikoshi, Zhengming Chen, I Sadaf Farooqi, Nelly Pitteloud, Stefan Johansson, Felix R Day, John RB Perry, Ken K Ong

## Abstract

Pubertal timing varies considerably and has been associated with a range of health outcomes in later life. To elucidate the underlying biological mechanisms, we performed multi-ancestry genetic analyses in ∼800,000 women, identifying 1,080 independent signals associated with age at menarche. Collectively these loci explained 11% of the trait variance in an independent sample, with women at the top and bottom 1% of polygenic risk exhibiting a ∼11 and ∼14-fold higher risk of delayed and precocious pubertal development, respectively. These common variant analyses were supported by exome sequence analysis of ∼220,000 women, identifying several genes, including rare loss of function variants in *ZNF483* which abolished the impact of polygenic risk. Next, we implicated 660 genes in pubertal development using a combination of *in silico* variant-to-gene mapping approaches and integration with dynamic gene expression data from mouse embryonic GnRH neurons. This included an uncharacterized G-protein coupled receptor *GPR83*, which we demonstrate amplifies signaling of *MC3R*, a key sensor of nutritional status. Finally, we identified several genes, including ovary-expressed genes involved in DNA damage response that co-localize with signals associated with menopause timing, leading us to hypothesize that the ovarian reserve might signal centrally to trigger puberty. Collectively these findings extend our understanding of the biological complexity of puberty timing and highlight body size dependent and independent mechanisms that potentially link reproductive timing to later life disease.

## Introduction

Age at menarche (AAM), the onset of menses in females, represents the start of reproductive maturity and is a widely reported marker of pubertal timing. Menarche normally occurs between ages 10 to 15 years^1^, and its variation is associated with risks of several health outcomes, including obesity, type 2 diabetes (T2D), cardiovascular disease, and hormone-sensitive cancers^2,3,4,5,6^. Thus, widespread secular trends towards earlier puberty timing may have an important impact on public health^7^. AAM is a highly polygenic trait^8^ and previous genome-wide association studies (GWAS) have identified ∼400 common genetic loci^9,10,11,12,13^, the vast majority of which were discovered in samples of European ancestry. AAM has a strong genetic correlation with male puberty timing (*R*_*g*_=0.68)^14^, as well as with adiposity (BMI, *R*_*g*_=-0.35)^9^ and specific pathways have been identified that link nutrient sensing to reproductive hormone axis activation. For example, we recently reported that *MC3R* is the key hypothalamic sensor linking nutritional status to puberty timing^15^.

Previously reported GWAS signals in ∼370,000 women of European ancestry explained ∼7.4% of the population variance in AAM, corresponding to ∼25% of the estimated heritability^9^. Here, through an expanded GWAS in up to 799,845 women, including 166,890 of East Asian ancestry, we identify 1080 independent signals for AAM. Female participants who carry an excess of these alleles have equivalent risk of precocious or delayed puberty compared to those carrying clinically relevant monogenic alleles. We complement these common variant analyses by undertaking the first large-scale assessment of rare variation in puberty timing in 222,283 women with exome sequence data. Through subsequent variant to gene mapping approaches, we implicate 660 genes, which collectively shed further light on the biological determinants of puberty timing and the mechanisms linking it to disease risks.

## Results

We performed a GWAS meta-analysis for AAM, in up to 799,845 women, by combining data from five strata: i) 38 ReproGen consortium cohorts (N=180,269), ii) UK Biobank (N=238,040), iii) the Breast Cancer Association Consortium and the Ovarian Cancer Association Consortium (N=137,815), iv) 23andMe, Inc. (N=76,831), and v) three East Asian biobanks: the China Kadoorie Biobank, the Biobank Japan, and the Korean Genome and Epidemiology Study (N=166,890). All studies provided GWAS data imputed to at least 1000 Genomes reference panel density (Supplementary Table 1), yielding a total of ∼12.7 million genetic variants in the final meta-analysis. We did not find evidence of test statistic inflation due to population structure (LDSC intercept=1.07, SE=0.03).

To maximise the discovery of genomic signals for AAM, we used a combination of distance-based clumping and approximate conditional analysis (see Methods) in the European-strata meta-analysis and in the all-ancestry meta-analysis, to identify signals that are homogenous across the two ancestry groups. European-strata identified signals (N=935) were supplemented with additional signals from the all-ancestry analysis (N=145), resulting in a total of 1080 statistically independent signals for AAM at a genome-wide significance (*P*<5×10^−8^, Figure 1, Supplementary Table 2). Effect sizes ranged from 3.5 months/allele for rarer alleles (MAF=0.9%) to ∼5 days/allele for more common variants (Supplementary Figure 1). Across the 145 additional signals, we observed a median 1.16-fold increase in χ^2^ for their association with AAM in the all-ancestry analysis compared to European-only, which is proportionate to the added number of East-Asian samples (∼21% of the total).

**Figure 1.**
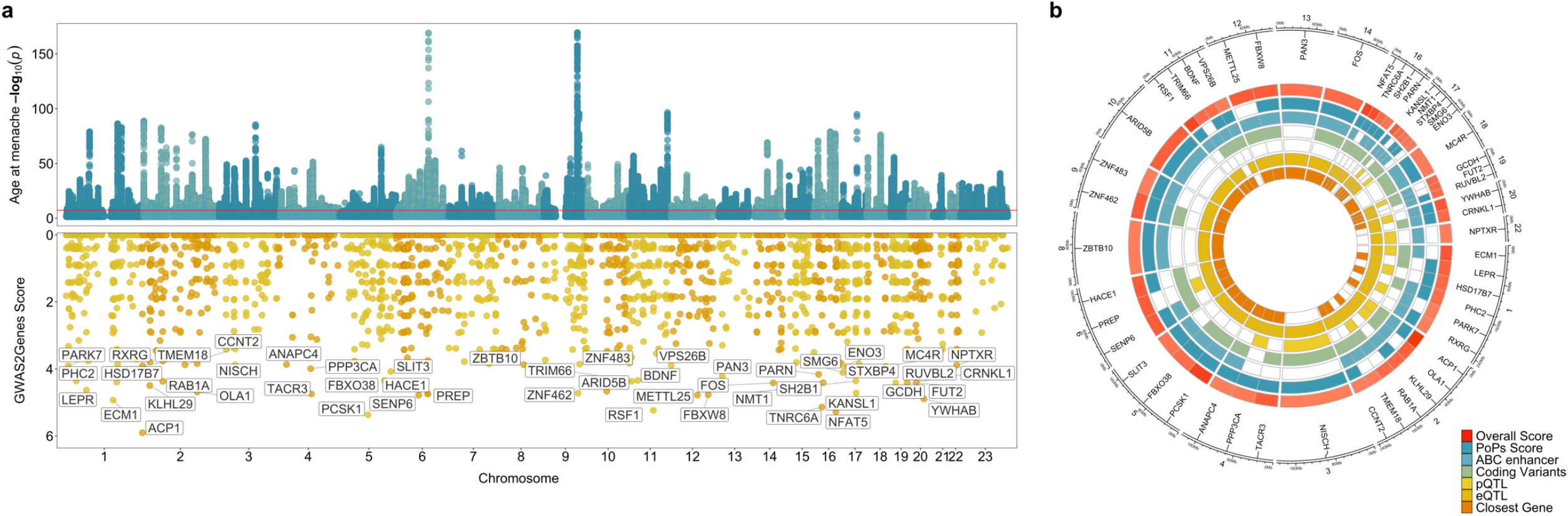
Age at menarche GWAS and gene prioritisation. (a) Miami plot showing signals from the European meta-analysis for age at menarche (upper panel) and genome-wide G2G scores with names of the top 50 genes annotated (lower panel). The upper panel Y-axis is capped at −log_10_(1×10^−150^) for visibility. (b) The 50 top scoring genes implicated by G2G, annotated by their sources of evidence. Relevant data are included in Supplementary Tables 2 and 13-16.

Independent replication data from the Danish Blood Donors study (N=35,467) (Supplementary Table 3) was available for 969/1080 signals^16^. Of these, 862 showed directionally concordant associations (89%, *P*_Binomial_=2.9×10^−147^). In this independent sample, the variance explained in AAM doubled from 5.6% for 355 available previously reported signals^9^ to 11% for the 969 signals with available data. We also sought indirect confirmation of AAM signals by association with age at voice breaking (AVB) in men from the UK Biobank study (N=191,235) and 23andMe (N=55,871) (Supplementary Table 4)^17,18,14^. Of the 1080 AAM signals, 909/1080 (84%, *P*_Binomial_=2.6×10^−122^) showed directionally concordant associations with AVB in UK Biobank (including 354 at *P*<0.05). Similarly, 852/1067 (79%, *P*Binomial=1.8×10^−90^) AAM signals available in 23andMe showed directionally concordant associations with AVB (217 at *P*<0.05).

### Exome sequence analyses identify novel rare variants of large effect

Previous genetic studies for AAM have largely been restricted to assessing the role of common, largely non-coding, genetic variation. We sought to address this, by performing an exome-wide association study (ExWAS) in 222,283 European ancestry women in UK Biobank. Gene burden tests were performed by collapsing rare variants (MAF<0.1%) in each gene according to two overlapping predicted functional categories: i) high-confidence protein truncating variants (PTVs) and ii) PTVs plus missense variants with CADD score^19^ ≥25 (termed ‘damaging variants’, DMG). Six genes were associated with AAM at exome-wide significance (*P*<1.54×10^−6^, 0.05/32,434 tests, Figure 2, Supplementary Figure 2, Supplementary Figure 3, Supplementary Figure 4, Supplementary Table 5). This included two genes previously reported in rare monogenic disorders of puberty: *TACR3* (beta=0.62 years, *P*=3.2×10^−19^, N=489 DMG carriers) previously implicated in normosmic idiopathic hypogonadotropic hypogonadism (IHH)^20^, and *MKRN3* (beta=-0.59 years, *P*=1.4×10^−7^, N=187 DMG carriers) previously implicated in familial central precocious puberty (CPP)^21^. Furthermore, *MC3R* (beta=0.33 years, *P*=1.6×10^−9^, N=796 DMG carriers) was recently reported to link nutrient sensing to key hypothalamic neurons^15^.

**Figure 2.**
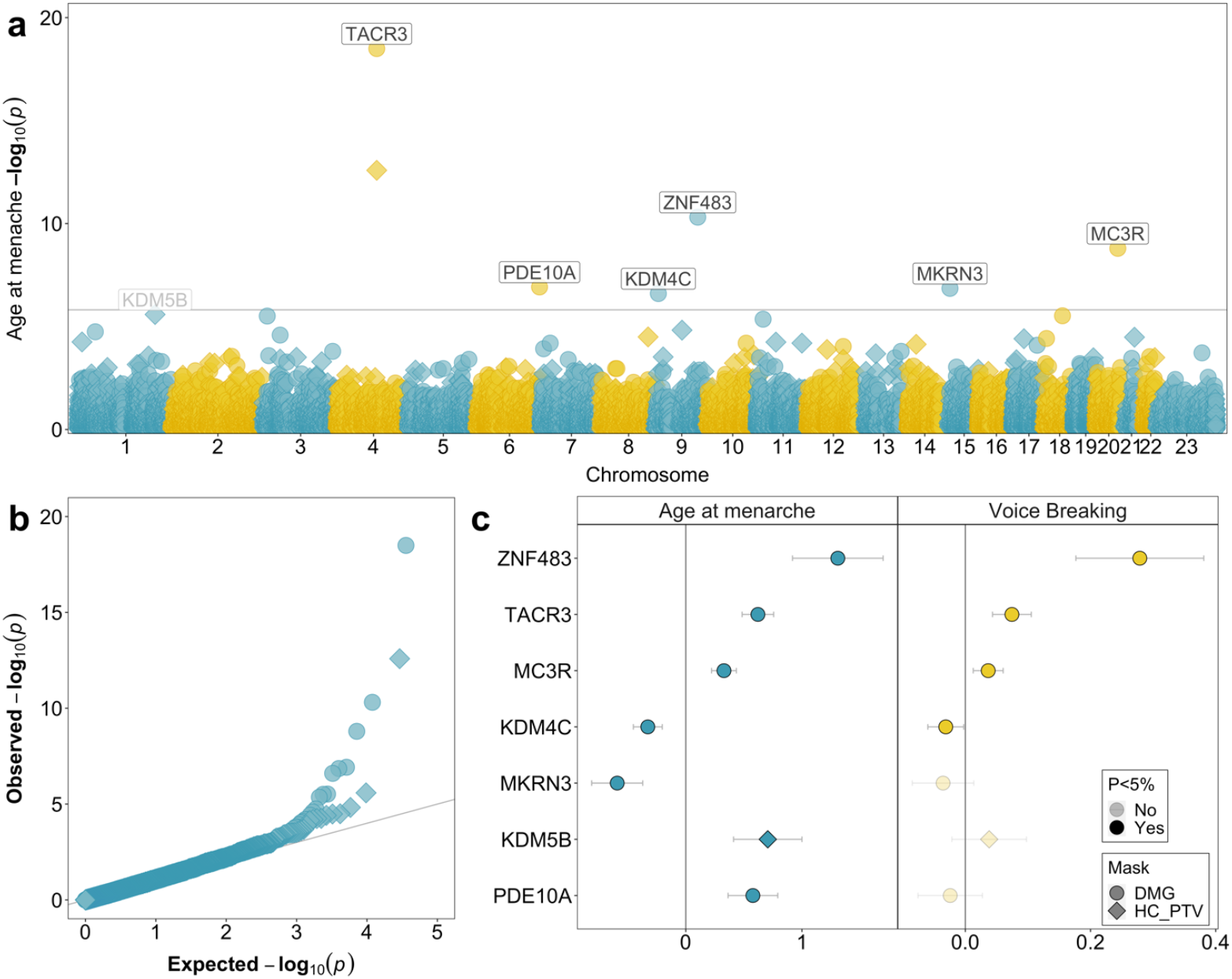
Exome-wide rare (MAF <0.1%) variant associations with age at menarche. (a) Manhattan plot showing gene burden test results for age at menarche. Genes passing exome-wide significance (*P*<1.54×10^−6^) are highlighted; in addition, *KDM5B* shows a sub-threshold association (*P*=2.6×10^−6^). Point shapes indicate variant predicted functional class (DMG, damaging; HC PTV, high confidence protein truncating). (b) QQ plot for gene burden tests. (c) Comparison of gene burden associations for age at menarche (female participants, years) and age at voice breaking (men, 3 categories). Relevant data are included in Supplementary Table 5.

Of the three novel genes, *KDM4C* (beta=-0.33 years, *P*=2.5×10^−7^, N=582 DMG carriers) encodes a lysine-specific histone demethylase likely involved in epigenetically regulating hypothalamic-pituitary-gonadal (HPG) axis genes^22^. A second gene in this small family (*KDM5B*) showed near exome-wide significant association with AAM (*P*=2.6×10^−6^). In addition, *PDE10A* (beta=0.58 years, *P*=1.2×10^−^ 7, N=196 DMG carriers) encodes phosphodiesterase 10A, which regulates the intracellular concentration of cyclic nucleotides and hence signal transduction^23^. Finally, *ZNF483* (beta=1.31 years, *P*=4.9×10^−11^, N=59 DMG carriers), encodes a zinc finger protein transcription factor involved in neuronal differentiation^24^ and self-renewal of pluripotent stem cells^25^.

We were able to confirm four of these seven genes (*KDM4C, MC3R, TACR3* and *ZNF483*) using voice breaking data in 178,625 men with exome sequence data in UK Biobank (*P*<0.05, Figure 2, Supplementary Table 5). Lack of association with AVB at *MKRN3* is consistent with previous reports that rare *MKRN3* mutations have greater clinical impact in girls than boys^21,26^. None of the seven genes showed an association with childhood or adult adiposity (Supplementary Table 6).

In addition, we specifically examined rare variant associations with AAM or VB for *ANOS1, CHD7, FGF8* and *WDR11*, which are clinically tested in hypogonadotropic hypogonadism (‘high evidence genes’ on the Genomics England IHH panel^27^) and show a dominant or X-linked mode of inheritance (Supplementary Table 7). Normal puberty timing (AAM: 10-15 years^1^ or VB: “about average”) was reported by all carriers of PTVs in *ANOS1* (N=5 male) and *CHD7* (N=5 female, N=1 male). PTVs in *WDR11* showed no association with delayed puberty with only 7/81 female and 5/68 male carriers reporting delayed puberty. Female carriers of PTVs in *FGF8* showed some evidence of later puberty (beta=1.4 years, *P*=3.6×10^−3^, N=5/10 reported delayed puberty) but with no effect in males (N=1/8 reported delayed puberty) (Supplementary Figure 5). These observations highlight the lower penetrance of rare deleterious variants in large population-based studies compared to in patient cohorts^28, 29,30^.

### Common genetic variants influence risk of phenotypic extremes

Rare pathogenic variants such as the above are described to cause disorders of puberty. However, it remains unclear whether common genetic variants also contribute to abnormal puberty timing. To assess this, we generated a polygenic score (PGS) of AAM in a penalised regression framework using lassosum^31^ and data from our meta-analysis of European ancestry cohorts but excluding UK Biobank. This PGS explained ∼12% of the phenotypic variance in UK Biobank. The PGS was informative in individuals experiencing menarche as early as 8 years old and later than 20, well beyond the normal AAM range (10 to 15 years, Supplementary Figure 6, Supplementary Tables 8 and 9).

We next sought to understand how the risks of early (<10 years) and delayed (>15 years) AAM were influenced by the PGS. Women in the lowest 1% PGS centile reported AAM at mean (SE) 11.49 (0.03) years, compared to 14.46 (0.04) years in the top 1% PGS centile (Supplementary Figure 6, Supplementary Table 10). Compared to women in the 50^th^ PGS centile, those in the top 1% PGS were 10.7 times more likely to report late AAM (OR [8.20-13.96], *P*=2.6×10^−68^), while women in the lowest 1% PGS were 14.2 times more likely to report early AAM (OR [7.13-28.39], *P*=5.1×10^−14^). Collectively these findings suggest that common genetic variants contribute to the risk of rare clinical disorders of extremely early (precocious) and delayed puberty.

To evaluate the predictive performance of our AAM signals, we compared these to phenotypic predictors in 3,140 female children from the ALSPAC study. The AAM signals in combination explained more variance in AAM than childhood BMI, parental BMI or mother’s AAM (Supplementary Table 11). Furthermore, they had a similar ability to predict extremes of AAM (beyond 2 SD) than a multi-phenotype predictor (Supplementary Figure 7), and a combined genotype and phenotype model showed high predictive ability for early AAM (AUROC = 0.75 [95% CI 0.68-0.82]) and late AAM (AUROC = 0.85 [95% CI 0.81-0.92]).

We next tested whether carrying rare variants in the AAM ExWAS genes modifies the common polygenic influence on AAM. We saw that the effect of the common variant PGS on AAM was attenuated in the 49 unrelated carriers of DMG variants in *ZNF483* (beta_non-carriers_=0.564 years/SD, SE=0.003; beta_carriers_=0.084, SE=0.214; *P*_interaction_=0.025, Figure 3, Supplementary Table 12). To confirm that this was not a reflection of reduced power due to the low number of carriers, we estimated the expected relationship for non-carriers in 10,000 random subsamples of 49 participants and found that the observed carrier effect was unlikely by chance (*P*=0.015, Supplementary Figure 8).

**Figure 3.**
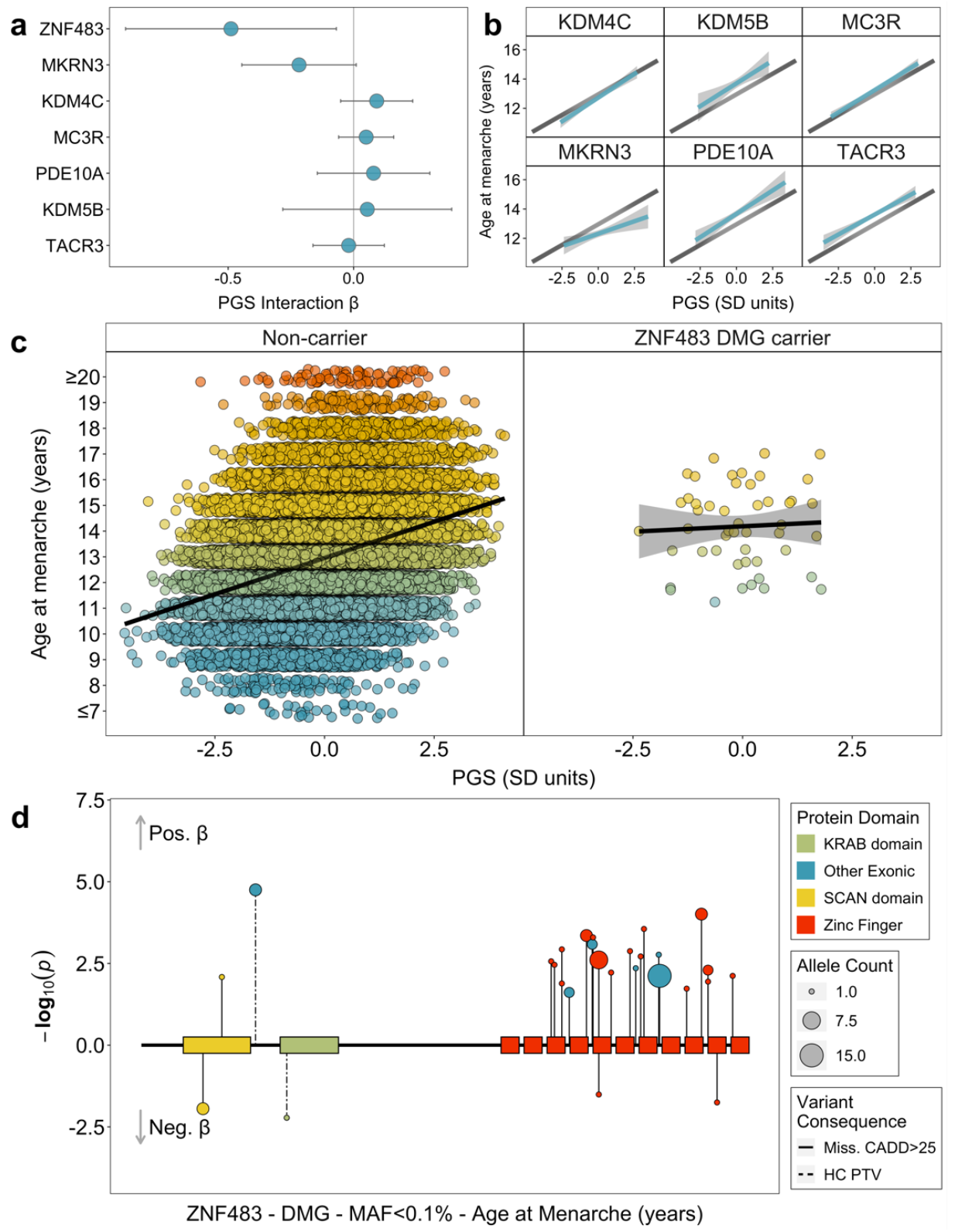
Epistatic interactions between rare coding variants and common genetic susceptibility on age at menarche in UK Biobank. (**a**) Interaction effects (95% CI) on age at menarche between a GWAS polygenic score (PGS) and carriage of qualifying rare variants in seven exome-highlighted genes. Predicted mean (95% CI) age at menarche in (**b**) non-carriers (black) and carriers (light blue) of rare variants in six genes without significant interaction effects and (**c**) in non-carriers (left panel) and carriers (right) of rare variants in *ZNF483* which shows significant interaction. In (c) points show individual age at menarche values. (**d**) Plot of individual rare damaging (DMG) variant associations with age at menarche by *ZNF483* functional domains. The coding part of *ZNF483* is depicted by the horizontal black line. Included damaging variants had a minor allele frequency (MAF) <0.1% and were annotated to either be high-confidence protein truncating variants or missense variants with CADD score >=25. Relevant data are included in Supplementary Table 12.

Using ENCODE ChIP-seq data^32^, we found that the transcriptional targets of *ZNF483* are enriched for in the AAM GWAS (fGWAS^32^; *P*=2.6×10^−7^), and that greater *ZNF483* binding confers earlier AAM (SLDP^33^; Z=-4.9, *P*=4.8×10^−7^), which is directionally-concordant with our observed effect of rare DMG variants on later AAM. This was further corroborated by functional-domain-specific gene burden analyses, which showed a larger effect on AAM of *ZNF483* DMG variants located within zinc finger domains (beta=1.615 years, SE=0.293, *P*=3.59×10^−8^), rather than DMG variants outwith these domains (beta=0.816 years, SE=0.298, *P*=6.2×10^−3^, Figure 3). This data suggests that rare DMG variants in *ZNF483* confer later AAM by disrupting the protein’s ability to bind to its multiple DNA targets.

### Implicating AAM genes through variant to gene mapping approaches

To implicate putatively causal genes that underlie our 1080 common variant signals for AAM, we developed the framework ‘GWAS to Genes’ (G2G) that integrates genomic and functional data across six sources (Methods, Figure 1, Supplementary Tables 13-16). We identified proximal genes (within 500kb up or downstream) to the 1080 AAM signals and scored genes based on the degree of evidence linking our lead index variants to the function of these genes. To achieve this, we implicated genes by identifying signals that co-localised with a) known enhancers and regulatory elements^34^, b) non-synonymous variants, c) expression quantitative trait loci (eQTL) specifically in tissues enriched for AAM associations (Supplementary Figure 9, Supplementary Table 17), and d) circulating protein QTL (pQTL) from whole blood (see Methods). In addition, we integrated gene-level associations for aggregated non-synonymous common variants using MAGMA^35^ and gene scores from PoPs^36^, which uses bulk human and mouse data with information on scRNA, gene pathways and protein interactions to link genes to GWAS signals. Individual genes were further upweighted if they were the nearest gene to the signal^37,38^.

Using this approach, our 1080 signals were found to be proximal to 10,341 genes, of which 660 ‘high-confidence AAM genes’ were identified as the highest-scoring gene at a locus and with at least two lines of evidence (Supplementary Figure 10 & Supplementary Table 18; top-scoring genes at each of the 1080 loci are also listed in Supplementary Table 19). High-confidence AAM genes include established components of the HPG axis that are disrupted in rare monogenic disorders of puberty (*CADM1, CHD4, CHD7, FEZF1, GNRH1, KISS1, SPRY4, TAC3, TACR3, TYRO3*)^39^, and other recently reported candidate genes (*PLEKHA5, TBX3, ZNF462*)^40,41^. Other AAM genes have recognised roles in sex hormone secretion and gametogenesis (*ACVR2A, CYP19A1, HSD17B7, INHBA, INHBB, MC3R, PCSK2*)^42^, are disrupted in rare monogenic disorders of multiple pituitary hormone deficiency (*OTX2, SOX2, SOX3, SST*)^43^, monogenic obesity (*BDNF, LEPR, MC4R, NTRK2, PCSK1, SH2B1*)^44^or syndromes characterised by hypogonadism (Noonan Syndrome: *BRAF, SOS1*; Bardet-Biedl Syndrome: *BBS4*; Prader-Willi/Angelman Syndrome: *NDN, SNRPN, UBE3A*)^45,46,47,48^. Other mechanisms implicated by high-confidence AAM genes include: insulin and insulin-like growth factor (IGF) signalling (*CALCR, GHR, IGF1R, INSR, NEUROD1, NSMCE2, PAPPA2, SOCS2*)^49^; thyroid hormone signalling (*THRB*)^50^; and the Polycomb silencing complex (*CBX4, CTBP2, FBRSL1, JARID2, PHC2, SCMH1, TNRC6A*)^51^. We also found strong supportive evidence for all genes identified by the exome variant associations, except *KDM5B* (Supplementary Figure 11).

### Weight gain related and unrelated puberty signals

Phenotypic, genetic and mechanistic links between higher BMI and earlier AAM are well described^15^, but it is challenging to distinguish whether individual AAM signals have a primary effect on puberty or weight status^9^. Here, 83 out of the 1080 AAM signals colocalised (at PP>=0.5) and also showed genome-wide significant association with adult BMI (Supplementary Table 20), and 53 further AAM signals colocalised with adult BMI and showed association with BMI at *P*<4.6×10^−5^ (based on 1080 tests). Of these 136 colocalising signals, at 126 the AAM-reducing allele was associated with higher adult BMI (Supplementary Table 20).

To identify AAM signals with or without a primary effect on early weight gain, we clustered the 1080 AAM signals by their associations with body weight from birth to age 8 years (before the normal age at puberty onset) in the Norwegian MoBa Cohort (N=26,681 children)^52^. We identified three trajectories: 464 AAM signals (44%) formed a ‘moderate early weight gain’ trajectory, and 15 (1%) formed a ‘high early weight gain’ trajectory; both trajectories were characterised by effects of AAM-reducing alleles on higher weight gain across early childhood. The remaining 586 (55%) AAM signals formed a ‘no early weight gain’ trajectory; yet in combination AAM-reducing alleles in this trajectory increased adult BMI (beta=0.487 kg/m^2^/year; *P*=1.6×10^−20^) (Supplementary Tables 21 and 22, Figure 4, Supplementary Figure 12). This data indicates a bidirectional causal relationship between AAM and body size, with greater early weight gain leading to earlier AAM, and also earlier AAM leading to higher adult BMI. This approach provides a clear distinction between AAM signals that have primary effects on puberty timing or early weight gain.

**Figure 4.**
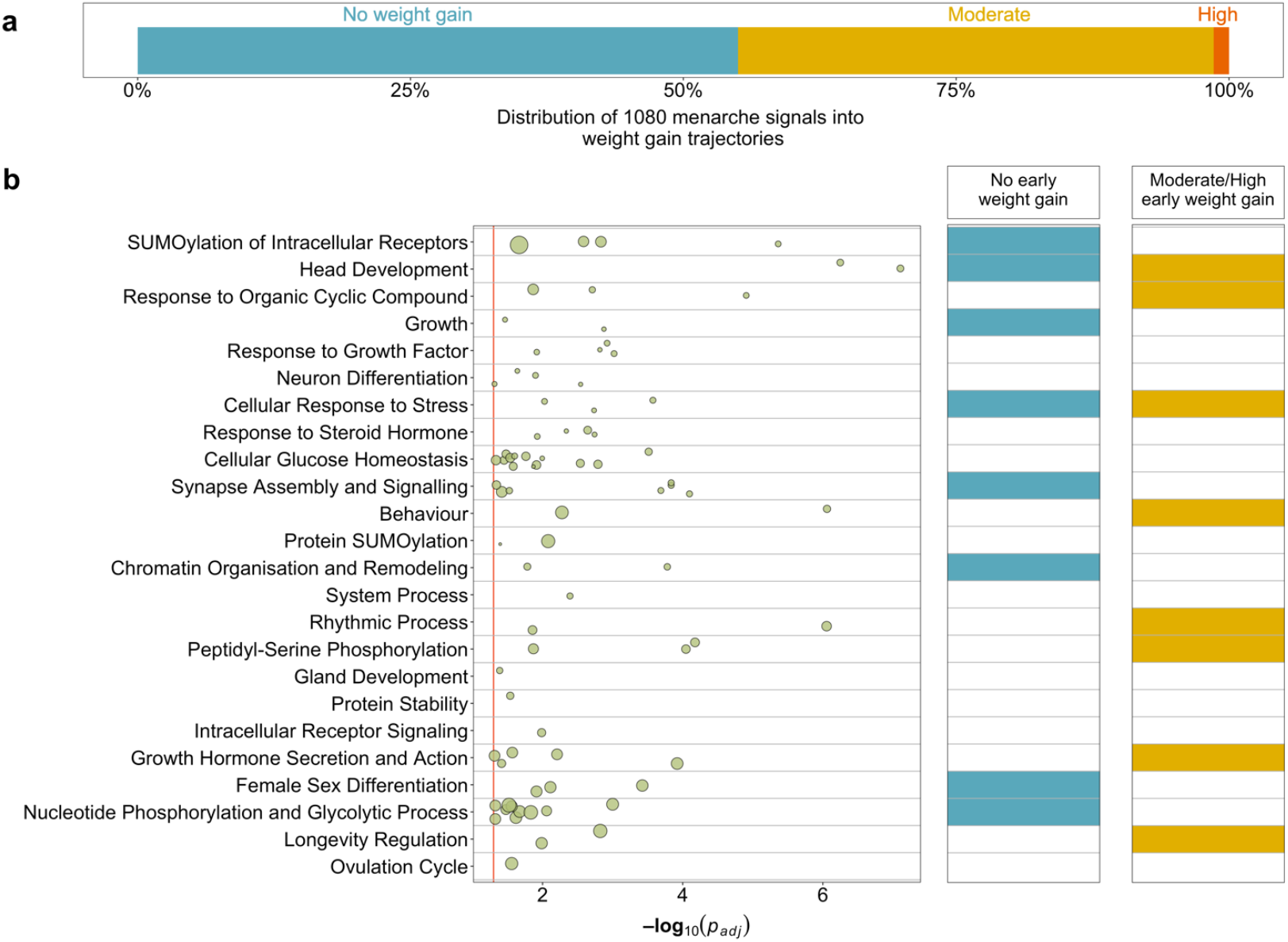
Stratification of age at menarche signals and biological pathway enrichments by their influence on early childhood weight. (a) Proportion of GWAS signals for age at menarche by early childhood weight trajectory. (b) Biological pathways enriched for high confidence age at menarche genes, plus enrichment within early childhood weight trajectories. Row names describe pathway clusters. Strength of associations with individual pathways are indicated by circles. Circle size reflects the proportion of pathway genes that are high confidence age at menarche genes. The right-hand panel indicates whether each pathway cluster remains enriched for age at menarche genes when stratified by early childhood weight trajectory. Extended data are included in Supplementary Tables 21 and 23-26.

### Pathway, tissue and cell-type enrichment of menarche-implicated genes

Genome-wide common variant AAM associations were enriched for genes expressed in several brain regions, and enrichment was highest in the hypothalamus. Outside the brain, we also observed enrichment for genes expressed in the adrenal gland (Supplementary Figure 9 & Supplementary Table 17).

We next performed gene-based pathway analyses on the 660 high-confidence AAM genes using g:Profiler^53^ and identified 83 enriched biological pathways (Supplementary Table 23), which grouped into 24 clusters (Figure 4, Supplementary Figure 13, Supplementary Table 24). These included a number of neuro-endocrine, sexual development, protein and chromatin regulation pathways. To explore distinct biological pathways by early weight trajectories, we repeated the gene-based pathways analysis after stratifying the 660 high-confidence AAM genes into ‘early weight gain’ AAM genes (N=341) or ‘no early weight gain’ AAM genes (N=315) (Supplementary Table 25). Early weight gain AAM genes specifically highlighted hormone regulation, feeding behaviour, rhythmical process, AKT phosphorylation targets and peptidyl-serine modification (Supplementary Figure 14, Supplementary Table 26). Conversely, the no early weight gain AAM genes highlighted female sex differentiation, histone modification, negative regulation of transcription by RNA polymerase II, synapse organisation and DNA repair (Supplementary Figure 15, Supplementary Table 26). Head development and cellular response to stress pathways were enriched among both early weight AAM gene groups (Figure 4, Supplementary Figures 13-15, Supplementary Table 26).

To understand how AAM-associated genes may exert effects on the HPG axis, we explored their expressional dynamics in mouse embryonic GnRH neurons [manuscript in preparation]. RNAseq in GnRH neurons previously identified 2182 genes that showed differential expression between embryonic migration stages (early, intermediate or late, Figure 5), and were categorised into 23 spatio-temporal expression trajectories [manuscript in preparation]. At the genome-wide level, we observed enrichment for GWAS AAM associations among genes that become upregulated in the late (Trajectory01, *P*_adj._=3.8×10^−5^) and mid- to late-stages of GnRH neuron development (Trajectory03, *P*_adj._=0.032, Supplementary Table 27), i.e. when GnRH neurons have completed their migration process and start to make synaptic connections. Of the 660 high-confidence AAM genes, 28 assign to Trajectory01 (*P*_Exact_=2.3×10^−6^), including *NEGR1* and *TNRC6A*, and 31 assign to Trajectory03 (*P*_Exact_=5.2×10^−3^), including *KDM4C, PDE10A* and *TP53BP1*. Both of these GnRH expressional trajectories remained enriched when considering only the subset of non-early weight high-confidence AAM genes (Trajectory01 *P*_Exact_=5.4×10^−4^; Trajectory03 *P*_Exact_=1.4×10^−2^), while Trajectory01 was also enriched when considering only AAM genes that influence early weight gain (Trajectory01 *P*_Exact_=9.3×10^−4^; Trajectory03 *P*_Exact_=0.08).

**Figure 5.**
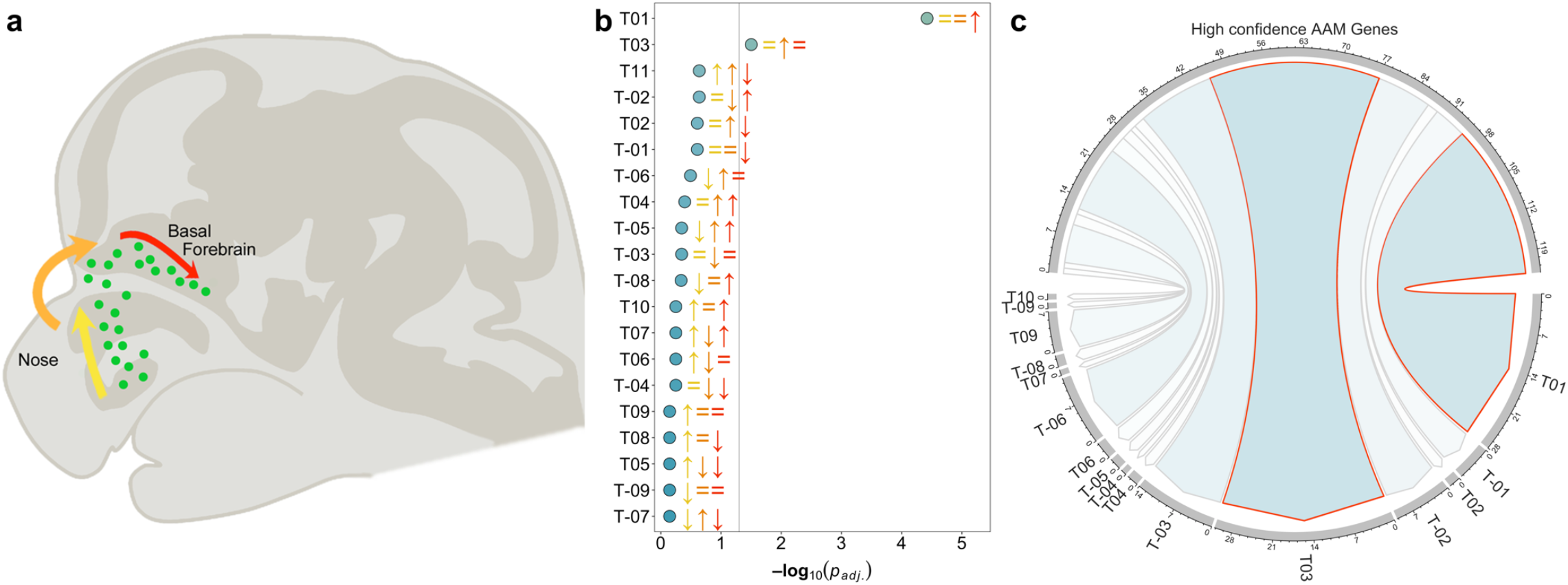
Enrichment of gene drivers of GnRH migration and maturation in the age at menarche GWAS. (a) Schematic representation of the stages of GnRH neuron migration during embryonic development. Using RNAseq data, Pitteloud and colleagues [manuscript in preparation] grouped differentially-expressed genes into 23 expressional trajectories based on their comparative level of expression during the Early (yellow), Intermediate (amber) and Late (red) stages of GnRH migration. (b) Genome-wide MAGMA enrichment for age at menarche associations within each expression trajectory. (c) Trajectories significantly enriched at the genome-wide level in (b) show significant overlap with the 660 high-confidence age at menarche genes. Extended data are included in Supplementary Tables 18 and 27.

### GPCRs and puberty timing

G protein-coupled receptors (GPCRs) regulate several endocrine processes and diseases, including puberty timing^54^ and are therapeutic targets. Here, 24 of the 161 brain-expressed GPCRs (Methods) were implicated in AAM by at least one G2G predictor (Figure 6, Supplementary Table 28). These include *MC3R*, where we recently reported that rare LOF variants, which impair signalling, were associated with delayed puberty^15^, and *GPR83* which encodes a Gα_q11_- and Gαi-coupled GPCR widely expressed in several brain regions^55,56^ and is implicated in energy metabolism^57^. In mice, *Gpr83* and *Mc3r* are reportedly co-expressed in key hypothalamic neurons that control reproduction (KNDy neurons) and growth (GHRH neurons)^15^.

**Figure 6.**
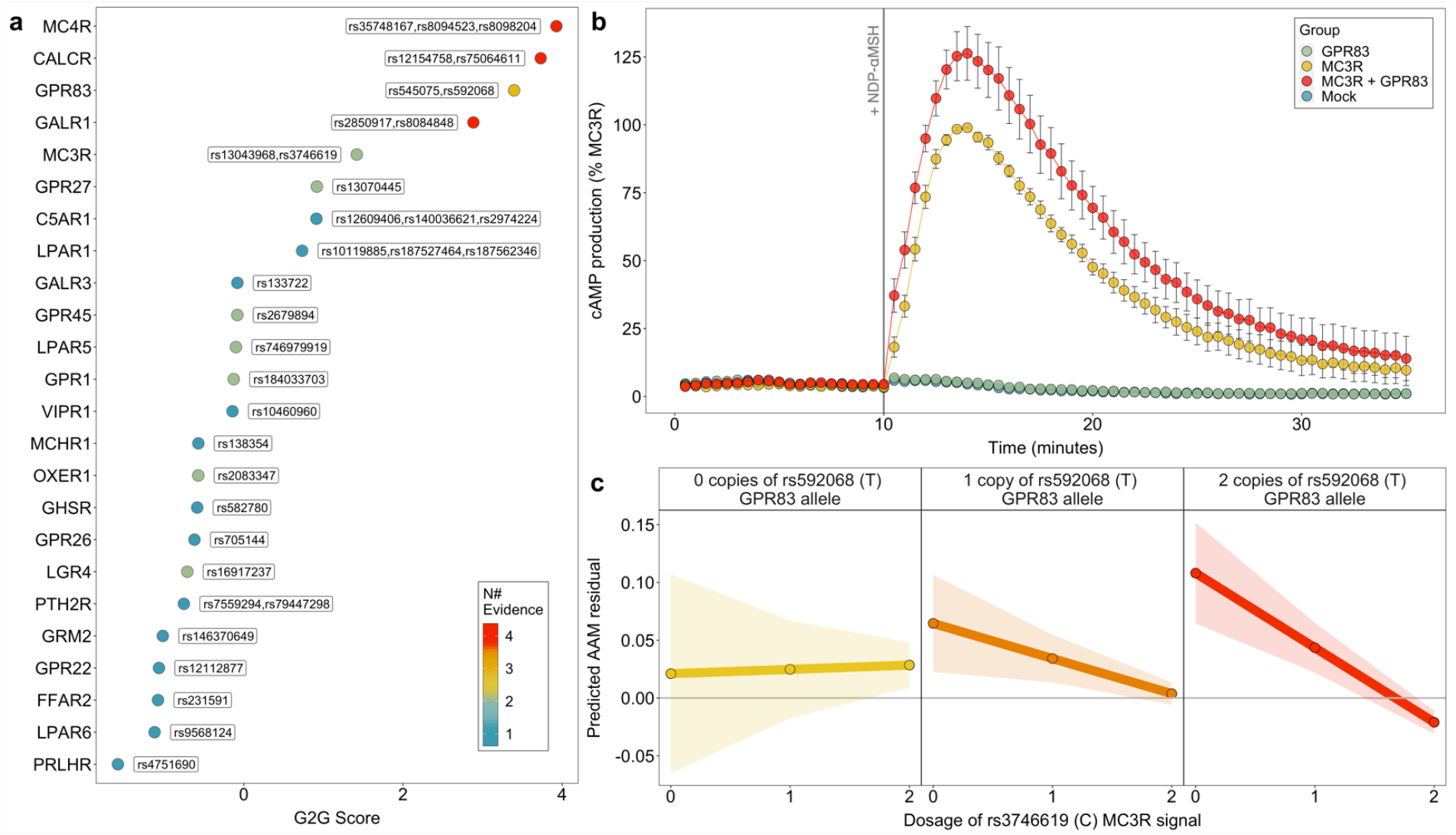
Interactions between G protein-coupled receptors (GPCRs) on age at menarche. (a) 24 brain-expressed GPCRs implicated in age at menarche by G2G analysis of white European GWAS data (b) Time-resolved NDP-αMSH-stimulated cAMP production in HEK293 cells expressing *MC3R*-alone or with both *MC3R* and *GPR83*. Data are mean (standard error) % of the maximal *MC3R*-alone response (from 6 independent experiments). (c) Predicted mean (95% CI) age at menarche according to dosage of *MC3R* function-increasing C alleles at rs3746619 (X-axis in each panel) and *GPR83* expression-increasing T alleles at rs592068 (panels). Beta_interaction_= -0.034 ± 0.015 years, *P*_interaction_=0.02. Extended data are included in Supplementary Tables 28 and 30.

Since dimerisation between GPCRs may affect their signalling^58^, we tested for physical and functional interactions between *MC3R* and *GPR83 in vitro*. Using a Bioluminescence Resonance Energy Transfer (BRET)-based assay in HEK293 cells, we observed a physical and specific interaction between *GPR83* and *MC3R* (Supplementary Figure 16, Supplementary Table 29). We then tested whether *GPR83* modifies canonical *MC3R* signalling, by measuring NDP-α-melanocyte-stimulating hormone (NDP-αMSH)-stimulated cyclic AMP production in HEK293 cells following transfection with plasmids encoding wild-type *GPR83* and *MC3R* separately or together (1:1 ratio). *GPR83* and *MC3R* co-transfection increased cAMP production by 43% compared to *MC3R* alone (*P*=0.03, Figure 6, Supplementary Figure 16, Supplementary Tables 30 and 31).

Consistent with this *in vitro* interaction, we observed statistical genetic epistasis between the common AAM signals at *MC3R* rs3746619; a 5’UTR SNP highly correlated with predicted deleterious coding variants, and *GPR83* rs592068; which colocalises with eQTLs for *GPR83* in brain^59^ and across tissues^60^. Among white European unrelated UK Biobank female participants, *MC3R* function-increasing alleles conferred increasingly earlier AAM in the presence of *GPR83* expression-increasing alleles (beta_interaction_=-0.034 ± 0.015 years, *P*_interaction_=0.02, Figure 6). These findings extend our previous observation that *MC3R* loss of function causes delayed puberty^15^, by indicating that increased *MC3R* function through enhanced *GPR83* expression leads to earlier puberty timing.

### Joint regulation of ages at menarche and menopause

Previous GWASs have estimated a modest shared genetic aetiology between AAM and age at natural menopause (ANM) (genome-wide genetic correlation: r_g_=0.14; *P*=0.003)^61^. ANM gene candidates are mainly expressed in the ovary and implicate DNA damage sensing and repair (DDR) processes that maintain genome stability and hence preserve the ovarian primordial follicle pool^62^.

Of the 1080 AAM signals, nine colocalised (at PP>=0.5) and showed genome-wide significant (*P*<5×10^−8^) association with ANM, and a further 11 AAM signals colocalised and showed association with ANM at *P*<4.6×10^−5^ (=0.05/1080, Supplementary Table 32). We also considered if ANM signals influence AAM. Of the 290 previously reported ANM signals^62^, 21 colocalised and showed association with AAM at *P*<1.7×10^−4^ (=0.05/290), 13 of which were additional to the above AAM signals (Supplementary Table 33). Consistent with the phenotypic association between AAM and ANM^63^, most of the shared common signals (25/33) showed directionally-concordant effects on AAM and ANM (shifting reproductive lifespan earlier or later). Several of these shared signals map to components of the HPG axis, including *GNRH1, INHBB* and *FSHB* (lead SNP rs11031006), which has previously reported associations with related reproductive phenotypes^8,64,65^.

Several other shared AAM and ANM signals map to genes that encode components of DDR processes (*CHD4, CHEK2, DEPTOR, E2F1, MSH6, MSI2, PPARG, RAD18, RAD51, RAD52, SCAI, SPRY4, SUMO1, TP53BP1, TRIP12*, and *WWOX*, see Supplementary Table 34), central to the establishment and maintenance of ovarian oocyte numbers^62^, and not previously implicated in puberty timing. A notable example is rs746979919 (*P*_AAM_=1.5×10^−20^; *P*_ANM_=1.5×10^−34^), which is intronic in *MSH6*, a DNA mismatch repair gene that is primarily expressed in peripheral reproductive tissues, such as ovary and uterus (Supplementary Table 35). Furthermore, the co-localised ANM signal at *CHEK2* captures the previously described frameshift variant 1100delC^66^. This association was further supported by the exome data, with the 347 women carrying rare *CHEK2* protein truncating variants (excluding 1100delC) reporting on average 2 months later AAM (SE=0.99, *P*=0.04). *CHEK2* encodes a cell cycle checkpoint inhibitor that plays a crucial role in culling oocytes with unrepaired DNA damage^62^.

Three of the shared AAM and ANM signals that map to DDR genes were assigned to the ‘moderate early weight gain’ trajectory and further colocalised with adult BMI (*RAD52*: *P*_BMI_=4.8×10^−22^, *TP53BP1*: *P*_BMI_=1.2×10^−5^ and *TRIP12*: *P*_BMI_=2.9×10^−13^), suggesting that some DDR genes might influence AAM via early weight gain (Supplementary Table 20). Other shared AAM and ANM signals that map to DDR genes were assigned to the ‘no early weight gain’ trajectory (*CHD4, MSH6, SCAI* and *SUMO1*) and/or showed no association (*P*>0.05) with adult BMI (*CHEK2, MSI2, PPARG*, and *WWOX*).

## Summary and conclusions

The GWAS signals identified by this expanded multi-ancestry GWAS double the variance explained in AAM compared to previous findings^9^. Furthermore, the common variant PGS contributes substantially to risks of extremely early and late puberty timing. Future studies should explore the potential of this PGS to predict extreme disorders of puberty timing, in contrast to the effects of known monogenic causes.

We describe the first systematic characterisation of common genetic determinants of both ends of reproductive lifespan, AAM and ANM. The 33 identified shared signals highlight the concordant effects of HPG axis genes on both AAM and ANM, and also the influence of ovary-expressed genes involved in DNA damage response. DDR processes have been well described to regulate ovarian oocyte numbers throughout life^62^, but have not previously been implicated in puberty timing. When stratified by their effects on early childhood weight, DDR pathways were enriched among AAM genes that do not show a primary effect on early weight gain. Our findings suggest that the ovarian reserve, established during early fetal development, might signal centrally to influence the timing of puberty.

We address the considerable challenge of deriving biological insights from common variant signals^67^ by developing G2G (GWAS to Genes), an analytical pipeline that integrates a variety of data sources to enable gene prioritisation. While comprehensive experimental validation of G2G is infeasible, its utility is supported by the prioritisation from GWAS AAM data of many genes with known involvement in sex hormone regulation and rare monogenic or syndromic disorders of puberty, obesity and hormone function. The validity of G2G prioritised genes is also supported by evidence for their enrichment for dynamic expression in GnRH neurons during their late stage of embryonic migration, when they begin their integration into the hypothalamic neural network controlling puberty^68^. Furthermore, we provide experimental support for one novel high-scoring AAM gene, *GPR83*, which is co-expressed with, interacts with, and enhances *MC3R* function. Future studies should explore further the emerging role of brain expressed GPCRs in linking central nutritional sensing to reproductive function.

Finally, we provide one of the few examples to date of epistatic interaction between common and rare genetic variants. Linked to puberty timing by both common and rare variants, the transcription factor *ZNF483* has diverse binding sites across the genome. We infer that greater *ZNF483* binding promotes earlier AAM, whereas rare deleterious variants in *ZNF483* appear to abolish the influence of common genetic influence on puberty timing.

Together, these insights shed light on mechanisms, including early weight gain and adiposity, hormone secretion and response, and cellular susceptibility to DNA damage, that potentially explain the widely reported relationships between earlier puberty timing and higher risks of later life mortality, metabolic disease, and cancer.

## Data Availability

Cohorts should be contacted individually for access to their raw data. UK Biobank data are available on application (https://www.ukbiobank.ac.uk/enable-your-research/register). Summary statistics from the meta-analysis excluding 23andMe will be made available upon publication. Access to the full summary statistics including 23andMe results, can be obtained from 23andMe after completion of a Data Transfer Agreement (https://research.23andme.com/dataset-access/).

## Acknowledgements

This research has been conducted using the UK Biobank Resource under Application Number 9905. Other study specific acknowledgements can be found in the supplementary note.

## Competing interests

J.R.B.P. and E.J.G. are employed by Adrestia Therapeutics. D.J.T. is employed by Genomics PLC. D.L.C. and J.P.B. are employed by GSK. E.T. is employed by Pfizer. D.A.L. has received support from Roche Disgnostics and Medtroic Ltd for work unrelated to the research in this paper. T.D.S. is co-founder and stakeholder of Zoe Global Ltd. P.A.F. conducts research funded by Amgen, Novartis and Pfizer, he received Honoraria from Roche, Novartis and Pfizer. M.W.B. conducts research funded by Amgen, Novartis and Pfizer.

## Methods

### GWAS meta-analysis for age at menarche

Association summary statistics were collated from studies on age at menarche (AAM, predominantly recalled in adulthood) and genome-wide SNP arrays imputed to the 1000 Genomes reference panel or more recent (Supplementary Table 1). Genetic variants and individuals were filtered based on study-specific quality control metrics. In each study, genetic variants were tested for association with AAM in additive linear regression models, including as covariates: age and any study-specific variables, such as genetic principal components. Insertion and deletion polymorphisms were coded as “I” and “D” to allow harmonization across all studies. Association statistics for each SNP were then processed centrally using a standardized quality control pipeline^69^. Each variant was meta-analysed using a fixed-effects inverse-variance-weighted model using METAL^70^. This was done in two stages. First, summary statistics from studies within each stratum (i. ReproGen consortium studies, ii. reproductive cancer consortium studies, iii. East Asian studies) were meta-analysed and then filtered to include only variants present in more than half of the studies within each stratum. Second, strata-level results were meta-analysed with data from UK Biobank^71^, using ‘first instance’ data for AAM (field 2714), and 23andMe. Initially we performed a European-only analysis (N=632,955). This combined file was filtered to include only variants present in the UK Biobank and at least one other stratum. Variants were also filtered to include minor allele frequency (MAF) >= 0.1%. We then performed a second analysis, by adding the data from the East-Asian studies, and followed the same sample filtering steps and identification of independent signals (described below).

### Replication and explained variance

Independent replication of identified signals was performed in data from the Danish Blood Donors study^16^ (DBDS). The DBDS includes questionnaire recalled AAM data on 35,472 European women (“Age when menstruation started?”). Mean age at recall was 38.4 years (SD=12.9 years) and mean AAM was 13.1 years (SD=1.4 years). Indirect confirmation of AAM signals was sought by association with age at voice breaking (AVB) in men in UK Biobank^17^ (N=191,235 European men - data field 2385) and the 23andMe study^18^ (N=55,781 European men). For signals with missing data for either AVB dataset, we identified proxies using the UK Biobank White European dataset (within 1Mb of the reported signal and *R*^2^>0.6), choosing the variant with the highest *R*^2^ value. Given the smaller sample sizes of these cohorts, we performed a Binomial sign test for global replication. The variance explained by each lead AAM signal in the DBDS was calculated using the formula 2×f(1−f)β2a, where f denotes the variant MAF and βa is the effect estimate in additive models. Overall variance explained was calculated as the sum of individual variants.

### UK Biobank phenotype preparation

For downstream analyses in UK Biobank, we derived a AAM variable, using data from field 2714. To maximise sample size, individuals with missing or implausibly early or late ‘first instance’ AAM (<8 years or >19 years old) were imputed using data from the next available instance (if plausible). We also derived two binary traits to represent abnormally early (precocious) and delayed puberty. Early puberty was defined as AAM <10 years old (N=1,321). Delayed puberty was defined as AAM >15 years old (N=10,530). For comparison, women reporting AAM at 12 or 13 years were controls (N=81,950). All data analysis and visualisation were conducted in R (version 4.2.1, 2022-06-23), unless otherwise stated.

### Rare variant associations with age at menarche

To identify gene-level rare variant associations with AAM, we performed an exome-wide association study analysis (ExWAS) using whole-exome sequencing (WES) data on 222,283 UK Biobank women of European genetic-ancestry^72^. WES data processing and quality control was performed as described by Gardner *et al*.^30^. Individual gene burden tests were performed by collapsing variants with MAF <0.1% per gene according to their predicted functional consequence. We defined two functional categories of rare variants i) high confidence protein truncating variants (HC PTV) annotated using VEP^73^ and LOFTEE^74^ and ii) damaging variants (DMG) including HC PTVs plus missense variants with CADD score^19^ ≥25. We analysed a maximum of 17,885 protein-coding genes, each with at least 10 rare allele carriers in either of the two variant categories (*P*<1.54×10^−6^, 0.05/32,434 tests). Gene burden association tests were performed using BOLT-LMM^75^. Validity of the ExWAS analysis was indicated by the absence of significant association with the synonymous variant mask (Supplementary Figure 1) and low exome-wide inflation scores (λ-PTV =1.047 and λ-DMG =1.047). Where applicable, protein domains were annotated using information from UniProt^76^ and domain-level burden tests were then performed using linear models.

### Rare variant associations with other traits

We assessed the associations of any ExWAS AAM-associated genes in UK Biobank with a range of related phenotypes: age at natural menopause (based on field 3581), body mass index (BMI, field 21001), comparative body size age 10 (field 1687), adult height (field 50), comparative height age 10 (field 1697), and circulating IGF-1 concentrations (field 30770). We considered only the top AAM-associated variant mask for each gene. We also performed a similar look-up of these genes across a broader range of phenotypes using the AstraZeneca Portal^77^.

### Rare variants in IHH panel app genes

We selected high evidence (“green”) genes with an established monoallelic/X-linked mode of inheritance from the routine clinical investigation Genomics England gene panel for idiopathic hypogonadotropic hypogonadism (IHH). At the time of the study, this included four genes, *ANOS1, CHD7, FGF8* and *WDR11*. We performed a look-up of these genes in the UK Biobank WES data for AAM (N=222,283) and VB (N=178,625) considering only HC PTVs with MAF <0.1%. We also extracted the phenotype of individual carriers. As in the ExWAS analysis, normal pubertal timing was defined in women as AAM between 10-15 years of age^1^ and in men as AVB at an “about average age” (UK Biobank data field 2385).

### Polygenic score calculation

We calculated a genome-wide polygenic score (PGS) for AAM using lassosum^31^. To keep PGS generation independent of PGS testing, we generated the PGS using our European-ancestry GWAS data excluding UK Biobank. We randomly selected 25,000 unrelated Europeans in UK Biobank to generate the LD (Linkage Disequilibrium) reference. The resulting PGS was standardised, by subtracting the mean and dividing by the standard deviation.

We divided the PGS into 100 centiles, and calculated the mean AAM for each PGS centile, as well as PGS centile-specific odds ratios for precocious or delayed AAM (as defined above) compared to individuals in the 50^th^ centile of the PGS. We also calculated the mean PGS for each completed whole year of AAM.

We next tested whether carriage of ExWAS AAM-associated rare variants modifies the influence of the PGS on AAM. We performed linear models that included interaction terms [PGS × rare variant carrier status] in the subsample of unrelated white-European UK Biobank women with WES, PGS and AAM data (N=187,941). To test for chance effects due to low sample size, we randomly subsampled non-carriers to a sample size equivalent to that of carriers and compared this distribution of AAM to that observed in carriers.

A PGS comprising the 882 available lead AAM SNPs or their proxies (out of the 935 independent AAM signals from the EUR-MA) was computed in 3,140 girls with available imputed GWAS data from the ALSPAC study. Linear or logistic regression models for continuous AAM, early AAM (−2 SDs, corresponding to <10.38 years), and delayed AAM (+2 SDs, corresponding to >14.95 years) were tested, controlling for the first 20 genetic PCs. Other models assessed the predictive performance of BMI at age 8 years, and mother’s AAM; finally a model including all predictors as covariables was calculated. The predictive performance of each model was evaluated by the *R*^2^ metric for continuous AAM and by the area under the receiver operating characteristic curve (AUROC) for binary AAM outcomes.

### GWAS to Genes (G2G) pipeline

#### Mapping GWAS signals to genes

To perform signal selection, GWAS AAM summary statistics were filtered to remove variants with MAF <0.1%. The remaining variants were merged with allele information from UK Biobank to provide the genomic sequence for any missing alleles. Genome-wide significant signals (*P*<5×10^−8^) were selected initially based on proximity (in 1Mb windows). Secondary signals within these windows were then identified using approximate conditional analysis (GCTA^78^), using an LD reference panel derived from 25,000 randomly selected UK Biobank participants. Secondary signals were defined as uncorrelated (*R*^2^<0.05) with another signal and without an overt change in their AAM association between baseline and conditional models (change in beta <20% or change in *P*-value by less than four orders of magnitude). Primary and secondary AAM signals were further checked for pairwise LD within 10 Mb windows using plink (v1.90b6.18)^79^ and only independent signals (*R*^2^<0.05) were retained, prioritising distance-based signals in the case of linkage. Signal selection was performed first using the European-ancestry GWAS meta-analysis, and then supplemented by any signals identified by the All-ancestry GWAS meta-analysis that were uncorrelated (*R*^2^<0.05) with any European-ancestry signal.

Independent GWAS AAM signals were examined for proximal genes, defined as those within 500kb up- or downstream of the genes start or end sites, using the NCBI RefSeq gene map for GRCh37 (via http://hgdownload.soe.ucsc.edu/goldenPath/hg19/database/).

#### Colocalization with expression or protein QTL data

Tissue enrichment for GWAS associations was performed using LD score regression applied to tissue-specific expression (LDSC-SEG)^80^ and tissue-specific annotations from GTEx, accessed via https://github.com/bulik/ldsc/wiki/Cell-type-specific-analyses. Significantly enriched tissues (*P*<0.05) were then included in colocalization analyses with the tissue-specific and cross-tissue meta-analysed GTEx eQTL data (V7^60^, available via https://gtexportal.org and using the fixed-effects summary statistics for the latter), in addition to data from the eQTLGen^81^ and Brain-eMeta^59^ studies.

Including genomic variants with at least suggestive association with AAM (GWAS *P*<5×10^−5^), we applied Summary data-based Mendelian Randomization and Heterogeneity in Independent Instruments (SMR-HEIDI, version 0.68^82^) and the Approximate Bayes Factor (ABF) method in the R package “coloc” (version 5.1.0^83^). For the former, we considered gene expression to be influenced by the same GWAS AAM variant if the FDR-corrected SMR test *P*<0.05 and HEIDI test *P*>0.001. For the latter, genomic regions were defined as ±500 kb around each gene and loci exhibiting a H4 posterior probability >0.75 were considered to show evidence of colocalization. We also tested for colocalization between GWAS AAM variants and pQTLs using data from the Fenland study^84^ and using the same procedure as above. It is important to note that colocalization analysis cannot determine causal relationships or the direction of causality between the two phenotypes.

#### Mapping GWAS signals to enhancers and coding variants

For genes proximal to (within 500 kb) GWAS AAM signals, we calculated genomic windows of high LD (*R*^2^>0.80) around each signal and mapped these to the locations of known enhancers for the genes, using activity-by-contact (ABC) enhancer maps^34^. This was done across the 131 available cell/tissue types and genes were matched to enhancers only in the tissues/cells where they were actively expressed.

We also checked whether GWAS AAM signals were in LD (*R*^2^>0.80) with any coding variants within the paired genes and what the predicted consequence of those coding variants, using SIFT^85^ and POLYPHEN^86^.

#### Gene-level GWAS associations with AAM

We performed a gene-level MAGMA analysis^35^, which collapses common GWAS variants within each gene and calculates aggregate gene-level associations with the outcome trait, as described by de Leeuw et al.^35^. To enhance validity of this approach, we restricted the analysis to include only coding variants. Genes with FDR-corrected MAGMA *P*<0.05 were considered associated with AAM.

Finally, we used the Polygenic Priority Score (PoPS^36^), which is a similarity-based gene prioritisation method and uses cell-type specific gene expression, biological pathways, and protein-protein interactions to prioritise likely causal genes from GWAS data. At each locus, the gene with the numerically highest PoPS score was determine to be the PoPS-prioritised gene.

#### Calculation of G2G scores

From the above analyses, gene-level results were scored for each of the six sources as follows:

1. **Closest gene**. Gene proximity to a GWAS signal is a good predictor of causality^37^. The genes closest to each AAM signal (if also within 500 kb) were assigned. All genes with an intragenic signal were assigned as closest. Closest genes were scored 1.5 points.
2. **eQTL colocalisation**. Genes with evidence of eQTL colocalisation via both SMR-HEIDI and coloc were scored 1.5 points. Genes with evidence of colocalisation via only one of these received 1.0 point. A further (1.0) point was assigned to genes if the most likely shared causal variant between eQTL and GWAS AAM was independent of the proximal GWAS signal (*R*^2^<0.05).
3. **pQTL colocalisation**. The same scoring as in (2) was applied to pQTL analyses.
4. **Coding variants**. As the evidence was overlapping for coding variant gene-level MAGMA analysis and signals correlated with coding variants, these analyses were scored concomitantly. Genes with an FDR-corrected MAGMA *P*<0.05 were scored 0.5 points. Genes containing coding variants of deleterious or damaging predicted consequence in LD with GWAS AAM signals were scored 1.0 point, or only 0.5 points if the coding variants were predicted to be benign or tolerated.
5. **ABC enhancers**. Genes targeted by enhancers which overlapped with or were correlated with GWAS AAM signals were scored 1.0 point.
6. **PoPS**. PoPs prioritised genes at each locus were scored 1.5 points.

G2G scores for each gene-signal pair were calculated as the sum of scores from these six sources. Genes that scored >0 points and were located within 500 kb of a GWAS AAM signal were considered further. To account for confounding due to large LD blocks, G2G scores were adjusted for signal LD window size (defined as the genomic distance containing variants with pairwise *R*^2^>0.50 with the lead SNP) using linear regression models.

For genes proximal to more than one GWAS AAM signal (and hence with multiple G2G scores), the signal with the most concordant sources for that gene (highest residual G2G score) was retained and a further (1.0) point was added to reflect evidence from multiple signals. This resulted in a unique summarised G2G score for each included gene. To account for confounding due to gene size, G2G scores were further adjusted for gene length using linear regression models. The resulting residuals were considered to be the final G2G scores.

To prioritise likely causal AAM genes, all G2G scored genes (i.e., highlighted as potentially causal by at least one source) were ranked and also allocated a G2G centile position. In addition, the number of concordant predictors (sources) for each gene were noted (range: 1 to 6 sources). Finally, to reflect uncertainty due to multiple high scoring genes for the same signal, genes were flagged if they were proximal (within 1 Mb) to other genes with a similar G2G score (within 0.5 points or greater, and highlighted by at least the same number of sources).

#### High-confidence AAM genes

Independent GWAS signals from the All- and the European-ancestry meta-analyses were annotated with their top G2G scoring gene, using corresponding GWAS data (i.e., European analysis signals were annotated with genes from the European G2G, etc.). Genes implicated by at least two concordant sources were considered to be high-confidence AAM genes.

High-confidence AAM genes were functionally annotated using STRING^87^. Links to rare monogenic disorders were annotated from the Online Mendelian Inheritance in Man (OMIM) database (via Online Mendelian Inheritance in Man, OMIM®. McKusick-Nathans Institute of Genetic Medicine, Johns Hopkins University (Baltimore, MD), accesed November 2022. World Wide Web URL: https://omim.org/). Finally, we used GTEx, a publicly available resource of tissue-specific gene expression, to lookup the tissue expression of 1080 AAM genes highlighted by G2G^60^.

### ZNF483 genome-wide binding analysis

We used fGWAS (v.0.3.6^32^), a hierarchical model for joint analysis of GWAS and genomic annotations, to test for enrichment of GWAS AAM signals among ZNF483 transcription factor binding sites. fGWAS models a maximum likelihood parameter estimate for enrichment of a transcription factor (in this case ZNF483). To perform this, we annotated the European-ancestry GWAS AAM summary statistics with the ZNF483 binding sites from the ENCODE ChIP-seq data derived from human HepG2 cell line (ENCSR436PIH).

We also used Signed LD Profile regression (SLDP, https://github.com/yakirr/sldp, Reshef *et al*. ^33^) to explore the directional effect of ZNF483 function on AAM. We tested whether alleles that are predicted to increase the binding of ZNF483 have a combined tendency to increase or decrease AAM. SLDP requires signed LD profiles for ZNF483 binding, a signed background model and reference panel in a SLDP compatible format. We used a 1000 Genomes Phase 3 European reference panel containing approximately 10M SNPs and 500 individuals.

### Clustering of AAM signals by early childhood body weight

We analysed repeated measurements of early childhood body weight from the MoBa cohort study^52,88^ to investigate the relationship between early growth and puberty timing. Childhood body weight values were extracted from the study questionnaires for 12 different time-points from birth to age 8 years using previously reported exclusion criteria^52^. Weight values were standardized and adjusted for sex and gestational age using the generalized additive model for location, scale and shape (GAMLSS; v5.1-7, via www.gamlss.com) in R (v3.6.1) as previously reported^52^ with the exception that a Box-Cox t distribution was used to standardize body weight values (instead of the log-normal distribution used for BMI)^52^. GWAS for these traits was performed using BOLT-LMM (v2.3.4) as previously reported^52^.

We performed Mendelian randomisation (MR) analyses to assess the likely causal effects of AAM on childhood weight at each time-point^89^. As instrumental variables (IVs), we used all 1080 AAM-associated lead SNPs individually. As outcome data, we used childhood weight at the 12 time-points. For SNPs with missing outcome data, we identified proxies within 1 Mb and *R*^2^>0.6, choosing the variant with the highest *R*^2^ value, using a random selection of 25,000 unrelated European-ancestry UK Biobank individuals for the LD reference. Genotypes at all variants were aligned to the AAM-increasing allele. We used inverse-variance weighted (IVW) MR models, as this has the greatest statistical power^90^.

Next, we stratified the 1080 AAM lead SNPs by their effects on early childhood weight used a k-means clustering approach for longitudinal data^91^. We performed five different models with k-means for k∈{2,3,4,5,6} clusters 20 times each. To find the optimal partition, we used the “nearlyAll” option which uses several different initialisation methods in alternation. As the assumption of homoscedasticity was not met, we used the Carolinski-Harabatz criterion, a non-parametric quality criterion, to derive the optimal number of clusters.

We then performed additional MR analyses, combining AAM signals within each identified cluster as IVs and, as the outcomes, childhood weight at each time-point and also adult BMI (on N=450,706 UK Biobank participants). We grouped together ‘high early weight’ and ‘moderate early weight’ AAM SNPs into a single IV to maximise power.

### Biological pathway enrichment analysis

We performed gene-centric biological pathway enrichment analysis using g:Profiler (via the R client “gprofiler2”, version 0.2.1^53^). We used a filtered set of GO pathways (accessed on the 21/02/2023), focusing on GO:BP, KEGG and REACTOME, and restricted the analysis to those pathways with 1000 genes or fewer, reasoning that these are more biologically specific. Pathway enrichment analyses were performed using the set of 660 high-confidence AAM genes, and repeated when stratified by their effects on early childhood weight (see above). Pathways with Bonferroni corrected *P*<0.05 were considered to be associated with AAM.

As the pathways derived from overlapping sources, we clustered the AAM-associated pathways to aid interpretation. Clustering was based on shared AAM genes across pathways. We used a “complete” clustering algorithm and a custom distance calculated as [one minus the proportion of the overlap between any two pathways relative to the pathway with the smaller overlap]. Thus, between two pathways a value of 0 indicates that all the shared AAM genes in the pathway with fewer genes are also enriched in the other pathway. To define clusters, we chose an arbitrary overlap value of 0.5, which indicates that pathways in the same cluster share 50% or more of their AAM genes.

Each pathway cluster was annotated by i) the pathway with the most significant enrichment, ii) the pathway with the highest proportion of AAM genes, iii) biological coherence of the pathways, and iv) shared genes common to all included pathways. We considered that pathways were overlapping between the total AAM gene set and the two early-weight subgroups if there were common pathways across either i) the most significant pathway or ii) the pathway with the highest proportion of AAM-associated genes.

### Expression of AAM genes in GnRH neurons

We tested for enrichment of AAM-associated genes in RNAseq data from embryonic GnRH mouse neurons [manuscript in preparation]. All expressed genes were sorted into different expressional trajectories, based on shared dynamic expression profiles across three developmental stages (early, intermediate or late), as described by Pitteloud et al. [manuscript in preparation]. We tested for enrichment of AAM-associated genes (from our European-ancestry GWAS meta-analysis) in any identified trajectory, using MAGMA^35^ with custom pathways. As a sensitivity test, we used Fisher’s Exact test to confirm over-representation of AAM-associated genes within each trajectory.

### Colocalization of AAM signals with BMI and menopause

To explore the shared genetic architecture between AAM, age at natural menopause (ANM) and adult BMI, we performed a colocalization analysis for each of the 1080 AAM signals. ANM GWAS summary statistics were from reported ReproGen data on ∼250,000 women of European ancestry^62^. Adult BMI GWAS summary statistics were derived from 450,706 individuals in UK Biobank. For AAM signals with missing outcome GWAS data, we identified proxies within 1 Mb and with an *R*^2^>0.6 using our 25,000 participant UK Biobank LD reference. We applied both Bonferroni correction (*P*≤0.05/1080=4.6×10^−5^) for association with the outcome trait, and a posterior probability (PP) of colocalization PP>0.5.

The same approach was applied in the opposite direction, by testing ANM signals identified in the most recent ReproGen GWAS^62^ for association with AAM. ANM signals were highlighted if they passed Bonferroni correction (*P*≤0.05/290=1.7×10^−4^) for association with AAM. As ANM signals are well-established to be enriched for DNA damage repair genes (DDR), we built a comprehensive list of DDR genes, integrating five different sources: i) an expert curated DDR gene list (“Broad DDR”) from the laboratory of Professor Stephen Jackson, this list encompasses a range of related pathways: DNA repair genes, broader DNA damage response genes (such as damage-induced chromatin remodelling, transcription regulation or cell cycle checkpoint induction); and general maintenance of genome stability (such as genes involved in DNA replication); ii) a second expert curated list previously reported^62^, assembled by John Perry, Eva Hoffmann and Anna Murray; iii) genes listed in the REACTOME^92^ “DNA Repair” pathway (R-HSA-73894); iv) genes listed in the Gene Ontology “DNA Repair” pathway (GO:0006281); and v) genes listed in the Gene Ontology^93^ “Cellular response to DNA damage stimulus” (GO:0006974).

### GPR83-MC3R interaction

#### Brain expressed GPCRs

We tested whether any brain-expressed G-protein coupled receptors (GPCRs) were implicated by GWAS AAM associations (G2G gene scores). We tested a curated list of brain-expressed GPCRs (Stephen O’Rahilly, personal communication): ACKR1, ACKR2, ACKR3, ACKR4, ADRB1, ADRB2, ADRB3, AGTR1, AGTR2, BRS3, C5AR1, C5AR2, CALCR, CASR, CCKAR, CCR1, CCR10, CCR2, CCR3, CCR4, CCR5, CCR6, CCR7, CCR9, CCRL2, CNR1, CNR2, CXCR1, CXCR2, CXCR3, CXCR4, CXCR6, DRD1, DRD2, DRD3, DRD4, DRD5, EDNRA, EDNRB, FFAR1, FFAR2, FFAR3, FFAR4, FPR1, FPR2, FPR3, FSHR, GALR1, GALR2, GALR3, GHRHR, GHSR, GIPR, GLP1R, GLP2R, GNRHR, GPER1, GPR1, GPR12, GPR15, GPR17, GPR18, GPR19, GPR20, GPR22, GPR25, GPR26, GPR27, GPR3, GPR34, GPR35, GPR37, GPR39, GPR4, GPR42, GPR45, GPR52, GPR55, GPR6, GPR61, GPR62, GPR63, GPR75, GPR78, GPR82, GPR83, GPR84, GPR85, GPR87, GPR88, GRM1, GRM2, GRM3, GRM4, GRM5, GRM6, GRM7, GRM8, GRPR, HCAR1, HCAR2, HCAR3, HRH1, HRH2, HRH3, HRH4, LGR4, LGR5, LGR6, LPAR1, LPAR2, LPAR3, LPAR4, LPAR5, LPAR6, MC3R, MC4R, MC5R, MCHR1, MCHR2, NMBR, NMUR1, NMUR2, NPSR1, NPY1R, NPY2R, NPY4R, NPY5R, NPY6R, OXER1, OXGR1, P2RY1, P2RY2, P2RY4, P2RY6, P2RY8, PRLHR, PTAFR, PTH1R, PTH2R, QRFPR, RGR, RXFP1, RXFP2, S1PR1, S1PR2, S1PR3, S1PR4, S1PR5, SCTR, SSR1, SSR2, SSR3, SSR4, TSHR, VIPR1, VIPR2, VN1R1, VN1R2, VN1R5 and XCR1. For any GPCR scored by our G2G AAM pipeline, colocalisation was tested between GWAS signals for AAM and adult BMI (colocalisation methods as described above).

#### Cell culture and transfection

To investigate the effect of *GPR83* of MC3R signalling, we performed *in vitro* assays in transiently transfected HEK293 cells maintained in Dulbecco’s modified eagle medium (high glucose DMEM, GIBCO, 41965) supplemented with 10% fetal bovine serum (GIBCO, 10270), 1% GlutaMAX^™^ (100X) (GIBCO, 35050), and 100 units/mL penicillin and 100 mg/mL streptomycin (Sigma-Aldrich, P0781). Cells were incubated at 37°C in humidified air containing 5% CO2 and transfections were performed using Lipofectamine 2000 (GIBCO, 11668) in serum-free Opti-MEM I medium (GIBCO, 31985), according to the manufacturer’s protocols. The plasmids used encode the C-FLAG-tagged human *GPR83* WT (NM_016540.4) or N-FLAG-tagged human MC3R WT (NM_019888.3) ligated into pcDNA3.1(+) (Invitrogen).

#### Bioluminescence Resonance Energy Transfer (BRET) to measure dimerization

Heterodimerization between *GPR83* and *MC3R* was quantified using BRET1 in titration configuration. Briefly, 12,000 HEK293 cells seeded in 96-well plates were transfected with a constant dose of MC3R-RlucII plasmid (0.5 ng/well) and increasing doses of *GPR83*-Venus plasmids, or soluble (s) Venus as negative control. All conditions were topped up with empty vector (pcDNA3.1 (+)) to a total of 100 ng plasmid/well. Twenty-four hours post transfection, cells were washed once with Thyrode’s buffer and total Venus fluorescence was measured in a Spark 10M Microplate reader (Tecan) using monochromators (excitation 485 ± 20 nm, emission 535 ± 20 nm). BRET was quantified 10 minutes after the addition of coelenterazine H (NanoLight Technology, 2.5 mM). netBRET was calculated as [(absorbance at 533 ± 25 nm/absorbance at 480 ± 40 nm)] – [background (absorbance at 533 ± 25 nm/absorbance at 480 ± 40 nm)], with the background corresponding to the signal in cells expressing the RlucII protomer alone under similar conditions. Data on the X-axis represent the ratio between acceptor (Venus) fluorescence and donor (RlucII) luminescence. Representative data are from four independent experiments.

#### Time-resolved cAMP assay

Measurement of ligand-induced cAMP generation in HEK293 cells transiently expressing either *MC3R* or both *MC3R* and *GPR83* was performed using GloSensor™ cAMP biosensor (Promega), according to manufacturer’s protocol. Briefly, 12,000 cells were seeded in white 96-well poly-D-lysine-coated plates. After 24 hours, cells were transfected with both 100 ng/well of pGloSensorTM-20F cAMP plasmid (Promega, E1171) and 30 ng/well of each plasmid encoding either MC3R or MC3R and GPR83, using Lipofectamine 2000 (GIBCO, 11668). All conditions were topped up with empty vector (pcDNA3.1 (+)) to a total of 160 ng plasmid/well. The day after transfection, cell media were replaced by 90 mL of fresh DMEM with 2% v/v GloSensorTM cAMP Reagent (Promega, E1290) and incubated for 120 min at 37°C. Firefly luciferase activity was measured at 37°C and 5% CO2 using a Spark 10M microplate reader (Tecan). After initial measurement of the baseline signal for 10 min (30 seconds intervals), cells were stimulated with 10 mL of 10x stock solution of the MC3R agonist NDP-aMSH (final concentration 1 mM) and real-time chemiluminescent signals were quantified for 25 minutes (30 seconds intervals). In each experiment, a negative control using mock transfected cells (empty pcDNA3.1(+) plasmid) was assayed. The area under the curve (AUC) was calculated for each cAMP production curve considering total peak area above the baseline calculated as the average signal for mock pcDNA3.1(+)-transfected cells. For data normalization, the AUC from mock transfected cells was set as 0 and the AUC from WT MC3R was set as 100%. Results are from six independent experiments.

#### Genetic epistasis between GPR83 and MC3R

To corroborate the *in vitro* interaction, we tested for evidence of a specific epistatic interaction between AAM GWAS signals at *GPR83* (rs592068-C) and *MC3R* (rs3746619-A). We extracted genotypes for these SNPs in white-European unrelated UK Biobank women (N=204,303). After adjusting AAM for standard covariates (GWAS chip, age, sex, PC1-10), we modelled the interaction between genotype dosages at the two signals using a linear model.

## Notes

### Funding Statement

Study-specific funding information can be found in the supplementary note.

