## Supplementary Figures and Information for "Understanding the genetic complexity of puberty timing across the allele frequency spectrum"

### Supplementary information

#### Supplementary Figures

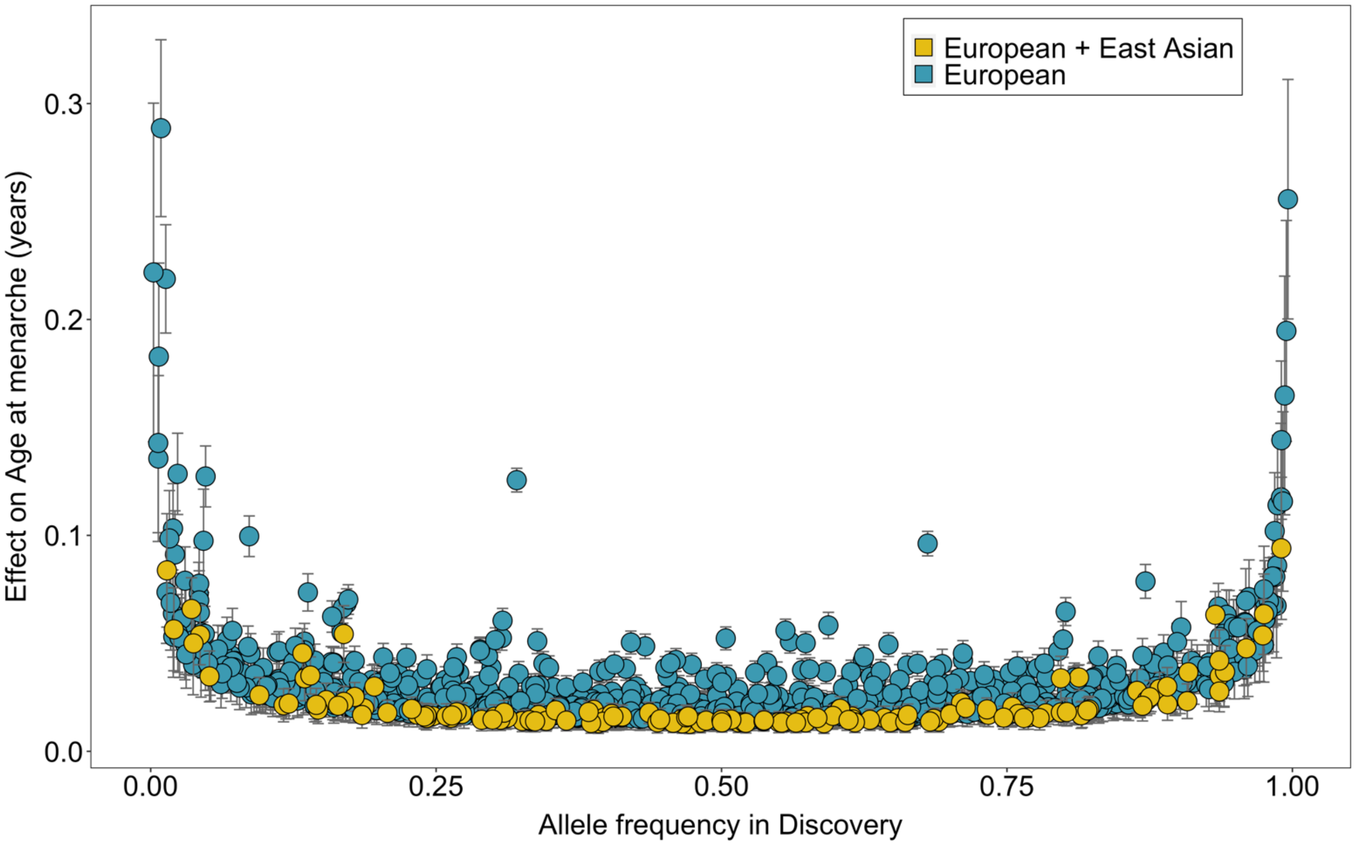

**Supplementary Figure 1 | Distribution of effect sizes across 1080 independent signals with age at menarche (AAM).** For each signal the effect size estimate and 95% confidence intervals are shown from the corresponding Discovery analysis, i.e. from the European-only or the All-ancestry meta-analysis, as indicated by point colours. Estimates are aligned towards the menarche-increasing alleles. Extended data are shown in Supplementary Table 2.

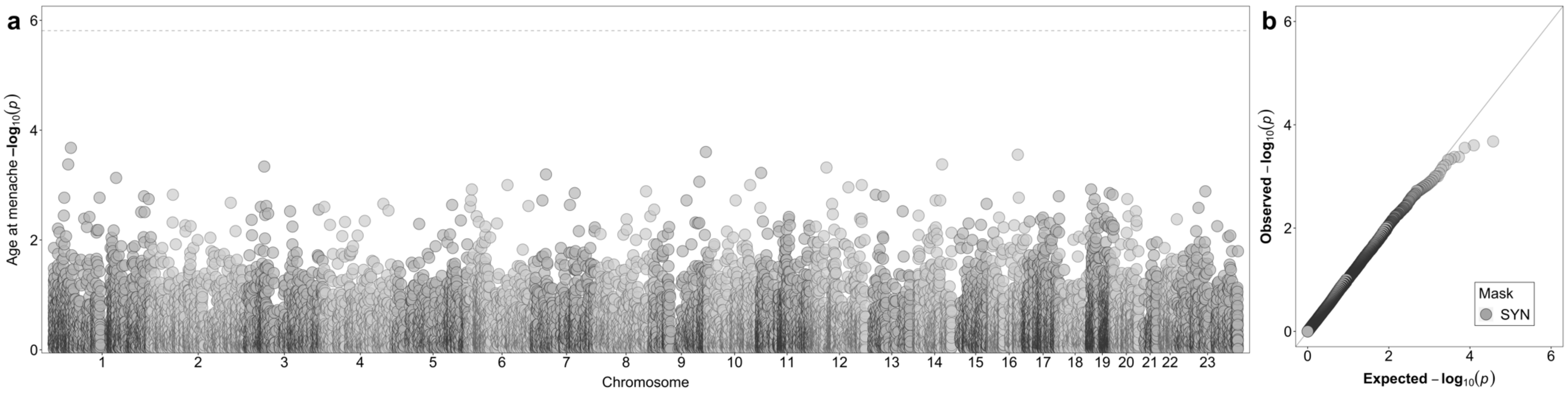

**Supplementary Figure 2 | Synonymous variant associations with AAM.** (a) Manhattan plot showing the gene burden associations for the synonymous variant mask towards AAM. The horizontal line indicates the exome-wide significance threshold ($P<1.54 \times 10^{-6}$). (b) Quantile-quantile (QQ) plot of the gene burden associations for the synonymous variant mask.

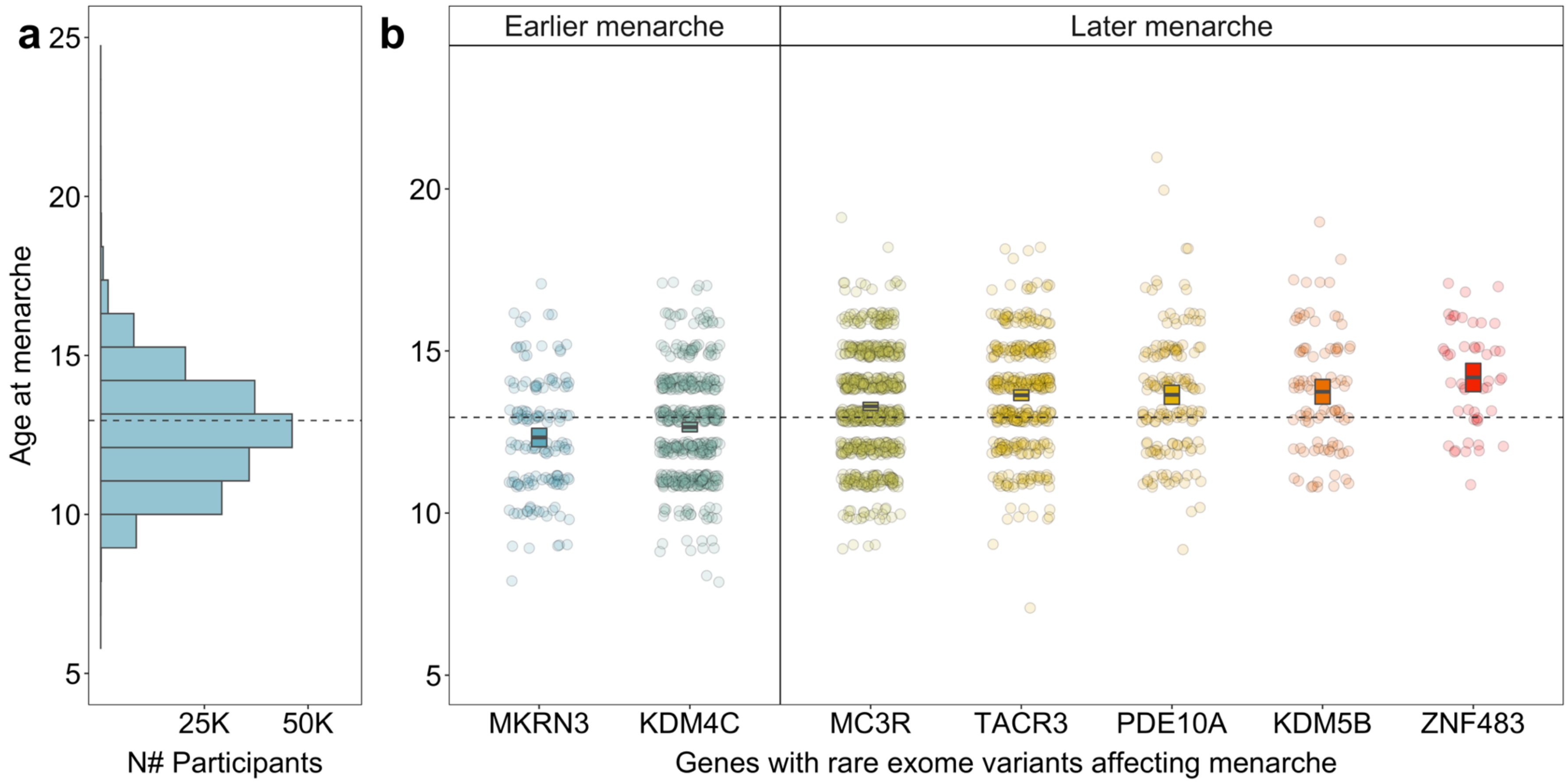

**Supplementary Figure 3 | Distribution of AAM in UK Biobank.** (a) Reported AAM from all white European unrelated women (N=187,941) (b) Women carrying qualifying rare variants in the associated genes. The horizontal dotted line indicates mean age at menarche among non-carriers (N=185,929). Mean and 95% CIs for each carrier group are indicated by horizontal bars and boxes.

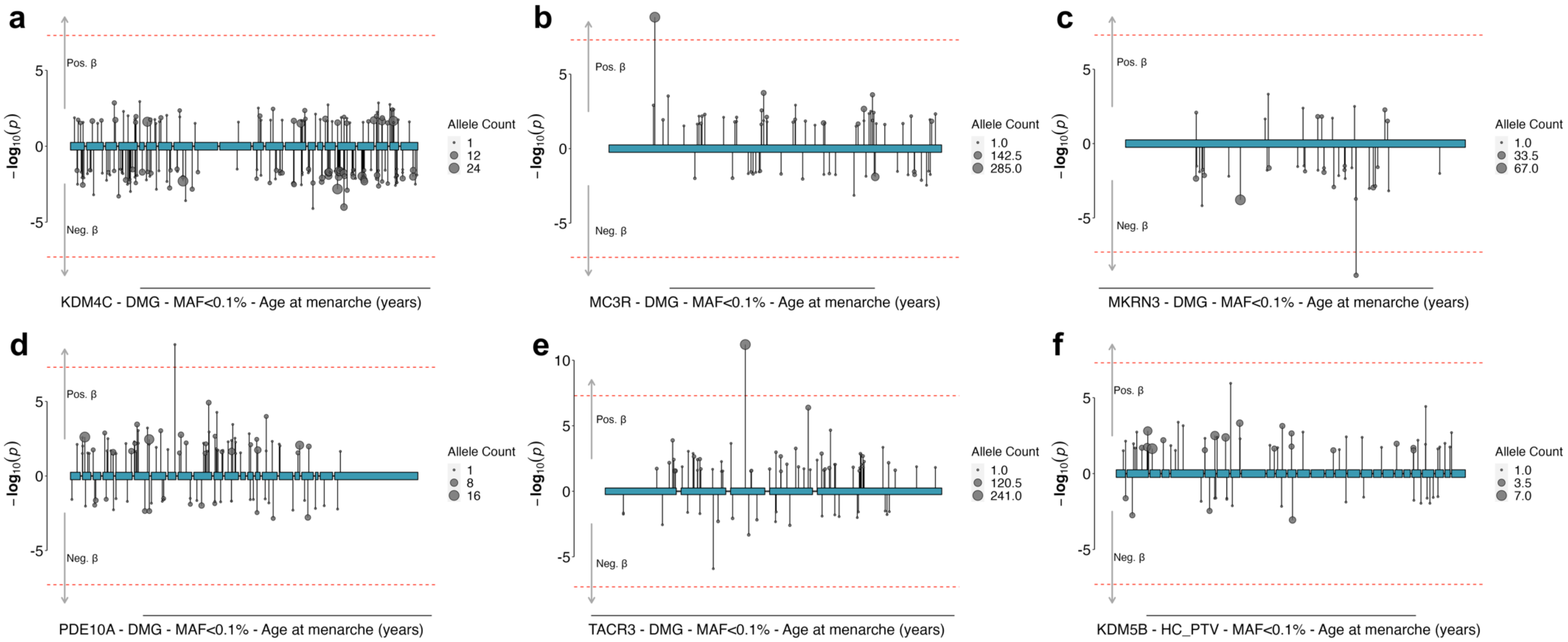

**Supplementary Figure 4 | Variant-level associations with AAM in identified genes in UK Biobank.** Rare exome variant associations with AAM for variants within KDM4C (a), MC3R (b), MKRN3 (c), PDE10A (d), TACR3 (e), KDM5B (f). Variant collapsing masks included variants with a minor allele frequency (MAF) < 0.1% and annotated as either high-confidence protein truncating variants (HC_PTV) or HC_PTV plus missense variants with a high CADD score (>=25, denoted DMG). Each variant association is represented by a circle and vertical line: the line length indicates the P-value (-log10), in the direction of its effect on AAM in carriers of the rare allele, and the circle size indicates the number of carriers of each variant (i.e. the allele count). Exons are indicated by the blue boxes and connected by the intron line.

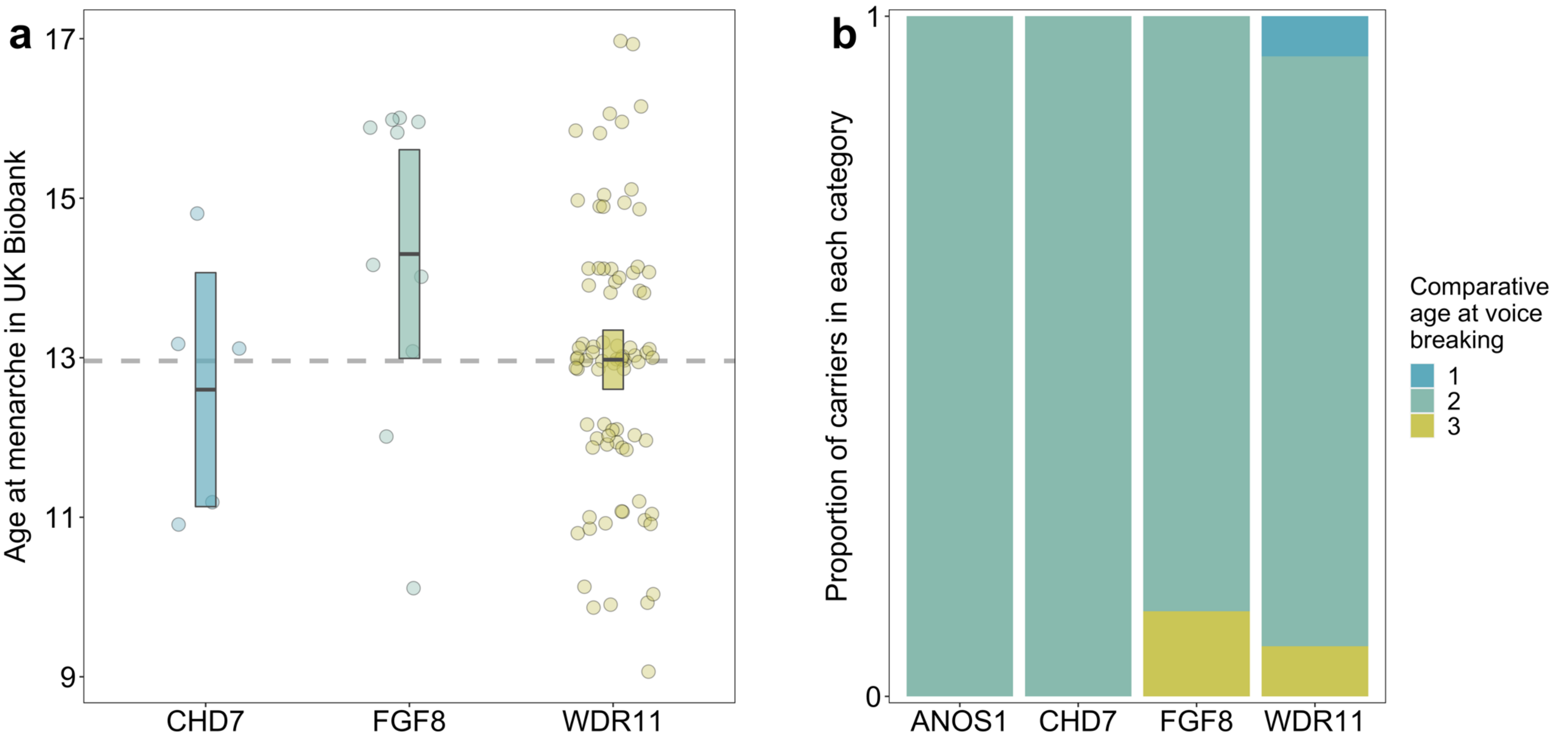

**Supplementary Figure 5 | Puberty onset in carriers of IHH panel genes in UK Biobank.** (a) Distribution of age at menarche in UK Biobank white European unrelated women carrying qualifying rare variants in IHH panel genes. The horizontal dotted line indicates mean AAM among non-carriers (N=185,929). Mean and 95% CI for each carrier group are indicated by horizontal bars and boxes. (b) Comparative age at voice breaking in men carrying qualifying rare variants in IHH panel genes. 1 indicates self-reported younger than average age at voice breaking, 2 indicates average age and 3 indicates later than average age.

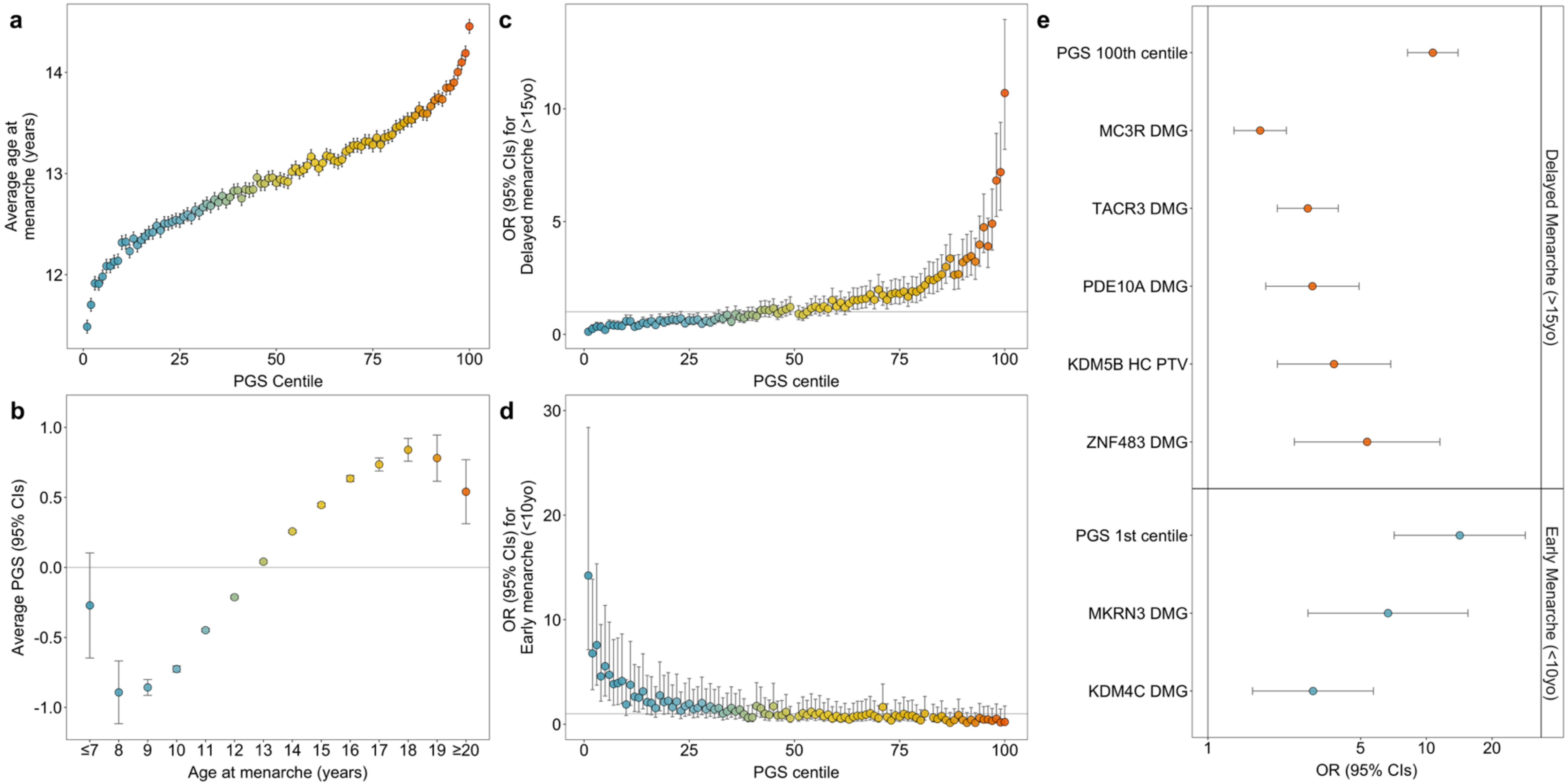

**Supplementary Figure 6 | Polygenic score (PGS) associations with AAM.** The PGS for AAM was derived using summary statistics from the EUR-MA and excluding UK Biobank, and then applied to white European, unrelated women in UK Biobank (N=187,941). (a) Means with 95% CIs of AAM by PGS centile (N=1,880 per centile). (b) Means with 95% CIs of standardised PGS by AAM. (c-d) Associations of each PGS centile compared to the 50th PGS centile with (c) delayed menarche (menarche after 15 years) and (d) early menarche (menarche before 10 years). Controls were women who reported menarche at 12 or 13 years. PGS centiles with fewer than 2 participants were omitted. (e) Gene burden associations between qualifying variants in the exome-identified genes compared to extreme PGS centiles, with delayed or early menarche (defined as above). Extended data are shown in Supplementary Tables 8-10.

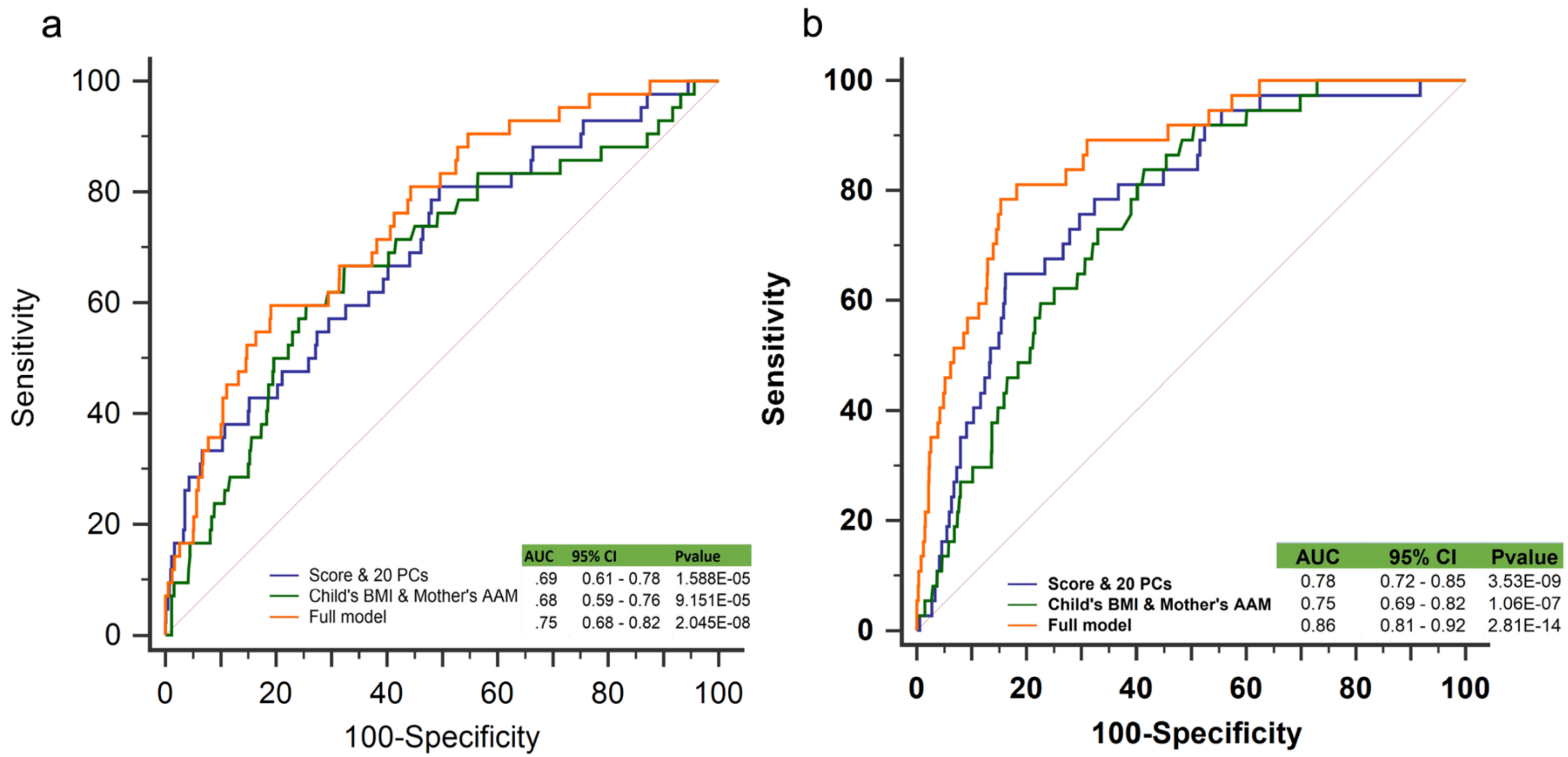

**Supplementary Figure 7 | Receiver operating characteristic (ROC) curve for predicting extremes of menarche age in the ALSPAC study.** ROC graphs of the genetic, clinical, and combined predictor on (a) early AAM and (b) delayed AAM in the ALSPAC study (N= 3,140). Extended data are shown in Supplementary Table 11.

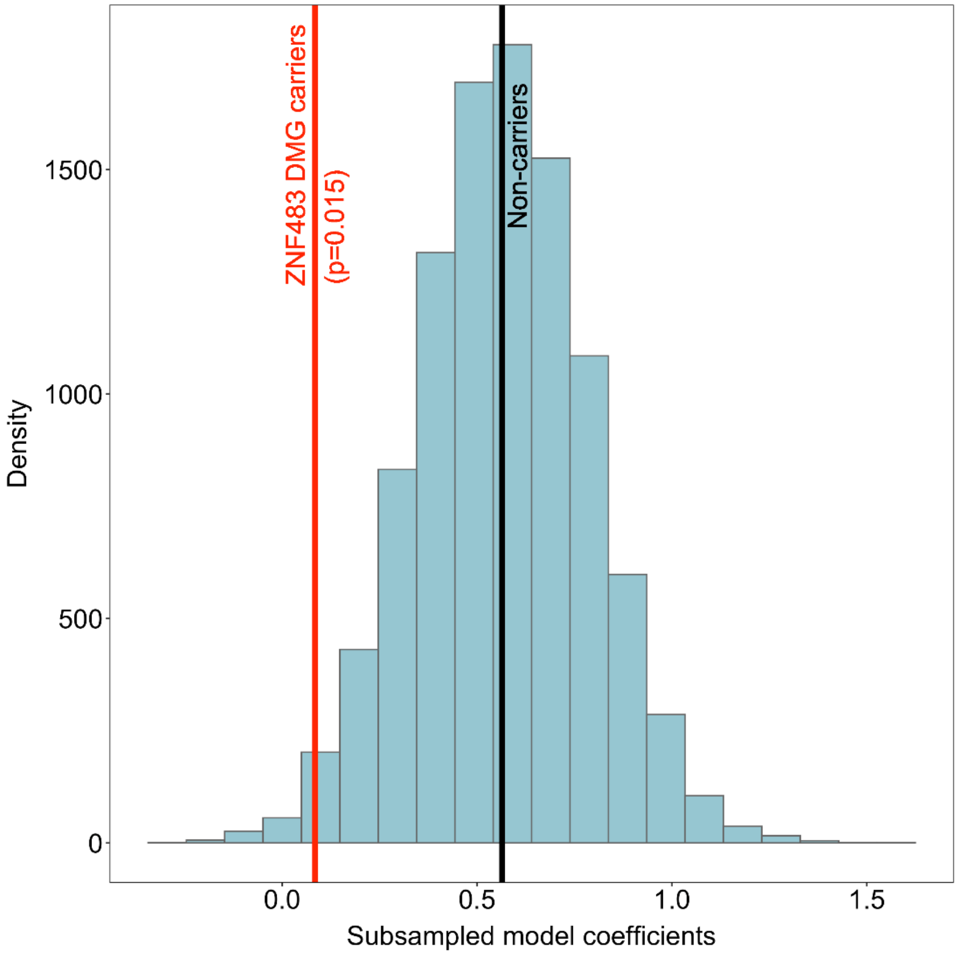

**Supplementary Figure 8 | Epistatic interaction between the PGS and carriage of rare damaging variants in *ZNF483* on AAM.** Blue columns indicate the beta coefficients for the PGS-AAM association among 10,000 random subsamples of 49 white European unrelated female non-carriers in UK Biobank. The beta coefficient from the full sample of non-carriers is indicated by the black vertical line (0.564 years per SD, SE 0.003, $P<2x10^{-16}$) and the beta coefficient seen in *ZNF483* variant carriers is indicated by the red vertical line (difference in coefficients: -0.480, SE 0.214, P=0.025). The probability of observing a beta coefficient smaller than that in *ZNF483* variant carriers was estimated as the proportion of subsampled coefficients that were smaller than 0.084 (i.e., 0.564-0.480).

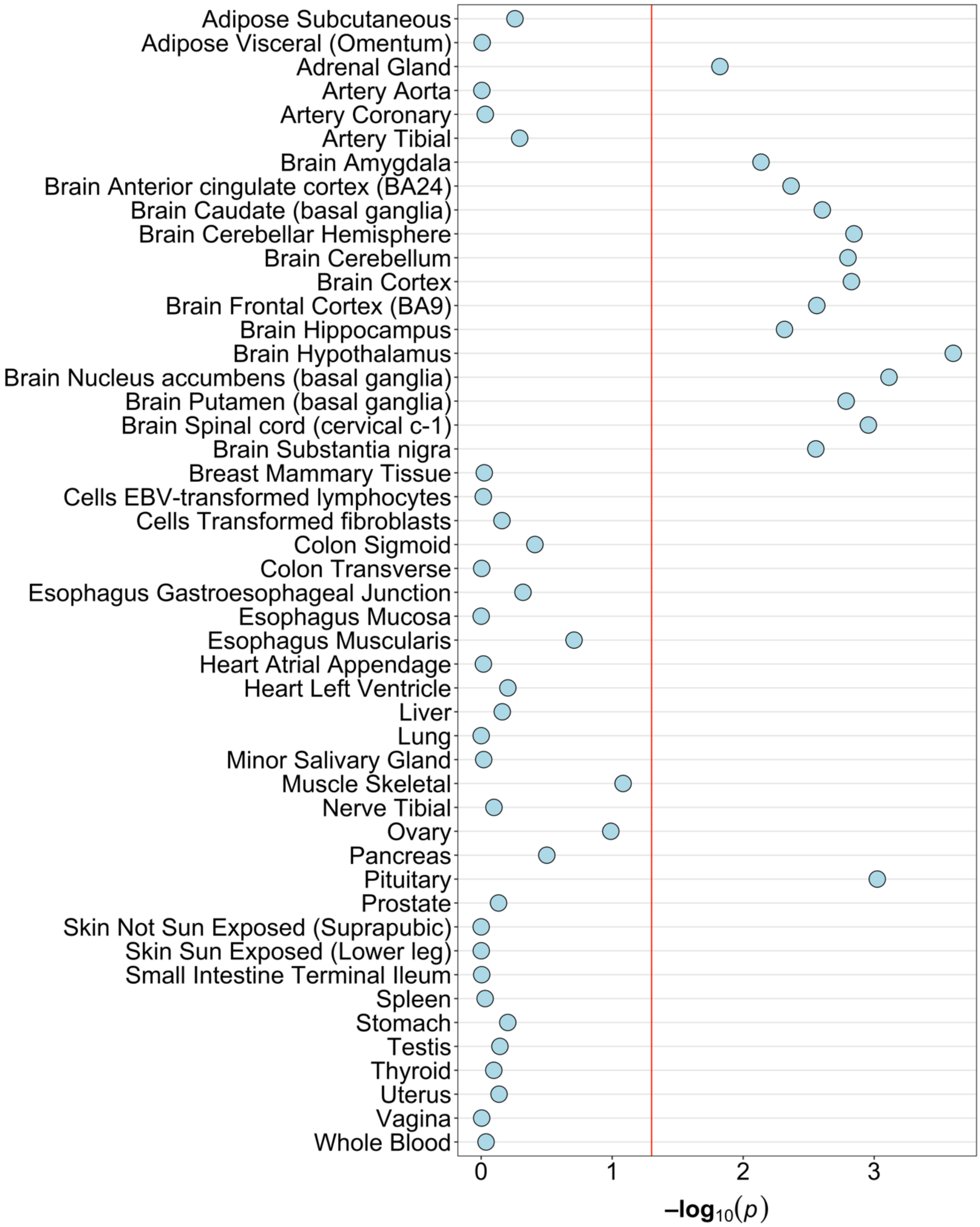

**Supplementary Figure 9 | Tissue enrichment for AAM GWAS associations.** Linkage-disequilibrium score regression to specifically expressed genes (LDSC-SEG) was used to test for enrichment of the EUR-MA GWAS data for genes specifically expressed across the different GTEx tissues. LDSC-SEG P-values represent a one-sided test that the coefficient is greater than zero. Extended data are shown in Supplementary Table 17.

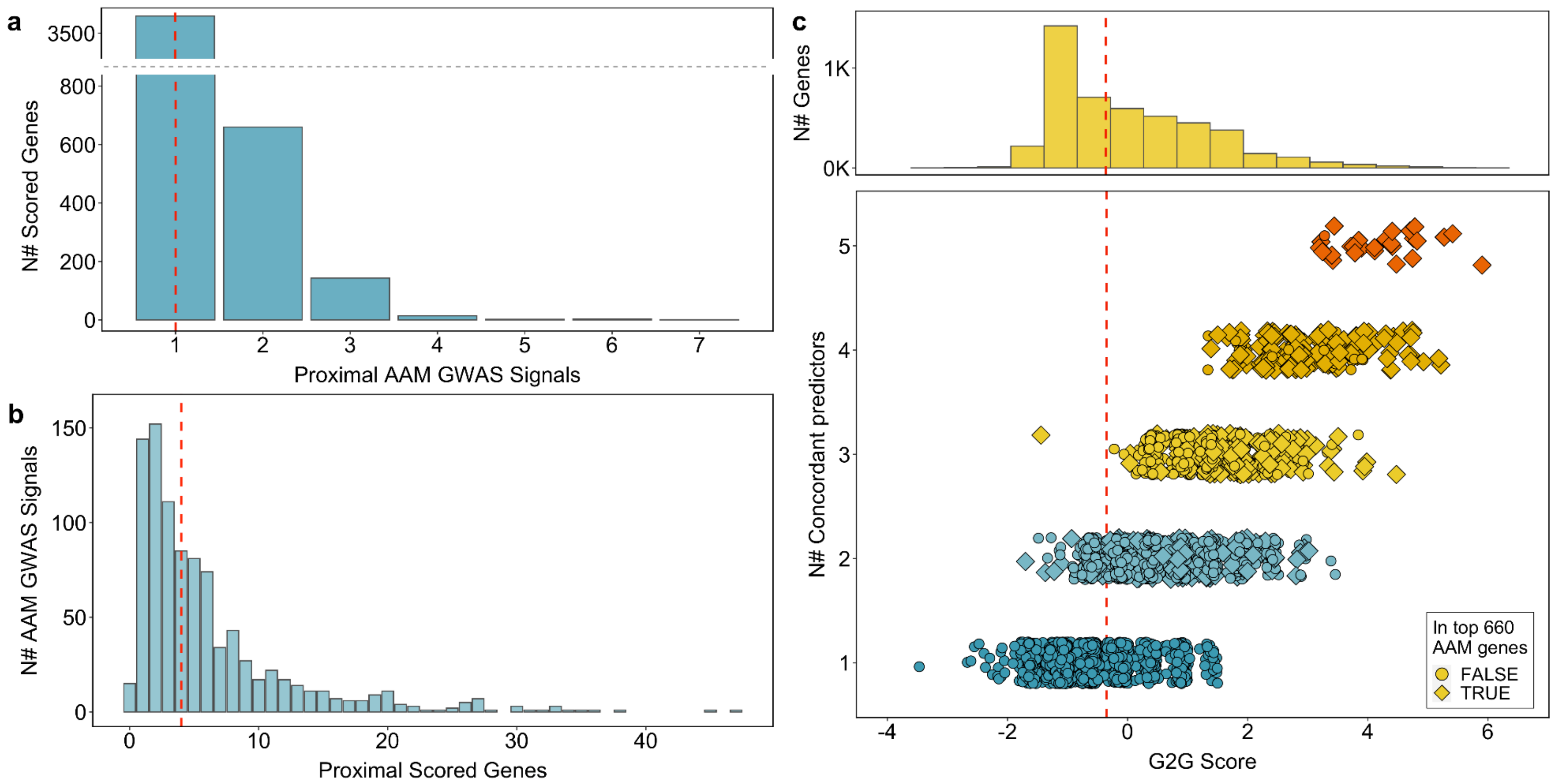

**Supplementary Figure 10 | Distribution of AAM GWAS signals and genes, as identified by GWAS to Genes (G2G).** (a) Based on the EUR-MA GWAS, 4,689 genes were implicated by G2G as potential regulators of AAM. Most were proximal to (within 500kb) only 1 GWAS signal (median signals per gene: 1; range: 1 to 7). (b) Conversely, each GWAS signal was annotated to be proximal (within 500kb) to (median) 4 genes (range: 0 to 47). (c) The 4,689 genes had a median G2G score -0.35 (range -3.52 to 5.90) derived from a maximum of 6 predictors (up to 5 observed). The 660 high confidence AAM genes were defined as the top-scoring gene at each independent AAM signal with at least 2 concordant predictors.  Extended data are shown in Supplementary Table 13.

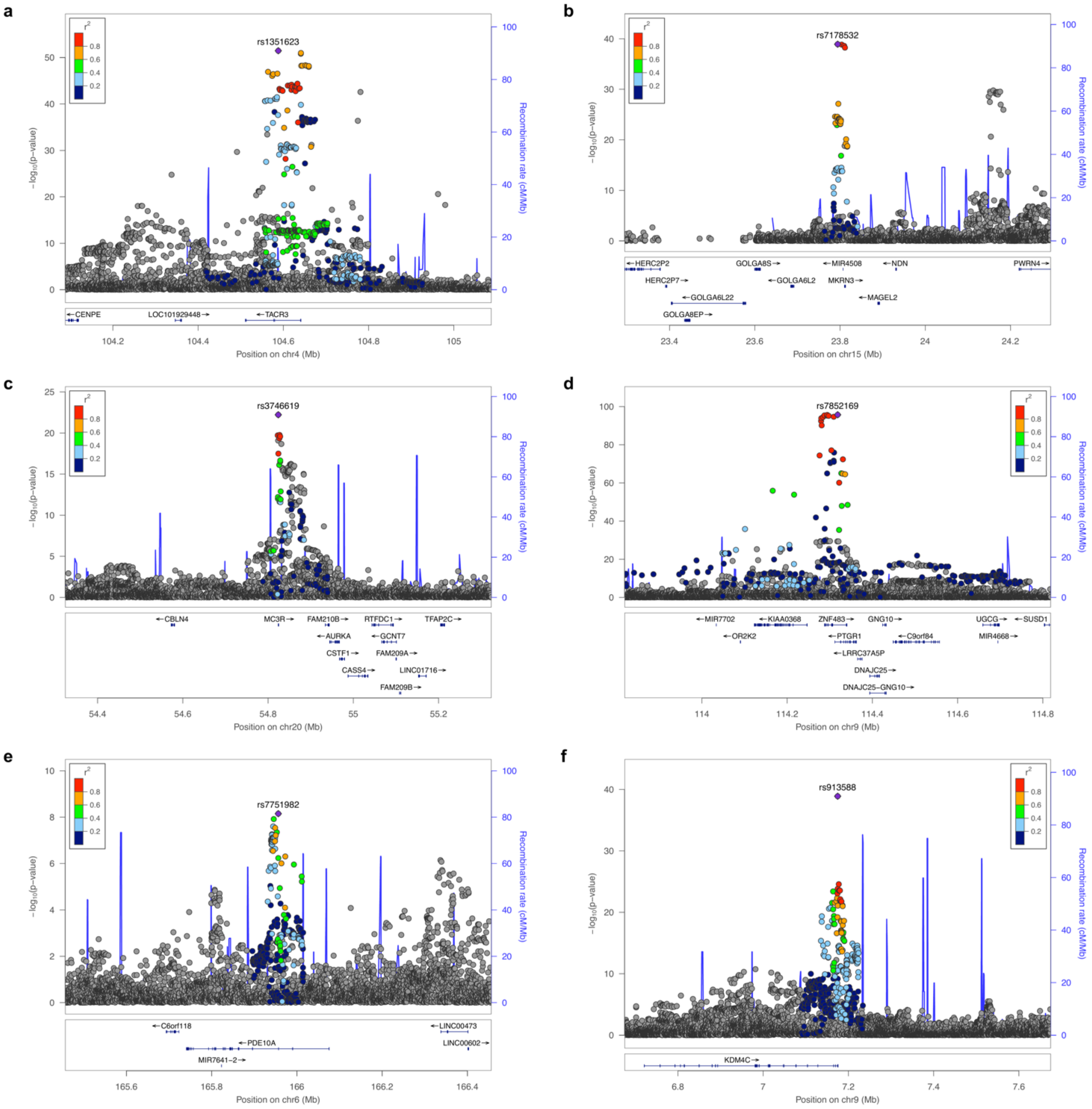

**Supplementary Figure 11 | GWAS loci proximal to the exome-wide significant genes.** Associations in the EUR-MA surrounding the 6 genes (±500kb) identified via exome-wide associations with AAM; (a) TACR3, (b) MKRN3, (c) MC3R, (d) ZNF483, (e) PDE10A, and (f) KDM4C.

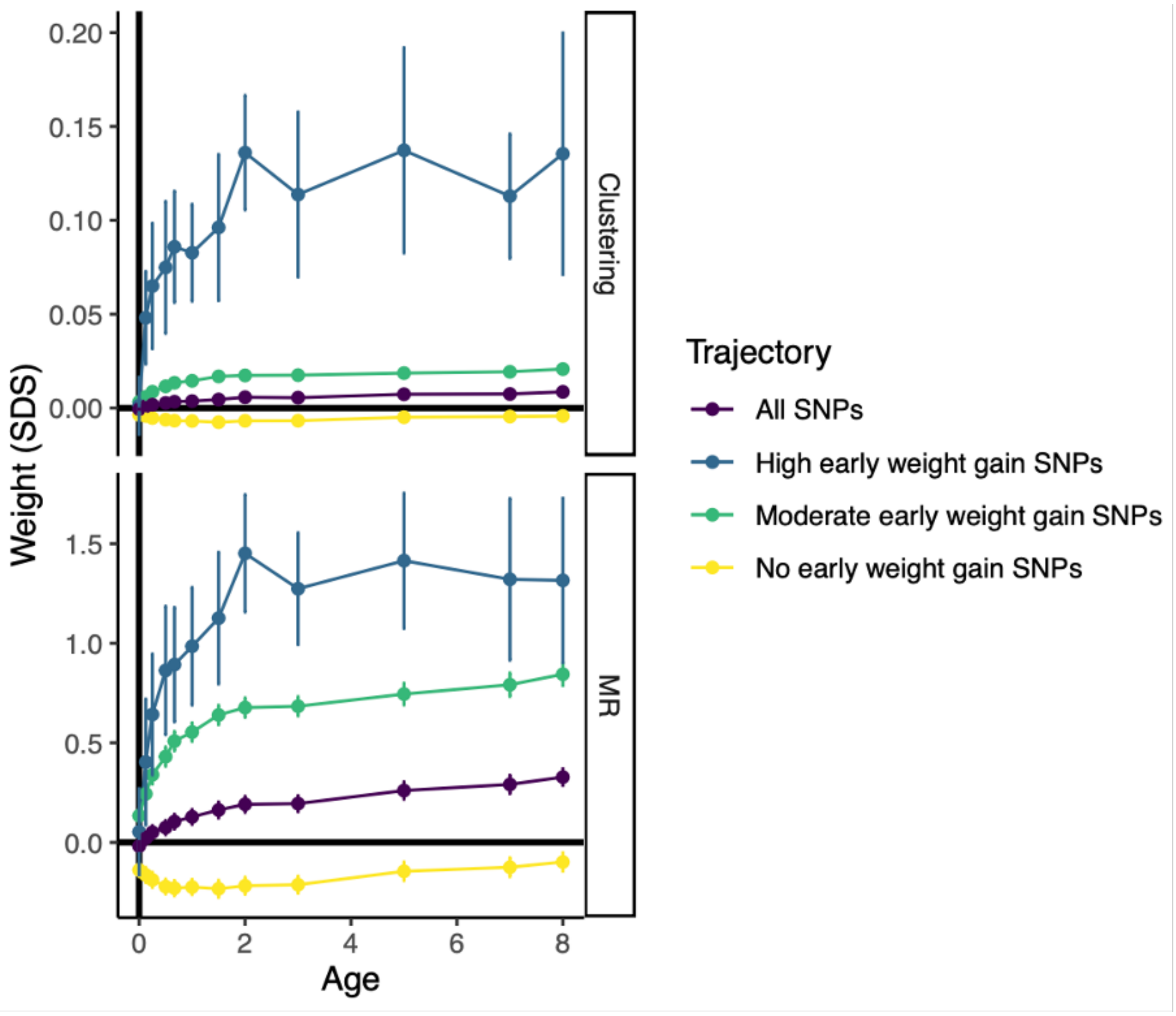

**Supplementary Figure 12 | Mean childhood body weight trajectory for each of the three AAM SNP clusters (aligned to the AAM-decreasing allele).** Mendelian randomisation (MR) estimates for all 1080 AAM signals, and the three AAM SNP clusters as separate exposures, on childhood body weight at 12 time-points and also adult BMI. Extended data are shown in Supplementary Tables 21 and 22.

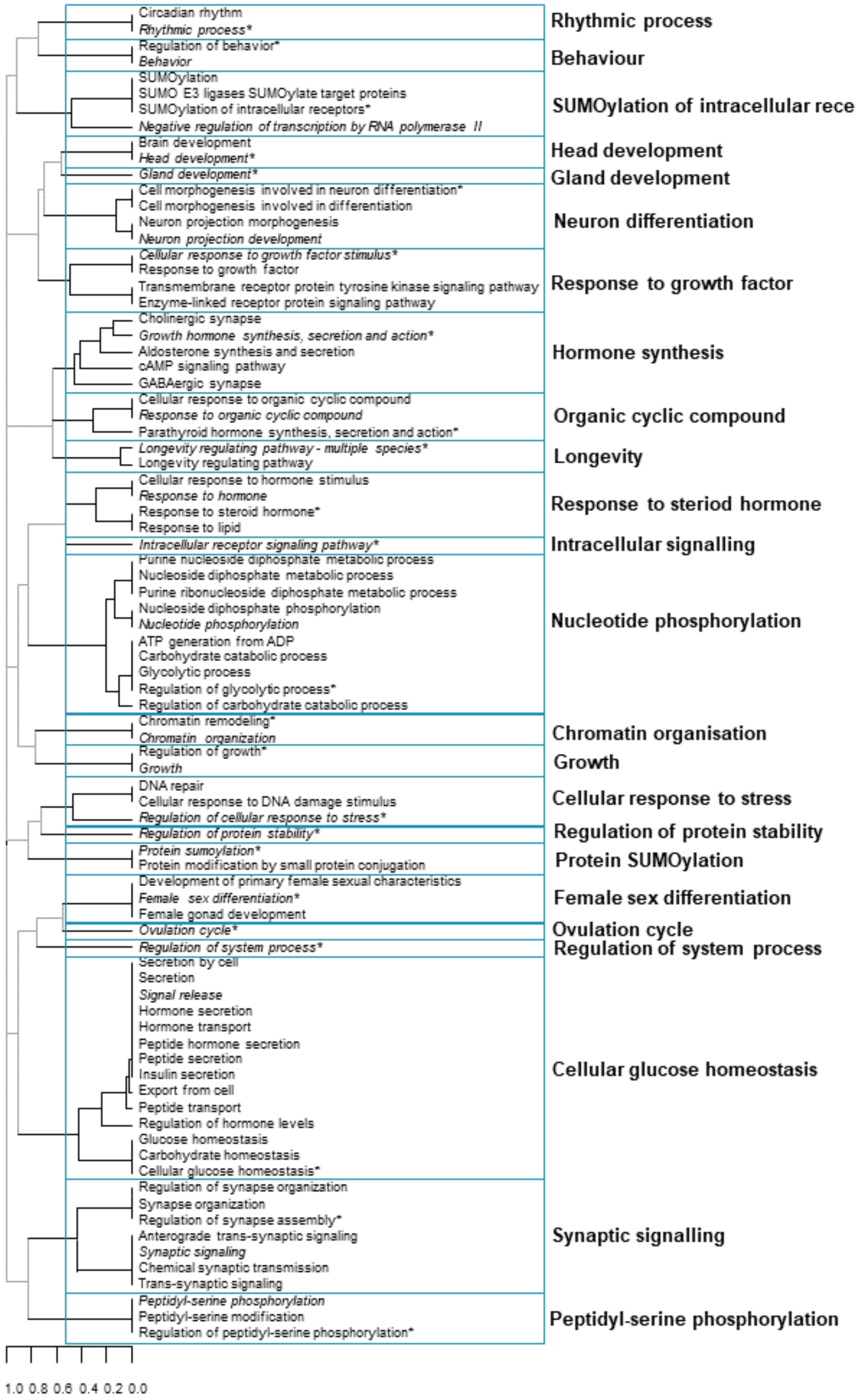

**Supplementary Figure 13 | Dendrogram showing the clustering of pathways enriched in all 660 high confidence AAM genes.** In each cluster the most significant pathway is in italics and the mostly strongly enriched pathway is marked by an asterisk. Extended data are shown in Supplementary Table 23.

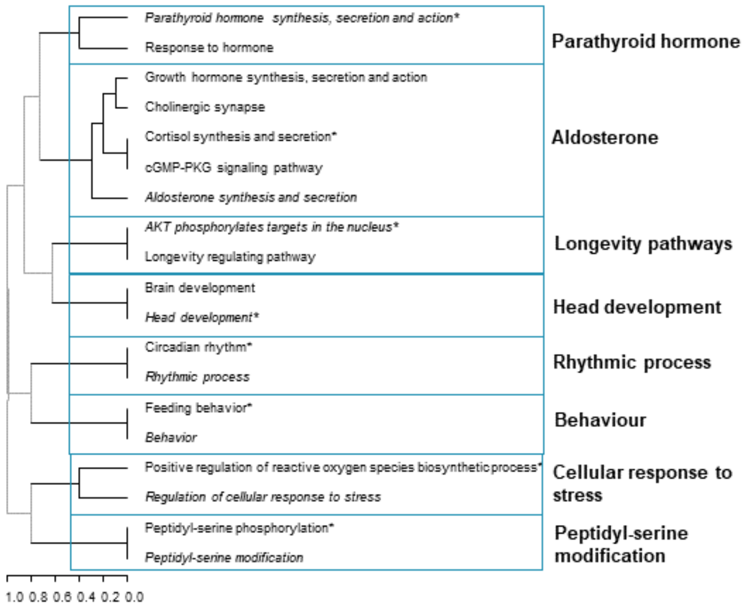

**Supplementary Figure 14 | Dendrogram showing the clustering of pathways enriched for AAM genes in the "early weight gain" trajectory.** In each cluster the most significant pathway is in italics and the mostly strongly enriched pathway is marked with an asterisk. Extended data are shown in Supplementary Table 25.

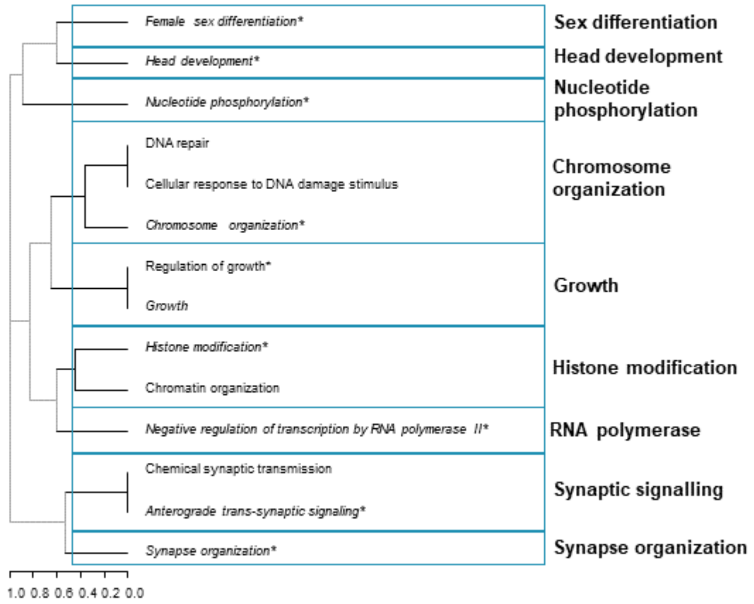

**Supplementary Figure 15 | Dendrogram showing the clustering of pathways enriched for AAM genes in the "no early weight gain" trajectory.** In each cluster the most significant pathway is in italics and the mostly strongly enriched pathway is marked with an asterisk. Extended data are shown in Supplementary Table 25.

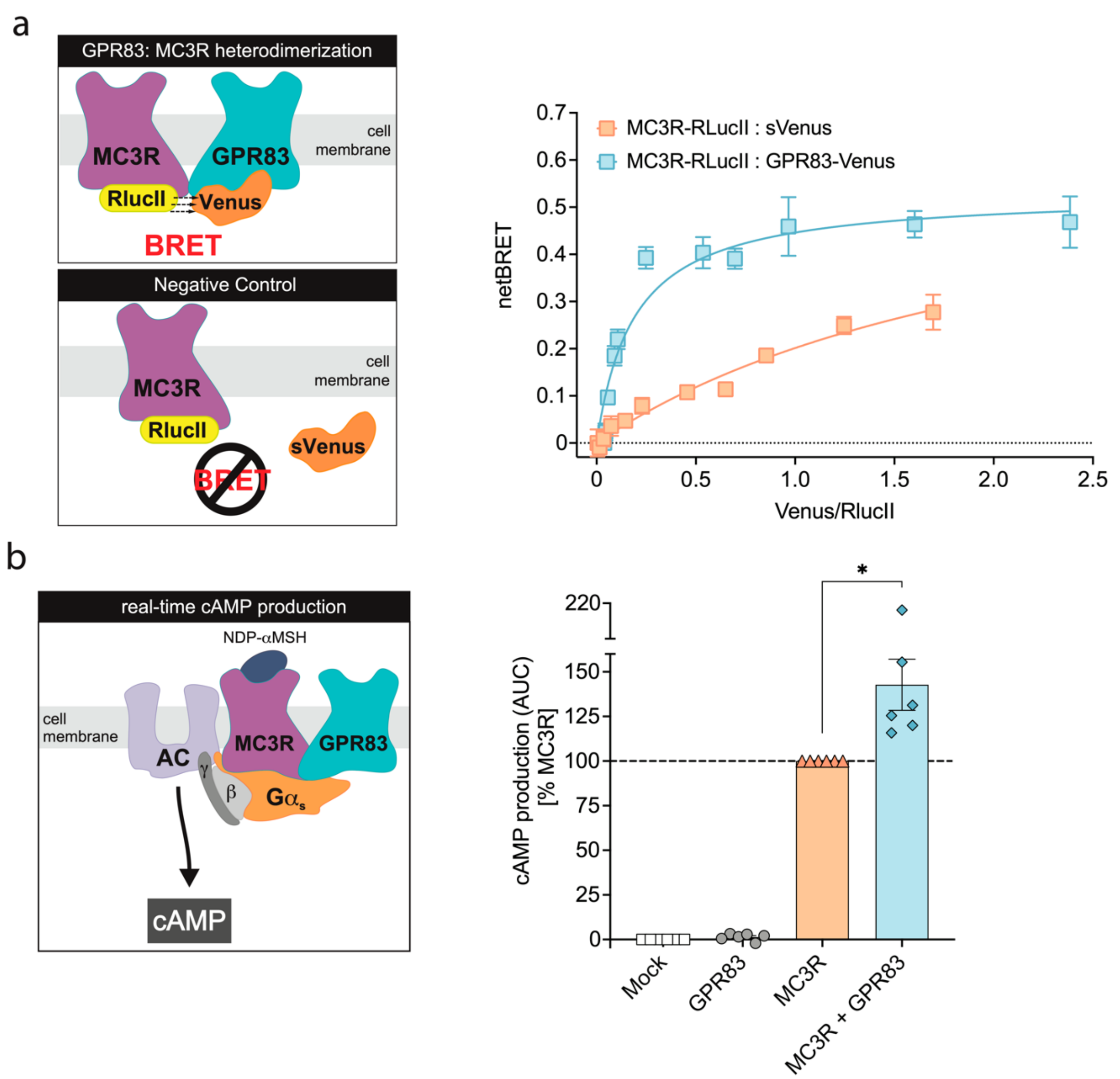

**Supplementary Figure 16 | GPR83-MC3R heterodimerization modulates canonical MC3R-cAMP signalling pathway.** (a) BRET saturation curve from HEK293 cells co-transfected with a constant amount of MC3R-RlucII donor construct and increasing amounts of the GPR83-Venus acceptor construct, indicating a specific and saturable GPR83-MC3R interaction. The selectivity of the observed signal was further supported by the observation that co-expression of MC3R-RlucII with a soluble acceptor (sVenus) led to lower BRET signals that progressed linearly over the same range of acceptor/donor ratios. (b) NDP-αMSH-stimulated cAMP production area under the curve calculation from HEK293 cells transfected with MC3R or co-transfected with both MC3R and GPR83. The time-resolved data are shown in Figure 6. Data are expressed as a percentage of maximal control response [% MC3R] and plotted as mean ± standard error from 6 independent experiments. Statistical significance was determined by unpaired t-test with Welch's correction; * $P < 0.05$. Extended data are shown in Supplementary Tables 29-31.

#### Consortia membership

##### The China Kadoorie Biobank Collaborative Group

**International Steering Committee:** Junshi Chen, Zhengming Chen (PI), Robert Clarke, Rory Collins, Yu Guo, Liming Li (PI), Chen Wang, Jun Lv, Richard Peto, Robin Walters.

**International Co-ordinating Centre, Oxford:** Daniel Avery, Derrick Bennett, Ruth Boxall, Sushila Burgess, Ka Hung Chan, Yumei Chang, Yiping Chen, Zhengming Chen, Johnathan Clarke; Robert Clarke, Huaidong Du, Ahmed Edris, Hannah Fry, Simon Gilbert, Mike Hill, Michael Holmes, Pek Kei Im, Andri Iona, Maria Kakkoura, Christiana Kartsonaki, Hubert Lam, Kuang Lin, Mohsen Mazidi, Iona Millwood, Sam Morris, Qunhua Nie, Alfred Pozarickij, Paul Ryder, Saredo Said, Dan Schmidt, Paul Sherliker, Becky Stevens, Iain Turnbull, Robin Walters, Lin Wang, Neil Wright, Ling Yang, Xiaoming Yang, Pang Yao.

**National Co-ordinating Centre, Beijing:** Yu Guo, Xiao Han, Can Hou, Qingmei Xia, Chao Liu, Jun Lv, Pei Pei, Canqing Yu.

**Regional Co-ordinating Centres:**

**Gansu:** Gansu Provincial CDC – Caixia Dong, Pengfei Ge, Xiaolan Ren. Maiji CDC – Zhongxiao Li, Enke Mao, Tao Wang, Hui Zhang, Xi Zhang. **Haikou:** Hainan Provincial CDC – Jinyan Chen, Ximin Hu, Xiaohuan Wang. Meilan CDC – Zhendong Guo, Huimei Li, Yilei Li, Min Weng, Shukuan Wu. **Harbin:** Heilongjiang Provincial CDC – Shichun Yan, Mingyuan Zou, Xue Zhou. Nangang CDC – Ziyan Guo, Quan Kang, Yanjie Li, Bo Yu, Qinai Xu. **Henan:** Henan Provincial CDC – Liang Chang, Lei Fan, Shixian Feng, Ding Zhang, Gang Zhou. Huixian CDC – Yulian Gao, Tianyou He, Pan He, Chen Hu, Huarong Sun, Xukui Zhang. **Hunan:** Hunan Provincial CDC – Biyun Chen, Zhongxi Fu, Yuelong Huang, Huilin Liu, Qiaohua Xu, Li Yin. Liuyang CDC – Huajun Long, Xin Xu, Hao Zhang, Libo Zhang. **Liuzhou:** Guangxi Provincial CDC – Naying Chen, Duo Liu, Zhenzhu Tang. Liuzhou CDC – Ningyu Chen, Qilian Jiang, Jian Lan, Mingqiang Li, Yun Liu, Fanwen Meng, Jinhuai Meng, Rong Pan, Yulu Qin, Ping Wang, Sisi Wang, Liuping Wei, Liyuan Zhou. **Qingdao:** Qingdao CDC – Liang Cheng, Ranran Du, Ruqin Gao, Feifei Li, Shanpeng Li, Yongmei Liu, Feng Ning, Zengchang Pang, Xiaohui Sun, Xiaocao Tian, Shaojie Wang, Yaoming Zhai, Hua Zhang. Licang CDC – Wei Hou, Silu Lv, Junzheng Wang. **Sichuan:** Sichuan Provincial CDC – Xiaofang Chen, Xianping Wu, Ningmei Zhang, Weiwei Zhou. Pengzhou CDC – Xiaofang Chen, Jianguo Li, Jiaqiu Liu, Guojin Luo, Qiang Sun, Xunfu Zhong. **Suzhou:** Jiangsu Provincial CDC – Jian Su, Ran Tao, Ming Wu, Jie Yang, Jinyi Zhou, Yonglin Zhou. Suzhou CDC – Yihe Hu, Yujie Hua, Jianrong Jin Fang Liu, Jingchao Liu, Yan Lu, Liangcai Ma, Aiyu Tang, Jun Zhang. **Zhejiang**: Zhejiang Provincial CDC – Weiwei Gong, Ruying Hu, Hao Wang, Meng Wang, Min Yu. Tongxiang CDC – Lingli Chen, Qijun Gu, Dongxia Pan, Chunmei Wang, Kaixu Xie, Xiaoyi Zhang.

##### Lifelines Cohort Study

Behrooz Z Alizadeh[1], H Marike Boezen[1], Lude Franke[2], Pim van der Harst[3], Gerjan Navis[4], Marianne Rots[5], Harold Snieder[1], Morris Swertz[2], Bruce HR Wolffenbuttel[6], and Cisca Wijmenga[2]

1. Department of Epidemiology, University of Groningen, University Medical Center Groningen, The Netherlands
2. Department of Genetics, University of Groningen, University Medical Center Groningen, The Netherlands
3. Department of Cardiology, University of Groningen, University Medical Center Groningen, The Netherlands
4. Department of Internal Medicine, Division of Nephrology, University of Groningen, University Medical Center

Groningen, The Netherlands 5. Department of Pathology and Medical Biology, University of Groningen, University Medical Center Groningen, The Netherlands 6. Department of Endocrinology, University of Groningen, University Medical Center Groningen, The Netherlands

#### Danish Blood Donors Study

Karina Banasik[1], Jakob Bay[2], Jens Kjærgaard Boldsen[3], Thorsten Brodersen[2], Søren Brunak[1], Kristoffer Burgdorf[1], Mona Ameri Chalmer[4], Maria Didriksen[5], Khoa Manh Dinh[3], Joseph Dowsett[5], Christian Erikstrup[3,6], Bjarke Feenstra[5,7], Frank Geller[5,7], Daniel Gudbjartsson[8], Thomas Folkmann Hansen[4], Lotte Hindhede[3], Henrik Hjalgrim[9,7], Rikke Louise Jacobsen[5], Gregor Jemec[10], Bitten Aagaard Jensen[11], Katrine Kaspersen[3], Bertram Dalskov Kjerulff[3], Lisette Kogelman[4], Margit Anita Hørup Larsen[5], Ioannis Louloudis[1], Agnete Lundgaard[1], Susan Mikkelsen[3], Christina Mikkelsen[5], Ioanna Nissen[5], Mette Nyegaard[12], Sisse Rye Ostrowski[5,13], Ole Birger Pedersen[2,13], Alexander Pil Henriksen[1], Palle Duun Rohde[12], Klaus Rostgaard[9,7], Michael Schwinn[5], Kari Stefansson[8], Hreinn Stefánsson[8], Erik Sørensen[5], Unnur Þorsteinsdóttir[8], Lise Wegner Thørner[5], Mie Topholm Bruun[14], Henrik Ullum[15], Thomas Werge[16,13], and David Westergaard[1]

1. Novo Nordisk Foundation Center for Protein Research, Faculty of Health and Medical Sciences, University of Copenhagen, Copenhagen, Denmark, 2. Department of Clinical Immunology, Zealand University Hospital, Køge, Denmark, 3. Department of Clinical Immunology, Aarhus University Hospital, Aarhus, Denmark, 4. Danish Headache Center, Department of Neurology, Copenhagen University Hospital, Rigshospitalet-Glostrup, Copenhagen, Denmark, 5. Department of Clinical Immunology, Copenhagen University Hospital, Rigshospitalet, Copenhagen, Denmark, 6. Department of Clinical Medicine, Health, Aarhus University, Aarhus, Denmark, 7. Department of Epidemiology Research, Statens Serum Institut, Copenhagen, Denmark, 8. deCODE Genetics, Reykjavik, Iceland, 9. Danish Cancer Society Research Center, Copenhagen, Denmark, 10. Department of Dermatology, Zealand University hospital, Roskilde, Denmark, 11. Department of Clinical Immunology, Aalborg University Hospital, Aalborg, Denmark, 12. Department of Health Science and Technology, Faculty of Medicine, Aalborg Univeristy, Aalborg, Denmark, 13. Department of Clinical Medicine, Faculty of Health and Medical Sciences, University of Copenhagen, Copenhagen, Denmark, 14. Department of Clinical Immunology, Odense University Hospital, Odense, Denmark, 15. Statens Serum Institut, Copenhagen, Denmark, 16. Institute of Biological Psychiatry, Mental Health Centre, Sct. Hans, Copenhagen University Hospital, Roskilde, Denmark

#### The Ovarian Cancer Association Consortium

CM Phelan[1], KB Kuchenbaecker[2,3], JP Tyrer[4], SP Kar[4], K Lawrenson[5], SJ Winham[6], J Dennis[4], A Pirie[4], M Riggan[7], G Chornokur[8], MA Earp[9], PC Lyra, Jr.[8], JM. Lee[10], S Coetzee[10], J Beesley[11], L McGuffog[12], P Soucy[13], E Dicks[4], A Lee[12], D Barrowdale[12], J Lecarpentier[12], G Leslie[12], CM Aalfs[14], KKH Aben[15,16], M Adams[17], J Adlard[18], IL Andrulis[19], H Anton-Culver[20], N Antonenkova[21], AOCS study group[22], G Aravantinos[23], N Arnold[24], BK Arun[25], B Arver[26], J Azzollini[27], J Balmaña[28], SN Banerjee[29], L Barjhoux[30], RB Barkardottir[31,32], Y Bean[33], MW Beckmann[34], A BeeghlyFadiel[35], J Benitez[36], M Bermisheva[37], MQ Bernardini[38], MJ Birrer[39], L Bjorge[40,41], A Black[42], K Blankstein[43], MJ Blok[44], C Bodelon[42], N Bogdanova[45], A Bojesen[46], B Bonanni[47], Å Borg[48], AR Bradbury[49], JD Brenton[50], C Brewer[51], L Brinton[42], P Broberg[52], A Brooks-Wilson[53], F Bruinsma[54], J Brunet[55], B Buecher[56], R Butzow[57], SS Buys[58], T Caldes[59], MA Caligo[60], I Campbell[61,62], R Cannioto[63], ME Carney[64], T Cescon[43], SB Chan[65], J Chang-Claude[66,67], S Chanock[42], X Qing Chen[11], Y-E Chiew[68,69], J Chiquette[70], WK Chung[71], KBM Claes[72], T Conner[58], LS Cook[73], J Cook[74], DW Cramer[75], JM Cunningham[76], AA D'Aloisio[77], MB Daly[78], F Damiola[30], S Dina Damirovna[79], A Dansonka-Mieszkowska[80], F Dao[81], R Davidson[82], A DeFazio[68,69], C Delnatte[83], KF Doheny[17], O Diez[84,85], Y Chun Ding[86], J Anne Doherty[87], SM Domchek[49], CM Dorfling[88], T Dörk[89], L Dossus[90], M Duran[91], M Dürst[92], B Dworniczak[93], D Eccles[94], T Edwards[35], R Eeles[95], U Eilber[66], B Ejlertsen[96], AB Ekici[97], S Ellis[12], M Elvira[79], Study EMBRACE[22], KH Eng[98], C Engel[99], DG Evans[100], PA Fasching[101,34], S Ferguson[38], S Fert Ferrer[102], JM Flanagan[103], ZC Fogarty[6], RT Fortner[66], F Fostira[104], WD Foulkes[105], G Fountzilas[106], BL Fridley[107], TM Friebel[108], E Friedman[109], D Frost[12], PA Ganz[110], J Garber[111], MJ García[36], V GarciaBarberan[59], A Gehrig[112], GEMO Study Collaborators[22], A GentryMaharaj[113], A-M Gerdes[114], GG Giles[54,115,116], R Glasspool[117], G Glendon[118], AK Godwin[119], DE Goldgar[120], T Goranova[50], M Gore[121], MH Greene[122], J Gronwald[123], S Gruber[124], E Hahnen[125], CA Haiman[126],

N Håkansson[127], U Hamann[128], TVO Hansen[129], PA Harrington[4], HR Harris[127], J Hauke[125] HEBON Study[22], A Hein[34], A Henderson[130], MAT Hildebrandt[131], P Hillemanns[132], S Hodgson[133], CK Høgdall[134], E Høgdall[135,136], FBL Hogervorst[137], H Holland[11], MJ Hooning[138], K Hosking[4], R-Y Huang[139], PJ Hulick[140], J Hung[68,69], DJ Hunter[141], DG Huntsman[142], T Huzarski[123], EN Imyanitov[143], C Isaacs[144], ES Iversen[145], L Izatt[146], A Izquierdo[55], A Jakubowska[123], P James[147], R Janavicius[148,149], M Jernetz[150], A Jensen[135], U Birk Jensen[151], EM. John[152], S Johnatty[11], ME Jones[153], P Kannisto[150], BY Karlan[5], A Karnezis[142], K Kast[154], KconFab Investigators[22], CJ Kennedy[68,69], E Khusnutdinova[37], LA Kiemeney[15], JI Kiiski[155], S-W Kim[156], SK Kjaer[134,135], M Köbel[157], RK Kopperud[40,41], TA Kruse[158], J Kupryjanczyk[80], A Kwong[159,160,161], Y Laitman[109], D Lambrechts[162,163], N Larrañaga[164,165], MC Larson[6], C Lazaro[166] ND Le[167] L Le Marchand[168], J Won Lee[169], SB Lele[170], A Leminen[155], D Leroux[171], J Lester[5], F Lesueur[172], DA Levine[81], D Liang[173], C Liebrich[174], J Lilyquist[175], L Lipworth[176], J Lissowska[177], KH Lu[178], J Lubiński[123], C Luccarini[4], L Lundvall[179], PL Mai[122], G Mendoza-Fandiño[8], S Manoukian[27], LFAG Massuger[15], T May[38], S Mazoyer[180], JN McAlpine[181], V McGuire[182], JR McLaughlin[183], I McNeish[184], H MeijersHeijboer[185], A Meindl[186], U Menon[113], AR Mensenkamp[187], MA Merritt[188], RL Milne[54,115], G Mitchell[147,189], F Modugno[190,191,192], J Moes-Sosnowska[80], M Moffitt[193,194], M Montagna[195], KB Moysich[63], AM Mulligan[196,197], J Musinsky[198], KL Nathanson[49], L Nedergaard[199], RB Ness[200], SL Neuhausen[86], HNevanlinna[155], D Niederacher[201], RL Nussbaum[202], K Odunsi[170], E Olah[203], OI Olopade[204], H Jernström[205], C Olswold[175], DM O'Malley[206], K Ong[207], NC Onland-Moret[208], OPAL study group[22], N Orr[209], S Orsulic[5], A Osorio[36], D Palli[210], L Papi[211], T-W Park-Simon[132], J Paul[212], CL Pearce[213,126], I Søkilde Pedersen[214], PHM Peeters[208], B Peissel[27], A Peixoto[215], T Pejovic[193,194], LM Pelttari[155], JB Permuth[8], P Peterlongo[216], L Pezzani[27], G Pfeiler[217], K-A Phillips[189,115,218], M Piedmonte[219], MC Pike[126,220], AM Piskorz[50], SR Poblete[170], T Pocza[203], EM Poole[221], B Poppe[72], ME Porteous[222], F Prieur[223], D Prokofyeva[79], E Pugh[17], M Angel Pujana[224], P Pujol[225], P Radice[226], J Rantala[227], C Rappaport-Fuerhauser[217], G Rennert[228], K Rhiem[125], P Rice[43], A Richardson[229], M Robson[230], GC Rodriguez[231], C Rodríguez-Antona[232], J Romm[17], MA Rookus[233], M Anne Rossing[234], JH Rothstein[182], A Rudolph[66], IB Runnebaum[92], HB Salvesen[40,41], DP Sandler[235], MJ Schoemaker[153], L Senter[236], VW Setiawan[126], G Severi[237,238,239,240], P Sharma[241], T Shelford[17], N Siddiqui[242], LE Side[243], W Sieh[182], CF Singer[217], H Sobol[244], H Song[4], MC Southey[245], AB Spurdle[11], Z Stadler[230], D Steinemann[246], D Stoppa-Lyonnet[56], LE Sucheston-Campbell[247], G Sukiennicki[123], R Sutphen[248], C Sutter[249], AJ Swerdlow[153,250], CI Szabo[251], L Szafron[80], YY Tan[217], JA Taylor[235], M-K Tea[217], MR Teixeira[252], S-H Teo[253,254], KL Terry[75,255], PJ Thompson[256], LCV Thomsen[40,41], DL Thull[257], L Tihomirova[258] AV Tinker[259], M Tischkowitz[105,260], S Tognazzo[195], A Ewart Toland[261], A Tone[38], B Trabert[42], RC Travis[262], A Trichopoulou[263,264], N Tung[265], SS. Tworoger[221,255], AM van Altena[266], D Van Den Berg[126], AH van der Hout[267], RB van der Luijt[268], M Van Heetvelde[72], E Van Nieuwenhuysen[269] EJ van Rensburg[88], A Vanderstichele[269], R Varon-Mateeva[270], V Ana[271,272], D Velez Edwards[273], I Vergote[269], RA Vierkant[6], J Vijai[198], A Vratimos[104], L Walker[274], C Walsh[5], D Wand[275], S Wang-Gohrke[276], B Wappenschmidt[125], PM Webb[277], CR Weinberg[278], JN Weitzel[279], N Wentzensen[42], AS Whittemore[182,280], JT Wijnen[281], LR Wilkens[168], A Wolk[127], M Woo[142], X Wu[131], AH Wu[126], H Yang[42], D Yannoukakos[104], A Ziogas[282], KK Zorn[257], SA Narod[283], DF Easton[3,4], CI Amos[284], JM Schildkraut[28,5], SJ Ramus[286,287], L Ottini[288], MT Goodman[256,289], SK Park[290,291,292], LE Kelemen[293], HA Risch[294], M Thomassen[158], K Offit[198], J Simard[13], R Katharina Schmutzler[125], D Hazelett[295], AN Monteiro[8], FJ Couch[76], A Berchuck[7], G Chenevix-Trench[11], EL Goode[9], TA. Sellers[8], SA Gayther[10], AC Antoniou[3,4], and PDP Pharoah[3,4]

1. Departments of Cancer Epidemiology and Gynecologic Oncology, Moffitt Cancer Center, Tampa, FL, USA 2. Wellcome Trust Sanger Institute, Wellcome Genome Campus, Hinxton, Cambridgeshire CB10 1SA, UK 3. Centre for Cancer Genetic Epidemiology, Department of Public Health and Primary Care, University of Cambridge, Strangeways Research Laboratory, Worts Causeway, Cambridge, UK 4. Department of Oncology, Centre for Cancer Genetic Epidemiology, University of Cambridge, Strangeways Research Laboratory, Cambridge, UK 5. Women's Cancer Program at the Samuel Oschin Comprehensive Cancer Institute, CedarsSinai Medical Center, 8700 Beverly Boulevard, Suite 290W, Los Angeles, CA, USA 6. Department of Health Science Research, Division of Biomedical Statistics and Informatics, Mayo Clinic, Rochester, Minnesota, USA 7. Department of

Obstetrics and Gynecology, Duke University Medical Center, Durham, North Carolina, USA 8. Department of Cancer Epidemiology, Moffitt Cancer Center, Tampa, FL, USA 9. Department of Health Science Research, Division of Epidemiology, Mayo Clinic, Rochester, Minnesota, USA 10. Center for Bioinformatics and Functional Genomics, Samuel Oschin Comprehensive Cancer Institute, Cedars-Sinai Medical Center, Los Angeles, California, USA 11. Department of Genetics and Computational Biology, QIMR Berghofer Medical Research Institute, Brisbane, QLD, Australia 12. Centre for Cancer Genetic Epidemiology, Department of Public Health and Primary Care, University of Cambridge, Cambridge, UK 13. Genomics Center, Centre Hospitalier Universitaire de Québec Research Center and Laval University, 2705 Laurier Boulevard, Quebec City (Quebec), Canada 14. Department of Clinical Genetics, Academic Medical Center, Amsterdam, The Netherlands 15. Radboud university medical center, Radboud Institute for Health Sciences, Department for Health Evidence, Nijmegen, The Netherlands 16. Netherlands Comprehensive Cancer Organisation, Utrecht, The Netherlands 17. Center for Inherited Disease Research, Institute of Genetic Medicine, Johns Hopkins University School of Medicine, Baltimore, MD 21224 18. Yorkshire Regional Genetics Service, Chapel Allerton Hospital, Leeds, UK 19. Lunenfeld-Tanenbaum Research Institute, Mount Sinai Hospital, Toronto, Ontario M5G 1X5, Departments of Molecular Genetics and Laboratory Medicine and Pathobiology, University of Toronto, Ontario, Canada 20. Department of Epidemiology, Director of Genetic Epidemiology Research Institute, UCI Center for Cancer Genetics Research & Prevention, School of Medicine, University of California Irvine, Irvine, California, USA 21. N.N. Alexandrov National Cancer Centre of Belarus, Minsk, Belarus 22. A list of members and affiliations appears in the Supplementary note 23. “Agii Anargiri” Cancer Hospital, Athens, Greece 24. Department of Gynaecology and Obstetrics, University Hospital of Schleswig-Holstein, Campus Kiel, Christian-Albrechts University Kiel, Germany 25. Department of Breast Medical Oncology and Clinical Cancer Genetics Program, University Of Texas MD Andersson Cancer Center, 1515 Pressler Street, CBP 5, Houston, TX, USA 26. Department of Oncology and Pathology, Karolinska University Hospital and Karolinska Institutet, Stockholm, Sweden 27. Unit of Medical Genetics, Department of Preventive and Predictive Medicine, Fondazione IRCCS (Istituto Di Ricovero e Cura a Carattere Scientifico) Istituto Nazionale Tumori (INT), Via Giacomo Venezian 1, 20133 Milan, Italy 28. Department of Medical Oncology, University Hospital, Vall d’Hebron, Barcelona, Spain 29. Gynaecology Unit, The Royal Marsden Hospital, London, UK 30. Bâtiment Cheney D, Centre Léon Bérard, 28 rue Laënnec, Lyon, France 31. Laboratory of Cell Biology, Department of Pathology, hus 9, Landspitali-LSH v/Hringbraut, 101 Reykjavik, Iceland 32. BMC (Biomedical Centre), Faculty of Medicine, University of Iceland, Vatnsmyrarvegi 16, 101 Reykjavik, Iceland 33. Department of Gynecologic Oncology, Oregon Health & Science University, Portland, OR, USA; Knight Cancer Institute, Portland, OR, USA 34. Universtiy Hospital Erlangen, Department of Gynecology and Obstetrics, FriedrichAlexander-University Erlangen-Nuremberg, Comprehensive Cancer Center Erlangen Nuremberg, Universitaetsstrasse 21-23, 91054 Erlangen, Germany 35. Division of Epidemiology, Department of Medicine, Vanderbilt Epidemiology Center, Institute for Medicine and Public Health, Vanderbilt University Medical Center, VanderbiltIngram Cancer Center, Nashville, TN, USA 36. Human Genetics Group, Spanish National Cancer Centre (CNIO), and Biomedical Network on Rare Diseases (CIBERER), Madrid, Spain 37. Institute of Biochemistry and Genetics, Ufa Science Center, Russian Academy of Sciences, Ufa, Bashkortostan, Russia 38. Division of Gynecologic Oncology, Princess Margaret Hospital, University Health Network, Toronto, Ontario, Canada 39. Department of Medicine, Massachusetts General Hospital, Boston, USA 40. Department of Gynecology and Obstetrics, Haukeland University Hospital, Bergen, Norway 41. Centre for Cancer Biomarkers, Department of Clinical Science, University of Bergen, Bergen, Norway 42. Division of Cancer Epidemiology and Genetics, National Cancer Institute, Bethesda, MD, USA 43. Clinical Cancer Genetics, for the City of Hope Clinical Cancer Genetics Community Research Network, Duarte, CA, USA 44. Department of Clinical Genetics, Maastricht University Medical Center, Maastricht, The Netherlands 45. Radiation Oncology Research Unit, Hannover Medical School, Hannover, Germany 46. Department of Clinical Genetics, Vejle Hospital, Vejle, Denmark 47. Division of Cancer Prevention and Genetics, Istituto Europeo di Oncologia (IEO), via Ripamonti 435, 20141 Milan, Italy 48. Department of Oncology, Clinical Sciences, Lund University and Skåne University Hospital, Lund, Sweden 49. Department of Medicine, Abramson Cancer Center, Perelman School of Medicine at The University of Pennsylvania, Philadelphia, PA, USA 50. Cancer Research UK (CRUK) Cambridge Institute, University of Cambridge 51. Department of Clinical Genetics, Royal Devon and Exeter Hospital, Exeter, UK 52. Department of Cancer Epidemiology, University Hospital, Lund, Lund University, Lund, Sweden 53. Canada’s Michael Smith Genome Sciences Centre, BC Cancer Agency, Vancouver, BC, Canada 54. Cancer Epidemiology Centre, Cancer Council Victoria, Melbourne, VIC, Australia 55. Genetic Counseling Unit, Hereditary Cancer Program, IDIBGI (Institut d’Investigació Biomèdica de Girona), Catalan Institute of Oncology. Av. França s/n. 1707 Girona, Spain 56. Service de Génétique Oncologique, Institut Curie, 26, rue d’Ulm, Paris Cedex 05, France 57. Department of Pathology, University of Helsinki and Helsinki University Hospital, Helsinki, Finland 58. Department of Medicine, Huntsman Cancer Institute, 2000 Circle of Hope, Salt Lake City, UT 84112, USA 59. Molecular Oncology Laboratory, Hospital Clinico San Carlos, IdISSC (Instituto de Investigación Sanitaria del Hospital Clínico San Carlos), Martin Lagos s/n, Madrid, Spain 60. Section of Genetic Oncology, Dept. of Laboratory Medicine, University and University Hospital of Pisa, Pisa Italy 61. Cancer Genetics Laboratory, Research Division, Peter MacCallum Cancer Centre, Melbourne, VIC, Australia 62. Department of Pathology, University of Melbourne, Parkville, VIC, Australia 63. Cancer Pathology & Prevention, Division of Cancer Prevention and Population Sciences, Roswell Park Cancer Institute, Buffalo, NY 64. Department of Obstetrics and Gynecology, John A. Burns School of Medicine, University of Hawaii, Honolulu, Hawaii, US 65. University of California, San Francisco, 1600 Divisadero Street, C415, San Francisco, CA 94143 - 1714, USA 66. Division of Cancer Epidemiology, German Cancer Research Center (DKFZ), Heidelberg, Germany 67. University Cancer Center Hamburg (UCCH), University Medical Center HamburgEppendorf, Hamburg, Germany 68. Centre for Cancer Research, The Westmead Institute for Medical Research, The University of

Sydney, Sydney, New South Wales, Australia 69. Department of Gynaecological Oncology, Westmead Hospital, Sydney, New South Wales, Australia 70. Unité de Recherche en Santé des Populations, Centre des Maladies du Sein DeschênesFabia, Centre de Recherche FRSQ du Centre Hospitalier Affilié Universitaire de Québec, Québec, QC, Canada 71. Departments of Pediatrics and Medicine, Columbia University, New York, NY, USA 72. Center for Medical Genetics, Ghent University, Ghent, Belgium 73. Division of Epidemiology, Biostatistics and Preventative Medicine, Department of Internal Medicine, University of New Mexico, Albuquerque, New Mexico, USA 74. Sheffield Clinical Genetics Service, Sheffield Children's Hospital, Sheffield, UK 75. Obstetrics and Gynecology Epidemiology Center, Brigham and Women's Hospital and Harvard Medical School, Boston, MA, USA 76. Department of Laboratory Medicine and Pathology,Division of Experimental Pathology, Mayo Clinic, Rochester, MN, USA 77. Social & Scientific Systems, Inc. Durham, NC 27703, USA 78. Department of Clinical Genetics, Fox Chase Cancer Center, 333 Cottman Avenue, Philadelphia, PA 19111, USA 79. Department of Genetics and Fundamental Medicine, Bashkir State University, Ufa, Russia 80. Department of Pathology and Laboratory Diagnostics, the Maria Sklodowska-Curie Memorial Cancer Center and Institute of Oncology, Warsaw, Poland 81. Gynecology Service, Department of Surgery, Memorial Sloan Kettering Cancer Center, New York, NY, USA 82. Department of Clinical Genetics, South Glasgow University Hospitals, Glasgow, UK 83. Unité d'oncogénétique, ICO-Centre René Gauducheau, Boulevard Jacques Monod, 44805 Nantes Saint Herblain Cedex, France 84. Oncogenetics Group, Vall d'Hebron Institute of Oncology (VHIO), Barcelona, Spain 85. Clinical and Molecular Genetics Area, Vall d'Hebron University Hospital. Barcelona, Spain 86. Department of Population Sciences, Beckman Research Institute of City of Hope, Duarte, CA, USA 87. Department of Epidemiology, The Geisel School of Medicine - at Dartmouth, Hanover, New Hampshire, USA 88. Cancer Genetics Laboratory, Department of Genetics, University of Pretoria, Pretoria, South Africa 89. Gynaecology Research Unit, Hannover Medical School, Hannover, Germany 90. Nutrition and Metabolism Section, International Agency for Research on Cancer (IARCWHO), Lyon, France 91. Institute of Biology and Molecular Genetics, Universidad de Valladolid (IBGM-UVA), Valladolid, Spain 92. Department of Gynecology, Jena University Hospital - Friedrich Schiller University, Jena, Germany 93. Institute of Human Genetics, Münster, Germany 94. University of Southampton Faculty of Medicine, Southampton University Hospitals NHS Trust, Southampton, UK 95. Oncogenetics Team, The Institute of Cancer Research and Royal Marsden NHS Foundation Trust, Sutton, UK 96. Department of Oncology, Rigshospitalet, Copenhagen University Hospital, Copenhagen, Denmark 97. Institute of Human Genetics, Friedrich-Alexander-University Erlangen-Nuremberg, Erlangen, Germany 98. Department of Biostatistics & Bioinformatics, Roswell park Institute, Buffalo, NY, USA 99. Institute for Medical Informatics, Statistics and Epidemiology, University of Leipzig, Leipzig, Germany 100. Genomic Medicine, Manchester Academic Health Sciences Centre, University of Manchester, Central Manchester University Hospitals NHS Foundation Trust, Manchester, UK 101. University of California at Los Angeles, David Geffen School of Medicine, Department of Medicine, Division of Hematology and Oncology, Los Angeles, CA, USA 102. Laboratoire de Génétique Chromosomique, Hôtel Dieu Centre Hospitalier, Chambéry, France 103. Department of Surgery and Cancer, Imperial College London, London, UK 104. Molecular Diagnostics Laboratory, INRASTES, National Centre for Scientific Research "Demokritos", Aghia Paraskevi Attikis, Athens, Greece 105. Program in Cancer Genetics, Departments of Human Genetics and Oncology, McGill University, Montreal, Quebec, Canada 106. Department of Medical Oncology, Papageorgiou, Hospital, Aristotle University of Thessaloniki School of Medicine, Thessaloniki, Greece 107. Biostatistics and Informatics Shared Resource, University of Kansas Medical Center, Kansas City, KS, USA 108. Harvard TH Chan School of Public Health and Dana Farber Cancer Institute, 1101 Dana Building, 450 Brookline Ave, Boston, MA 02215, USA 109. The Susanne Levy Gertner Oncogenetics Unit, Institute of Human Genetics, Chaim Sheba Medical Center, Ramat Gan 52621, and Sackler Faculty of Medicine, Tel Aviv University, Ramat Aviv 69978, Israel 110. UCLA Schools of Medicine and Public Health, Division of Cancer Prevention and Control Research, Jonsson Comprehensive Cancer Center, Los Angeles, CA, USA 111. Cancer Risk and Prevention Clinic, Dana Farber Cancer Institute, Boston, MA, USA 112. Centre of Familial Breast and Ovarian Cancer, Department of Medical Genetics, Institute of Human Genetics, University Würzburg, Germany 113. Women's Cancer, Institute for Women's Health, University College London, London, United Kingdom 114. Department of Clinical Genetics, Rigshospitalet, Copenhagen University Hospital, Copenhagen, Denmark 115. Centre for Epidemiology and Biostatistics, School of Population and Global Health, University of Melbourne, Australia 116. Department of Epidemiology and Preventive Medicine, Monash University, Melbourne, VIC, Australia 117. The Beatson West of Scotland Cancer Centre, Glasgow, UK 118. Ontario Cancer Genetics Network: Samuel Lunenfeld Research Institute, Mount Sinai Hospital, Toronto, Ontario M5G 1X5 119. Department of Pathology and Laboratory Medicine, University of Kansas Medical Center, Kansas City, KS, USA 120. Department of Dermatology, University of Utah School of Medicine, Salt Lake City, Utah, USA 121. Department of Medicine, Royal Marsden Hospital, London, UK 122. Clinical Genetics Branch,DCEG, NCI, NIH, 9609 Medical Center Drive, Room 6E-454, Bethesda, MD, USA 123. Department of Genetics and Pathology, Pomeranian Medical University, Szczecin, Poland 124. Keck School of Medicine, and Norris Comprehensive Cancer Center, University of Southern California, Los Angeles, CA 125. Center for Familial Breast and Ovarian Cancer, Center for Integrated Oncology (CIO) and Center for Molecular Medicine Cologne (CMMC), University Hospital Cologne, Cologne, Germany 126. Department of Preventive Medicine, Keck School of Medicine, University of Southern California Norris Comprehensive Cancer Center, Los Angeles, CA, USA 127. Karolinska Institutet, Department of Environmental Medicine, Division of Nutritional Epidemiology, SE-171 77 STOCKHOLM, Sweden 128. Molecular Genetics of Breast Cancer, Deutsches Krebsforschungszentrum (DKFZ), Heidelberg, Germany 129. Center for Genomic Medicine, Rigshospitalet, Copenhagen University Hospital, Copenhagen, Denmark 130. Institute of Genetic Medicine, Centre for Life, Newcastle Upon Tyne Hospitals NHS Trust, Newcastle upon Tyne, UK 131. Department of Epidemiology, The University of Texas MD Anderson Cancer Center, Houston, TX, USA 132. Clinics of Obstetrics and Gynaecology,

Hannover Medical School, Hannover, Germany 133. Medical Genetics Unit, St George’s, University of London, UK 134. Department of Gynecology, Rigshospitalet, University of Copenhagen, Copenhagen, Denmark 135. Department of Virus, Lifestyle and Genes, Danish Cancer Society Research Center, Copenhagen, Denmark 136. Molecular Unit, Department of Pathology, Herlev Hospital, University of Copenhagen, Copenhagen, Denmark 137. Family Cancer Clinic, Netherlands Cancer Institute, Amsterdam, The Netherlands 138. Department of Medical Oncology, Family Cancer Clinic, Erasmus University Medical Center, Rotterdam, The Netherlands 139. Center For Immunotherapy, Roswell Park Cancer Institute, Buffalo, NY, USA 140. Center for Medical Genetics, NorthShore University HealthSystem, University of Chicago Pritzker School of Medicine, 1000 Central Street, Suite 620,Evanston, IL 60201,US 141. Program in Genetic Epidemiology and Statistical Genetics, Department of Epidemiology, The Harvard T.H. Chan School of Public Health, 677 Huntington Avenue, Boston, MA, 02115, USA 142. British Columbia’s Ovarian Cancer Research (OVCARE) Program, Vancouver General Hospital, BC Cancer Agency and University of British Columbia; Departments of Pathology and Laboratory Medicine, Obstetrics and Gynaecology and Molecular Oncology, Vancouver, British Columbia, CANADA 143. N.N. Petrov Institute of Oncology, St.Petersburg, Russia 144. Lombardi Comprehensive Cancer Center, Georgetown University, 3800 Reservoir Road NW, Washington, DC, USA 145. Department of Statistical Science, Duke University, Durham, North Carolina, USA 146. Clinical Genetics, Guy’s and St. Thomas’ NHS Foundation Trust, London, UK 147. Familial Cancer Centre, Peter MacCallum Cancer Centre, Locked Bag 1, A’Beckett Street, Melbourne, VIC 8006 AUSTRALIA; Sir Peter MacCallum Dept of Oncology, University of Melbourne, VIC 3010 148. Vilnius University Hospital Santariskiu Clinics, Hematology, Oncology and Transfusion Medicine Center, Department of Molecular and Regenerative Medicine, Vilnius, Lithuania 149. State Research Institute Centre for Innovative Medicine, Vilnius, Lithuania 150. Department of Obstetrics and Gynecology Lund University Hospital, Lund Sweden 151. Department of Clinical Genetics, Aarhus University Hospital, Brendstrupgaardsvej 21C, Aarhus N, Denmark 152. Department of Epidemiology, Cancer Prevention Institute of California, Fremont, California, USA 153. Division of Genetics and Epidemiology, Institute of Cancer Research, London, UK 154. Department of Gynaecology and Obstetrics, University Hospital Carl Gustav Carus, Technical University Dresden, Germany 155. Department of Obstetrics and Gynecology, University of Helsinki and Helsinki University Central Hospital, Helsinki, HUS, Finland 156. Department of Surgery, Breast Care Center, Daerim St. Mary’s Hospital, 657 Siheungdaero, Yeongdeungpo-gu, Seoul, 150-822, Korea 157. Department of Pathology, University of Calgary, Calgary, Alberta, Canada 158. Department of Clinical Genetics, Odense University Hospital, Odense C, Denmark 159. The Hong Kong Hereditary Breast Cancer Family Registry, Hong Kong 160. Department of Surgery, The University of Hong Kong, Hong Kong 161. Cancer Genetics Center and Department of Surgery, Hong Kong Sanatorium and Hospital, Hong Kong 162. Vesalius Research Center, VIB, Leuven, Belgium 163. Laboratory for Translational Genetics, Department of Oncology, KULeuven, Belgium 164. Public Health Division of Gipuzkoa, Regional Government of the Basque Country, Spain 165. CIBER of Epidemiology and Public Health (CIBERESP), Spain 166. Molecular Diagnostic Unit, Hereditary Cancer Program, IDIBELL-Catalan Institute of Oncology, Barcelona, Spain 167. Cancer Control Research, BC Cancer Agency, Vancouver, BC, Canada 168. Cancer Epidemiology Program, University of Hawaii Cancer Center, Hawaii, USA 169Department of Surgery, Ulsan College of Medicine and Asan Medical Center, Seoul, Korea 170. Department of Gynecological Oncology, Roswell Park Cancer Institute, Buffalo, NY, USA 171. Département de Génétique, Centre Hospitalier Universitaire de Grenoble, BP 217, Grenoble Cedex 9, France 172. Institut Curie, PSL Research Unviersity and Inserm, U900, Paris, France; Mines Paris Tech, Fontainebleau, France 173. College of Pharmacy and Health Sciences, Texas Southern University, Houston, Texas, USA 174. Cancer Center Wolfsburg, Clinics of Gynaecology, Wolfsburg, Germany 175. Department of Health Sciences Research, Division of Epidemiology, Mayo Clinic, Rochester, MN, USA 176Division of Epidemiology, Department of Medicine, Vanderbilt Epidemiology Center and Vanderbilt-Ingram Cancer Center, Vanderbilt University School of Medicine, Nashville, TN, USA 177. Department of Cancer Epidemiology and Prevention, Maria Sklodowska-Curie Memorial Cancer Center and Institute of Oncology, Warsaw, Poland 178. Department of Gynecologic Oncology, The University of Texas MD Anderson Cancer Center, Houston, TX, USA 179. The Juliane Marie Centre, Department of Gynecology, Rigshospitalet, University of Copenhagen, Copenhagen, Denmark 180. INSERM U1052, CNRS UMR5286, Université Lyon 1, Centre de Recherche en Cancérologie de Lyon, Centre Léon Bérard, Lyon, France 181. Ovarian Cancer Research (OVCARE) Program - Gynecologic Tissue Bank, Vancouver General Hospital and BC Cancer Agency, Vancouver, British Columbia CANADA 182. Department of Health Research and Policy - Epidemiology, Stanford University School of Medicine, Stanford, CA, USA 183. Public Health Ontario, Toronto, ON, Canada 184. Institute of Cancer Sciences, University of Glasgow, Wolfson Wohl Cancer Research Centre, Beatson Institute for Cancer Research, Glasgow, UK 185. Department of Clinical Genetics, VU University Medical Centre, P.O. Box 7057, 1007 MB Amsterdam, the Netherlands 186. Department of Gynaecology and Obstetrics, Division of Tumor Genetics, Klinikum rechts der Isar, Technical University of Munich, Munich, Germany 187. Department of Human Genetics, Radboud University Medical Centre, Nijmegen, The Netherlands 188. Department of Epidemiology and Biostatistics, School of Public Health, Imperial College London, London, W2 1PG, UK 189. Sir Peter MacCallum Department of Oncology, University of Melbourne, Parkville, VIC, Australia 190. Division of Gynecologic Oncology, Department of Obstetrics, Gynecology and Reproductive Sciences, University of Pittsburgh School of Medicine, Pittsburgh, Pennsylvania, USA 191. Department of Epidemiology, University of Pittsburgh Graduate School of Public Health, Pittsburgh, Pennsylvania, USA 192. Ovarian Cancer Center of Excellence, Womens Cancer Research Program, MageeWomens Research Institute and University of Pittsburgh Cancer Institute, Pittsburgh, Pennsylvania, USA 193. Department of Obstetrics and Gynecology, Oregon Health and Science University, Portland, OR, USA 194. Knight Cancer Institute, Oregon Health & Science University, Portland, Oregon, USA 195. Immunology and Molecular Oncology Unit, Veneto Institute of Oncology IOV - IRCCS, Via Gattamelata 64, Padua, Italy

196. Department of Laboratory Medicine and Pathobiology, University of Toronto, Toronto, ON, Canada 197. Department of Laboratory Medicine, and the Keenan Research Centre of the Li Ka Shing Knowledge Institute, St Michael's Hospital, Toronto, ON, Canada 198. Clinical Genetics Research Laboratory, Department of Medicine, Memorial Sloan Kettering Cancer Center, 1275 York Avenue, New York, NY 10044, USA 199. Department of Pathology, Rigshospitalet, University of Copenhagen, Denmark 200. The University of Texas School of Public Health, Houston, TX, USA 201. Department of Gynaecology and Obstetrics, University Hospital Düsseldorf, HeinrichHeine University Düsseldorf, Germany 202. Invitae Corporation and University of Southern California, San Francisco, 513 Parnassus Ave., HSE 901E, San Francisco, CA. 94143 – 0794 203. Department of Molecular Genetics, National Institute of Oncology, Budapest, Hungary 204. Center for Clinical Cancer Genetics and Global Health, University of Chicago Medical Center, 5841 South Maryland Avenue, MC 2115 Chicago, IL, USA 205. Oncology, Clinical Sciences in Lund, Lund University, 221 85 Lund, Sweden 206. The Ohio State University and the James Cancer Center, Columbus, Ohio, USA 207. West Midlands Regional Genetics Service, Birmingham Women's Hospital Healthcare NHS Trust, Edgbaston, Birmingham, UK 208. Julius Center for Health Sciences and Primary Care, UMC Utrecht, Utrecht, the Netherlands 209. The Breast Cancer Now Toby Robins Research Centre, The Institute of Cancer Research, 237 Fulham Road, London SW3 6JB, UK 210. Molecular and Nutritional Epidemiology Unit, Cancer Research and Prevention Institute ISPO, Florence, Italy 211. Unit of Medical Genetics, Department of Biomedical, Experimental and Clinical Sciences, University of Florence, Florence, Italy 212. Cancer Research UK Clinical Trials Unit, Institute of Cancer Sciences, University of Glasgow, UK 213. Department of Epidemiology, University of Michigan School of Public Health, Ann Arbor, Michigan, USA 214. Section of Molecular Diagnostics, Clinical Biochemistry, Aalborg University Hospital, Aalborg, Denmark 215. Department of Genetics, Portuguese Oncology Institute, Rua Dr. António Bernardino de Almeida, 4200-072 Porto, Portugal 216. IFOM, The FIRC (Italian Foundation for Cancer Research) Institute of Molecular Oncology, c/o IFOM-IEO campus, via Adamello 16 20139 Milan, Italy 217. Dept of OB/GYN, Medical University of Vienna and Comprehensive Cancer Center, Vienna, Austria, Waehringer Guertel 18-20, A 1090 Vienna, Austria 218. Division of Cancer Medicine, Peter MacCallum Cancer Centre, Locked Bag 1, A'Beckett St, East Melbourne, Victoria 8006, Australia 219. NRG Oncology, Statistics and Data Management Center, Roswell Park Cancer Institute, Elm St & Carlton St, Buffalo, NY 14263, USA 220. Department of Epidemiology and Biostatistics, Memorial Sloan Kettering Cancer Center, New York, NY, USA 221. Channing Division of Network Medicine, Brigham and Women's Hospital and Harvard Medical School, Boston, MA, USA 222. South East of Scotland Regional Genetics Service, Western General Hospital, Edinburgh, UK 223. Service de Génétique Clinique Chromosomique et Moléculaire, Centre Hospitalier Universitaire de St Etienne, St Etienne, France 224. Translational Research Laboratory, IDIBELL (Bellvitge Biomedical Research Institute),Catalan Institute of Oncology, Barcelona, Spain 225Unité d'Oncogénétique, CHU Arnaud de Villeneuve, Montpellier, France 226. Unit of "Predictive Medicine: Molecular Bases of Genetic Risk", Department of Experimental Oncology, Fondazione IRCCS Istituto Nazionale dei Tumori, Milan, Italy 227. Department of Clinical Genetics, Karolinska University Hospital, Stockholm, Sweden 228. Clalit National Israeli Cancer Control Center and Department of Community Medicine and Epidemiology, Carmel Medical Center and B. Rappaport Faculty of Medicine, Haifa, Israel 229. Brigham and Women's Hospital, Dana-Farber Cancer Institute, Boston, MA USA 230. Clinical Genetics Service, Department of Medicine, Memorial Sloan Kettering Cancer Center, New York, NY, USA 231. Division of Gynecologic Oncology, NorthShore University HealthSystem, University of Chicago, Evanston, IL, USA 232. Hereditary Endocrine Cancer group, Spanish National Cancer Research Center (CNIO), and Biomedical Network on Rare Diseases (CIBERER), Madrid, Spain 233. Department of Epidemiology, Netherlands Cancer Institute, Amsterdam, The Netherlands 234. Program in Epidemiology, Division of Public Health Sciences, Fred Hutchinson Cancer Research Center, Seattle, WA, USA 235. Epidemiology Branch, Division of Intramural Research, National Institute of Environmental Health Sciences, NIH, Research Triangle Park, NC, USA 236. Clinical Cancer Genetics Program, Division of Human Genetics, Department of Internal Medicine, The Comprehensive Cancer Center, The Ohio State University, Columbus, USA 237. Université Paris-Saclay, Université Paris-Sud, UVSQ, CESP, INSERM, Villejuif, France 238. Gustave Roussy, F-94805, Villejuif, France 239Human Genetics Foundation (HuGeF), Torino, Italy 240. Cancer Council Victoria and University of Melbourne, Australia 241. Department of Hematology and Oncology, University of Kansas Medical Center, Kansas City, KS, USA 242Department of Gynaecological Oncology, Glasgow Royal Infirmary, Glasgow, UK 243. North East Thames Regional Genetics Service, Great Ormond Street Hospital for Children NHS Trust, London, UK 244. Département Oncologie Génétique, Prévention et Dépistage, INSERM CIC-P9502, Institut Paoli-Calmettes/Université d'Aix-Marseille II, Marseille, France 245Genetic Epidemiology Laboratory, Department of Pathology, University of Melbourne, Parkville, VIC, Australia 246. Institute of Human Genetics, Hannover Medical School, Hannover, Germany 247. Department of Cancer Prevention and Control, Roswell Park Cancer Institute, Buffalo, NY, USA 248. Epidemiology Center, College of Medicine, University of South Florida, Tampa, Florida, USA 249. Institute of Human Genetics, Department of Human Genetics, University Hospital Heidelberg, Germany 250. Division of Breast Cancer Research, The Institute of Cancer Research, London, UK 251. National Human Genome Research Institute, National Institutes of Health, Bethesda, MD, USA 252. Department of Genetics, Portuguese Oncology Institute, Rua Dr. António Bernardino de Almeida, 4200-072 Porto, Portugal and Biomedical Sciences Institute (ICBAS), University of Porto, Porto, Portugal 253. Cancer Research Initiatives Foundation, Sime Darby Medical Centre, Subang Jaya, Malaysia 254. University Malaya Cancer Research Institute, Faculty of Medicine, University Malaya Medical Centre, University Malaya, Kuala Lumpur, Malaysia 255. Department of Epidemiology, Harvard T. Chan School of Public Health, Boston, MA, USA 256. Cancer Prevention and Control, Samuel Oschin Comprehensive Cancer Institute, CedarsSinai Medical Center, Los Angeles, CA, USA 257Magee-Womens Hospital of UPMC, University of Pittsburgh School of Medicine, Pittsburgh, PA, USA 258. Latvian Biomedical Research and Study Centre. Ratsupites str 1, Riga, Latvia 259. Ovarian Cancer Research (OVCARE)

Program - Cheryl Brown Ovarian Cancer Outcomes Unit (CBOCOU), BC Cancer Agency, Vancouver, British Columbia CANADA 260. Department of Medical Genetics, Box 134, Level 6 Addenbrooke's Treatment Centre, Addenbrooke's Hosptital, Hills Road, Cambridge CB2 0QQ, UK 261. Divison of Human Cancer Genetics, Departments of Internal Medicine and Molecular Virology, Immunology and Medical Genetics, Comprehensive Cancer Center, The Ohio State University, Columbus, OH, USA 262. Cancer Epidemiology Unit, Nuffield Department of Population Health, University of Oxford, Oxford, UK 263. Hellenic Health Foundation, Athens, Greece 264WHO Collaborating Center for Nutrition and Health, Unit of Nutritional Epidemiology and Nutrition in Public Health, Dept. of Hygiene, Epidemiology and Medical Statistics, University of Athens Medical School, Greece 265. Department of Medical Oncology, Beth Israel Deaconess Medical Center, Boston, MA, USA 266Division of Gynecologic Oncology, Department of Obstetrics and Gynaecology, Radboud University Medical Centre, Nijmegen, The Netherlands 267. Department of Genetics, University Medical Center, Groningen University, Groningen, The Netherlands 268. Department of Medical Genetics, University Medical Center Utrecht, Utrecht, The Netherlands 269. Division of Gynecologic Oncology , Department of Obstetrics and Gynaecology and Leuven Cancer Institute, University Hospitals Leuven, Leuven, Belgium 270. Institute of Human Genetics, Campus Virchov Klinikum, Charite Berlin, Germany 271. Fundación Pública Galega de Medicina Xenómica, Servizo Galego de Saúde (SERGAS), Instituto de Investigaciones Sanitarias (IDIS), Santiago de Compostela, Spain 272. Grupo de Medicina Xenómica, Centro de Investigación Biomédica en Red de Enfermedades Raras (CIBERER), Universidade de Santiago de Compostela (USC), Santiago de Compostela, Spain 273. Vanderbilt Epidemiology Center, Vanderbilt Genetics Institute, Department of Obstetrics and Gynecology, Vanderbilt University Medical Center, Nashville, TN, USA 274. Oxford Regional Genetics Service, Churchill Hospital, Oxford, UK 275. Institute of Human Genetics, University Hospital, Leipzig, Germany 276. Department of Obstetrics and Gynecology, University of Ulm, Ulm, Germany 277. Population Health Department, QIMR Berghofer Medical Research Institute, 300 Herston Road, Herston, QLD 4006, Australia 278. Biostatistics and Computational Biology Branch, Division of Intramural Research, National Institute of Environmental Health Sciences, NIH, Research Triangle Park, NC, USA 279. Clinical Cancer Genetics, City of Hope, Duarte, CA, USA 280. Department of Data Management Science-Stanford University School of Medicine, Stanford, CA, USA 281. Department of Human Genetics and Department of Clinical Genetics, Leiden University Medical Center, Leiden, The Netherlands 282. Department of Epidemiology, University of California Irvine, Irvine, CA, USA 283. Women's College Research Institute, University of Toronto, Toronto, ON, Canada 284. Center for Genomic Medicine, Department of Biomedical Data Science, Geisel School of Medicine at Dartmouth, Williamson Translational Research Building, Room HB 7261, Lebanon, NH 03756, USA 285. Department of Public Health Sciences, The University of Virginia, Charlottesville, VA, USA 286. School of Women's and Children's Health, Lowy Cancer Research Centre, The University of New South Wales UNSW Sydney NSW 2052 AUSTRALIA 287. The Kinghorn Cancer Centre, Garvan Institute of Medical Research, 384 Victoria Street, Darlinghurst NSW 2010, Australia 288. Department of Molecular Medicine, University La Sapienza, c/o Policlinico Umberto I, viale Regina Elena 324, 00161 Rome, Italy 289. Community and Population Health Research Institute, Department of Biomedical Sciences, Cedars-Sinai Medical Center, Los Angeles, California, USA 290Department of Preventive Medicine, College of Medicine, Seoul National University, 103 Daehak-ro, Jongno-gu, Seoul 110-799, Korea 291. Seoul National University Cancer Research Institute, 103 Daehak-ro, Jongno-gu, Seoul 110-799, Korea 292. Department of Biomedical Science, Graduate School, Seoul National University, 103 Daehak-ro, Jongno-gu, Seoul 110-799, Korea 293. Department of Public Health Sciences, College of Medicine, Medical University of South Carolina, Charleston, SC 29425 294. Department of Chronic Disease Epidemiology, Yale School of Public Health, New Haven, CT, USA 295. Department of Biomedical Sciences, Cedars-Sinai Medical Center, Los Angeles, California, USA

#### The Breast Cancer Association Consortium

K Michailidou[1,2], S Lindström[3], J Dennis[1], MK Bolla[1],Q Wang[1], R Keeman[4], S Behrens[5], H Anton-Culver[6], KJ Aronson[7], PL Auer[8,9], J Benitez[10,11], SE Bojesen[12,13,14], H Brauch[15,16,17], H Brenner[17,18,19], B Burwinkel[20,21], F Canzian[22], JE Castelao[23], JY Choi[24,25], CL Clarke[26], NBCS Collaborators, A Cox[27], K Czene[28], MB Daly[29], P Devilee[30,31], T Dörk[32], M Dwek[33], DM Eccles[34], PA Fasching[35,36], L Fritschi[37], M Gago-Dominguez[38,39], SM Gapstur[40], JA García-Sáenz[41], GG Giles[42,43], MS Goldberg[44,45], DE Goldgar[46], P Guénel[47], CA Haiman[48], U Hamann[49], M Hartman[50,51], A Hollestelle[52], MJ Hooning[52], JL Hopper[43], M Iwasaki[53], A Jakubowska[54], EM John[55], R Kaaks[5], D Kang[24,25,56], VN Kristensen[57,58], A Kwong[59,60,61], D Lambrechts[62,63], A Lindblom[64], J Lubinski[54], C Luccarini[65], A Mannermaa[66,67,68], S Margolin[69], K Matsuo[70,71], U Menon[72], K Muir[73,74], SL Neuhausen[75], H Nevanlinna[76], OI Olopade[77], H Jernström[78], SK Park[24,25,56], P Peterlongo[79], J Peto[80], KA Phillips[43,81,82,83], D Plaseska-Karanfilska[84], R Prentice[8], P Radice[85], G Rennert[86], A Romero[87], E Saloustros[88], DP Sandler[89], EJ Sawyer[90], RK Schmutzler[91,92,93], CY Shen[94,95], XO Shu[96], MC Southey[42,97,98], JJ Spinelli[99,100], J Stone[43,101], A Swerdlow[102,103], R Tamimi[104,105,106], JA Taylor[89,107], MB Terry[108], C Vachon[109], CR Weinberg[110], C Wendt[111], R Winqvist[112,113], A Wolk[114], AH Wu[48], W Zheng[96], E Ziv[115]; ABCTB Investigators; kConFab/AOCS Investigators, AC Antoniou[1], IL Andrulis[116,117], FJ Couch[118], PDP Pharoah[1,65], J Chang-Claude[5,119], P Hall[28,69], DJ Hunter[105,106], RL Milne[42,43],

M García-Closas[120], MK Schmidt[4,121], SJ Chanock[120], AM Dunning[65], G Chenevix-Trench[122] , J Simard[123], P Kraft[105,106] and DF Easton[1,65]

1. Centre for Cancer Genetic Epidemiology, Department of Public Health and Primary Care, University of Cambridge, Cambridge, UK 2. Department of Electron Microscopy/Molecular Pathology, The Cyprus Institute of Neurology and Genetics, Nicosia, Cyprus 3. Department of Epidemiology, University of Washington School of Public Health, Seattle, WA, USA 4. Division of Molecular Pathology, The Netherlands Cancer Institute - Antoni van Leeuwenhoek Hospital, Amsterdam, The Netherlands 5. Division of Cancer Epidemiology, German Cancer Research Center (DKFZ), Heidelberg, Germany 6. Department of Epidemiology, University of California Irvine, Irvine, CA, USA 7. Department of Public Health Sciences, and Cancer Research Institute, Queen's University, Kingston, ON, Canada 8. Cancer Prevention Program, Fred Hutchinson Cancer Research Center, Seattle, WA, USA 9. Zilber School of Public Health, University of Wisconsin-Milwaukee, Milwaukee, WI, USA 10. Human Cancer Genetics Program, Spanish National Cancer Research Centre, Madrid, Spain 11. Centro de Investigación en Red de Enfermedades Raras (CIBERER), Valencia, Spain 12. Copenhagen General Population Study, Herlev and Gentofte Hospital, Copenhagen University Hospital, Herlev, Denmark 13. Department of Clinical Biochemistry, Herlev and Gentofte Hospital, Copenhagen University Hospital, Herlev, Denmark 14. Faculty of Health and Medical Sciences, University of Copenhagen, Copenhagen, Denmark 15. Dr. Margarete Fischer-Bosch-Institute of Clinical Pharmacology, Stuttgart, Germany 16. iFIT-Cluster of Excellence, University of Tübingen, Tübingen, Germany 17. German Cancer Consortium (DKTK), German Cancer Research Center (DKFZ), Heidelberg, Germany 18. Division of Clinical Epidemiology and Aging Research, German Cancer Research Center (DKFZ), Heidelberg, Germany 19. Division of Preventive Oncology, German Cancer Research Center (DKFZ) and National Center for Tumor Diseases (NCT), Heidelberg, Germany 20. Molecular Epidemiology Group, C080, German Cancer Research Center (DKFZ), Heidelberg, Germany 21. Molecular Biology of Breast Cancer, University Womens Clinic Heidelberg, University of Heidelberg, Heidelberg, Germany 22. Genomic Epidemiology Group, German Cancer Research Center (DKFZ), Heidelberg, Germany 23. Oncology and Genetics Unit, Instituto de Investigacion Biomedica (IBI) Orense-PontevedraVigo, Xerencia de Xestion Integrada de Vigo-SERGAS, Vigo, Spain 24. Department of Biomedical Sciences, Seoul National University Graduate School, Seoul, Korea 25. Cancer Research Institute, Seoul National University, Seoul, Korea 26. Westmead Institute for Medical Research, University of Sydney, Sydney, Australia 27. Sheffield Institute for Nucleic Acids (SInFoNiA), Department of Oncology and Metabolism, University of Sheffield, Sheffield, UK 28. Department of Medical Epidemiology and Biostatistics, Karolinska Institutet, Stockholm, Sweden 29. Department of Clinical Genetics, Fox Chase Cancer Center, Philadelphia, PA, USA 30. Department of Pathology, Leiden University Medical Center, Leiden, The Netherlands 31. Department of Human Genetics, Leiden University Medical Center, Leiden, The Netherlands 32. Gynaecology Research Unit, Hannover Medical School, Hannover, Germany 33. School of Life Sciences, University of Westminster, London, UK 34. Faculty of Medicine, University of Southampton, Southampton, UK 35. Institute of Human Genetics, University Hospital Erlangen, Friedrich-Alexander University Erlangen-Nuremberg, Comprehensive Cancer Center Erlangen-EMN, Erlangen, Germany 36. David Geffen School of Medicine, Department of Medicine Division of Hematology and Oncology, University of California at Los Angeles, Los Angeles, CA, USA 37. School of Public Health, Curtin University, Perth, Australia 38. Genomic Medicine Group, Galician Foundation of Genomic Medicine, Instituto de Investigación Sanitaria de Santiago de Compostela (IDIS), Complejo Hospitalario Universitario de Santiago, SERGAS, Santiago de Compostela, Spain 39. Moores Cancer Center, University of California San Diego, La Jolla, CA, USA 40. Behavioral and Epidemiology Research Group, American Cancer Society, Atlanta, GA, USA 41. Medical Oncology Department, Hospital Clínico San Carlos, IdISSC (Centro Investigacion Biomedica en Red), CIBERONC (Instituto de Investigación Sanitaria San Carlos), Madrid, Spain 42. Cancer Epidemiology Division, Cancer Council Victoria, Melbourne, Victoria, Australia 43. Centre for Epidemiology and Biostatistics, Melbourne School of Population and Global Health, The University of Melbourne, Melbourne, Australia 44. Department of Medicine, McGill University, Montréal, QC, Canada 45. Division of Clinical Epidemiology, Royal Victoria Hospital, McGill University, Montréal, QC, Canada 46. Department of Dermatology, Huntsman Cancer Institute, University of Utah School of Medicine, Salt Lake City, UT, USA 47. Cancer & Environment Group, Center for Research in Epidemiology and Population Health (CESP), INSERM, University Paris-Sud, University Paris-Saclay, Villejuif, France 48. Department of Preventive Medicine, Keck School of Medicine, University of Southern California, Los Angeles, CA, USA 49. Molecular Genetics of Breast Cancer, German Cancer Research Center (DKFZ), Heidelberg, Germany 50. Saw Swee Hock School of Public Health, National University of Singapore, Singapore, Singapore 51. Department of Surgery, National University Health System, Singapore, Singapore 52. Department of Medical Oncology, Family Cancer Clinic, Erasmus MC Cancer Institute, Rotterdam, The Netherlands 53. Division of Epidemiology, Center for Public Health Sciences, National Cancer Center, Tokyo, Japan 54. Department of Genetics and Pathology, Pomeranian Medical University, Szczecin, Poland 55. Department of Medicine, Division of Oncology, Stanford Cancer Institute, Stanford University School of Medicine, Stanford, CA, USA 56. Department of Preventive Medicine, Seoul National University College of Medicine, Seoul, Korea 57. Department of Cancer Genetics, Institute for Cancer Research, Oslo University Hospital Radiumhospitalet, Oslo, Norway 58. Institute of Clinical Medicine, Faculty of Medicine, University of Oslo, Oslo, Norway 59. Hong Kong Hereditary Breast Cancer Family Registry, Happy Valley, Hong Kong 60. Department of Surgery, The University of Hong Kong, Pok Fu Lam, Hong Kong 61. Department of Surgery, Hong Kong Sanatorium and Hospital, Happy Valley, Hong Kong 62. VIB Center for Cancer Biology, Leuven, Belgium 63. Laboratory for Translational Genetics, Department of Human Genetics, University of Leuven, Leuven, Belgium 64. Department of Molecular Medicine and Surgery, Karolinska Institutet, Stockholm, Sweden 65. Centre for Cancer

Genetic Epidemiology, Department of Oncology, University of Cambridge, Cambridge, UK 66. Translational Cancer Research Area, University of Eastern Finland, Kuopio, Finland 67. Institute of Clinical Medicine, Pathology and Forensic Medicine, University of Eastern Finland, Kuopio, Finland 68. Imaging Center, Department of Clinical Pathology, Kuopio University Hospital, Kuopio, Finland 69. Department of Oncology, Södersjukhuset, Stockholm, Sweden 70. Division of Cancer Epidemiology and Prevention, Aichi Cancer Center Research Institute, Nagoya, Japan 71. Division of Cancer Epidemiology, Nagoya University Graduate School of Medicine, Nagoya, Japan 72. MRC Clinical Trials Unit at UCL, Institute of Clinical Trials & Methodology, University College London, London, UK 73. Division of Population Health, Health Services Research and Primary Care, School of Health Sciences, Faculty of Biology, Medicine and Health, The University of Manchester, Manchester, UK 74. Division of Health Sciences, Warwick Medical School, University of Warwick, Coventry, UK 75. Department of Population Sciences, Beckman Research Institute of City of Hope, Duarte, CA, USA 76. Department of Obstetrics and Gynecology, Helsinki University Hospital, University of Helsinki, Helsinki, Finland 77. Center for Clinical Cancer Genetics, The University of Chicago, Chicago, IL, USA 78. Oncology, Clinical Sciences in Lund, Lund University, 221 85 Lund, Sweden 79. Genome Diagnostics Program, IFOM - the FIRC (Italian Foundation for Cancer Research) Institute of Molecular Oncology, Milan, Italy 80. Department of Non-Communicable Disease Epidemiology, London School of Hygiene and Tropical Medicine, London, UK 81. Peter MacCallum Cancer Center, Melbourne, Australia 82. Sir Peter MacCallum Department of Oncology, The University of Melbourne, Melbourne, Australia 83. Department of Medicine, St Vincent's Hospital, The University of Melbourne, Fitzroy, Australia 84. Research Centre for Genetic Engineering and Biotechnology "Georgi D. Efremov", MASA, Skopje, Republic of North Macedonia 85. Unit of "Predictive Medicine: Molecular Bases of Genetic Risk", Department of Experimental Oncology, Fondazione IRCCS Istituto Nazionale dei Tumori, Milan, Italy. 86. Clalit National Cancer Control Center, Carmel Medical Center and Technion Faculty of Medicine, Haifa, Israel 87. Medical Oncology Department, Hospital Universitario Puerta de Hierro, Madrid, Spain 88. Department of Oncology, University Hospital of Larissa, Larissa, Greece 89. Epidemiology Branch, National Institute of Environmental Health Sciences, NIH, Research Triangle Park, NC, USA 90. Research Oncology, Guy's Hospital, King's College London, London, UK 91. Center for Familial Breast and Ovarian Cancer, Faculty of Medicine and University Hospital Cologne, University of Cologne, Cologne, Germany 92. Center for Integrated Oncology (CIO), Faculty of Medicine and University Hospital Cologne, University of Cologne, Cologne, Germany 93. Center for Molecular Medicine Cologne (CMMC), Faculty of Medicine and University Hospital Cologne, University of Cologne, Cologne, Germany 94. Taiwan Biobank, Institute of Biomedical Sciences, Academia Sinica, Taipei, Taiwan 95. School of Public Health, China Medical University, Taichung, Taiwan 96. Division of Epidemiology, Department of Medicine, Vanderbilt Epidemiology Center, Vanderbilt-Ingram Cancer Center, Vanderbilt University School of Medicine, Nashville, TN, USA 97. Precision Medicine, School of Clinical Sciences at Monash Health, Monash University, Clayton, Victoria, Australia 98. Department of Clinical Pathology, The University of Melbourne, Melbourne, Victoria, Australia 99. Population Oncology, BC Cancer, Vancouver, BC, Canada 100. School of Population and Public Health, University of British Columbia, Vancouver, BC, Canada 101. The Curtin UWA Centre for Genetic Origins of Health and Disease, Curtin University and University of Western Australia, Perth, Australia 102. Division of Genetics and Epidemiology, The Institute of Cancer Research, London, UK 103. Division of Breast Cancer Research, The Institute of Cancer Research, London, UK 104. Channing Division of Network Medicine, Department of Medicine, Brigham and Women's Hospital, Harvard Medical School, Boston, MA, USA 105. Department of Epidemiology, Harvard T.H. Chan School of Public Health, Boston, MA, USA 106. Program in Genetic Epidemiology and Statistical Genetics, Harvard T.H. Chan School of Public Health, Boston, MA, USA 107. Epigenetic and Stem Cell Biology Laboratory, National Institute of Environmental Health Sciences, NIH, Research Triangle Park, NC, USA 108. Department of Epidemiology, Mailman School of Public Health, Columbia University, New York, NY, USA 109. Department of Health Sciences Research, Division of Epidemiology, Mayo Clinic, Rochester, MN, USA 110. Biostatistics and Computational Biology Branch, National Institute of Environmental Health Sciences, NIH, Research Triangle Park, NC, USA 111. Department of Clinical Science and Education, Södersjukhuset, Karolinska Institutet, Stockholm, Sweden 112. Laboratory of Cancer Genetics and Tumor Biology, Cancer and Translational Medicine Research Unit, Biocenter Oulu, University of Oulu, Oulu, Finland 113. Laboratory of Cancer Genetics and Tumor Biology, Northern Finland Laboratory Centre Oulu, Oulu, Finland 114. Institute of Environmental Medicine, Karolinska Institutet, Stockholm, Sweden 115. Department of Medicine, Institute for Human Genetics, UCSF Helen Diller Family Comprehensive Cancer Center, University of California San Francisco, San Francisco, CA, USA 116. Fred A. Litwin Center for Cancer Genetics, Lunenfeld-Tanenbaum Research Institute of Mount Sinai Hospital, Toronto, ON, Canada 117. Department of Molecular Genetics, University of Toronto, Toronto, ON, Canada 118. Department of Laboratory Medicine and Pathology, Mayo Clinic, Rochester, MN, USA 119. Cancer Epidemiology Group, University Cancer Center Hamburg (UCCH), University Medical Center Hamburg-Eppendorf, Hamburg, Germany 120. Division of Cancer Epidemiology and Genetics, National Cancer Institute, National Institutes of Health, Department of Health and Human Services, Bethesda, MD, USA 121. Division of Psychosocial Research and Epidemiology, The Netherlands Cancer Institute - Antoni van Leeuwenhoek hospital, Amsterdam, The Netherlands 122. Department of Genetics and Computational Biology, QIMR Berghofer Medical Research Institute, Brisbane, Australia 123. Genomics Center, Centre Hospitalier Universitaire de Québec – Université Laval Research Center, Québec City, QC, Canada

#### Study-specific text & acknowledgements

**ALSPAC**
**Funding**: The UK Medical Research Council and Wellcome (Grant ref: 217065/Z/19/Z) and the University of Bristol provide core support for ALSPAC. A comprehensive list of grants funding is available on the ALSPAC website: (http://www.bristol.ac.uk/alspac/external/documents/grant-acknowledgements.pdf).
**Acknowledgements**: We are extremely grateful to all the families who took part in this study, the midwives for their help in recruiting them, and the whole ALSPAC team, which includes interviewers, computer and laboratory technicians, clerical workers, research scientists, volunteers, managers, receptionists and nurses. None of the funders influenced the study design, analyses or interpretation of the findings. This publication is the work of the authors and JRBP will serve as guarantor for the contents of this paper. The views expressed in this paper are those of the authors and not necessarily any of the funders.

**The Atherosclerosis Risk in Communities study**
The Atherosclerosis Risk in Communities study has been funded in whole or in part with Federal funds from the National Heart, Lung, and Blood Institute, National Institutes of Health, Department of Health and Human Services, under Contract nos. (75N92022D00001, 75N92022D00002, 75N92022D00003, 75N92022D00004, 75N92022D00005). The authors thank the staff and participants of the ARIC study for their important contributions.

GWAS was supported by R01HL087641, and R01HL086694; National Human Genome Research Institute contract U01HG004402; and National Institutes of Health contract HHSN268200625226C. Infrastructure was partly supported by Grant Number UL1RR025005, a component of the National Institutes of Health and NIH Roadmap for Medical Research. Funding support for “Building on GWAS for NHLBI-diseases: the U.S. CHARGE consortium” was provided by the NIH through the American Recovery and Reinvestment Act of 2009 (ARRA) (5RC2HL102419).

**INGI-Carlantino**
We are very grateful to the municipal administrators for their collaboration on the project and for logistic support. We would like to thank all participants in the study. The study was funded by Regione FVG (L.26.2008).

**CILENTO**
We address special thanks to the populations of Campora, Gioi, and Cardile for their participation in the study. This work was supported by grants from the Italian Ministry of Universities (PON03PE_00060_7, IDF SHARID ARS01_01270), the Assessorato Ricerca Regione Campania (POR CAMPANIA 2000/2006 MISURA 3.16).

**CROATIA-Korcula**
We would like to acknowledge the contributions of the recruitment team in Korcula, the administrative teams in Croatia and Edinburgh and the people of Korcula. The SNP genotyping for the KORCULA cohort was performed in Helmholtz Zentrum München, Neuherberg, Germany. Funding provided by Medical Research Council UK, the Ministry of Science, Education and Sport in the Republic of Croatia (number 216- 1080315-0302) and Croatian Science Foundation (grant 8875).

**CROATIA-Vis**
We would like to acknowledge the invaluable contributions of the recruitment teams in Vis (including those from the Institute of Anthropological Research in Zagreb and the Croatian Centre for Global Health at the University of Split), the administrative teams in Croatia and Edinburgh and the people of Vis. The CROATIA-Vis study was supported through grants from

the Medical Research Council UK and Ministry of Science, Education and Sport of the Republic of Croatia (number 108-1080315-0302) and the European Union framework program 6 EUROSPAN project (contract no. LSHG-CT-2006-018947). We would like to acknowledge the staff of several institutions in Croatia that supported the field work, including but not limited to The University of Split and Zagreb Medical Schools, the Institute for Antroplogical Research in Zagreb and the Croatian Institute for Public Health. We would like to acknowledge the invaluable contributions of the recruitment team in Korcula, the administrative teams in Croatia and Edinburgh and the participants. The SNP genotyping was performed in Helmholtz Zentrum München, Neuherberg, Germany. The study was funded by the Medical Research Council UK, The Croatian Ministry of Science, Education and Sports (grant 216-1080315-0302), the European Union framework program 6 EUROSPAN project (contract no. LSHG-CT-2006-018947), the Croatian Science Foundation (grant 8875), the Croatian National Center of Research Excellence in Personalized Healthcare (grant number KK.01.1.1.01.0010) and the Center of Competence in Molecular Diagnostics (KK.01.2.2.03.0006).

**EGCUT**
This study was supported by EU H2020 grants 692145, 676550, 654248, Estonian Research Council Grant IUT20-60, NIASC, and EU through the European Regional Development Fund (Project No. 2014- 2020.4.01.15-0012 GENTRANSMED).

**EPIC**
The coordination of EPIC is financially supported by the European Commission (DG-SANCO) and the International Agency for Research on Cancer. The national cohorts (that recruited male participants) are supported by Danish Cancer Society (Denmark); German Cancer Aid, German Cancer Research Center (DKFZ), Federal Ministry of Education and Research (BMBF), Deutsche Krebshilfe, Deutsches Krebsforschungszentrum and Federal Ministry of Education and Research (Germany); the Hellenic Health Foundation (Greece); Associazione Italiana per la Ricerca sul Cancro-AIRC-Italy and National Research Council (Italy); Dutch Ministry of Public Health, Welfare and Sports (VWS), Netherlands Cancer Registry (NKR), LK Research Funds, Dutch Prevention Funds, Dutch ZON (Zorg Onderzoek Nederland), World Cancer Research Fund (WCRF), Statistics Netherlands (The Netherlands); Health Research Fund (FIS), PI13/00061 to Granada, PI13/01162 to EPICMurcia, Regional Governments of Andalucía, Asturias, Basque Country, Murcia and Navarra, ISCIII RETIC (RD06/0020) (Spain); Swedish Cancer Society, Swedish Research Council and County Councils of Skåne and Västerbotten (Sweden); Cancer Research UK (14136 to EPIC-Norfolk; C570/A16491 and C8221/A19170 to EPICOxford), Medical Research Council (1000143 to EPIC-Norfolk, MR/M012190/1 to EPICOxford) (United Kingdom). The EPIC Norfolk Study is funded by Cancer Research United Kingdom and the Medical Research Council. This work was supported by the Medical Research Council [U106179472; MC_U106179472; U106179471; MC_U106179471]. The authors acknowledge the support of all EPICNorfolk staff and participants.

**INGI-FVG**
We are very grateful to the municipal administrators for their collaboration on the project and for logistic support. We would like to thank all participants in the study. The study was funded by Regione FVG (L.26.2008).

**Generation Scotland**
Generation Scotland received core support from the Chief Scientist Office of the Scottish Government Health Directorates [CZD/16/6] and the Scottish Funding Council [HR03006] and is currently supported by the Wellcome Trust [216767/Z/19/Z]. Genotyping of the GS:SFHS samples was carried out by the Genetics Core Laboratory at the Edinburgh Clinical Research Facility, University of Edinburgh, Scotland and was funded by the Medical Research Council UK and the Wellcome Trust (Wellcome Trust Strategic Award "STratifying Resilience and Depression Longitudinally" (STRADL) Reference 104036/Z/14/Z).

**HBCS**
We acknowledge support from grants from the Academy of Finland, the Finnish Diabetes Research Society, Folkhälsan Research Foundation, Novo Nordisk Foundation, Finska Läkaresällskapet, Signe and Ane Gyllenberg Foundation, University of Helsinki, European Science Foundation (EUROSTRESS), Ministry of Education, Ahokas Foundation, Emil Aaltonen Foundation.

**InCHIANTI**
The InCHIANTI study baseline (1998-2000) was supported as a "targeted project" (ICS110.1/RF97.71) by the Italian Ministry of Health and in part by the U.S. National Institute on Aging (Contracts: 263 MD 9164 and 263 MD 821336).

**InterAct**
We thank all EPIC participants and staff for their contribution to the study. We thank staff from the Technical, Field Epidemiology and Data Functional Group Teams of the MRC Epidemiology Unit in Cambridge, UK, for carrying out sample preparation, DNA provision and quality control, genotyping and data- handling work. The EPIC-InterAct study received funding from the European Union (Integrated Project LSHM- CT-2006-037197 in the Framework Programme 6 of the European Community).

**KORA**
KORA F3 and F4: The KORA study was initiated and financed by the Helmholtz Zentrum München – German Research Center for Environmental Health, which is funded by the German Federal Ministry of Education and Research (BMBF) and by the State of Bavaria. Data collection in the KORA study is done in cooperation with the University Hospital of Augsburg. We thank all participants for their long-term commitment to the KORA study, the staff for data collection and research data management and the members of the KORA Study Group (https://www.helmholtz-munich.de/en/epi/cohort/kora) who are responsible for the design and conduct of the study.

**Lifelines**
The Lifelines Biobank initiative has been made possible by funding from the Dutch Ministry of Health, Welfare and Sport, the Dutch Ministry of Economic Affairs, the University Medical Center Groningen (UMCG the Netherlands), University of Groningen and the Northern Provinces of the Netherlands. The generation and management of GWAS genotype data for the Lifelines Cohort Study is supported by the UMCG Genetics Lifelines Initiative (UGLI). UGLI is partly supported by a Spinoza Grant from NWO, awarded to Cisca Wijmenga.

The authors wish to acknowledge the services of the Lifelines Cohort Study, the contributing research centers delivering data to Lifelines, and all the study participants. The LifeLines Cohort Study, and generation and management of GWAS genotype data for the LifeLines Cohort Study is supported by the Netherlands Organization of Scientific Research NWO (grant 175.010.2007.006), the Economic Structure Enhancing Fund (FES) of the Dutch government, the Ministry of Economic Affairs, the Ministry of Education, Culture and Science, the Ministry for Health, Welfare and Sports, the Northern Netherlands Collaboration of Provinces (SNN), the Province of Groningen, University Medical Center Groningen, the University of Groningen, Dutch Kidney Foundation and Dutch Diabetes Research Foundation. The authors wish to acknowledge the services of the Lifelines Cohort Study, the contributing research centers delivering data to Lifelines, and all the study participants.

**NEO**
The authors of the NEO study thank all individuals who participated in the Netherlands Epidemiology in Obesity study, all participating general practitioners for inviting eligible participants and all research nurses for collection of the data. We thank the NEO study group, Petra Noordijk, Pat van Beelen and Ingeborg de Jonge for the coordination, lab and data management of the NEO study. The genotyping in the NEO study was supported by the Centre National de Génotypage (Paris, France), headed by Jean-Francois Deleuze. The NEO study is supported by the participating Departments, the Division and the Board of Directors of the Leiden University Medical Center, and by the Leiden University, Research Profile Area Vascular and Regenerative Medicine.

**NHS**
We would like to thank the participants and staff of the NHS and NHSII for their valuable contributions. The authors assume full responsibility for analyses and interpretation of these data. The NHS/NHSII studies thank the following state cancer registries for their help: AL, AZ, AR, CA, CO, CT, DE, FL, GA, ID, IL, IN, IA, KY, LA, ME, MD, MA, MI, NE, NH, NJ, NY, NC, ND, OH, OK, OR, PA, RI, SC, TN, TX, VA, WA, and WY. The NHS GWAS were supported by grants from the National Institutes of Health [NCI (CA40356, CA087969, CA055075, CA98233, U01 CA137088, R01 CA059045, R01 CA137178, R01 CA082838, R01 CA131332, R01 CA49449), NIDDK (DK058845, DK070756), NHGRI (HG004399, HG004728), NHLBI (HL35464), NIAMS (R01 AR056291)], UM1 CA186107, P01 CA87969.

**The Netherlands Twin Register**
We are grateful to all participants from the Netherlands Twin Register. We acknowledge funding the Netherlands Organization for Scientific Research (NWO) and The Netherlands Organization for Health Research and Development (ZonMW) grants, 904-61-193,480-04-004, 400-05-717, Addiction-31160008, 911-09-032, Biobanking and Biomolecular Resources Research Infrastructure (BBMRI –NL, 184.021.007); NWO 480-15-001/674: Netherlands Twin Registry Repository. European Research Council (ERC-230374 and ERC-284167); Rutgers University Cell and DNA Repository (NIMH U24 MH068457-06), the Avera Institute, Sioux Falls, South Dakota (USA) and the National Institutes of Health (NIH R01 HD042157-01A1). Part of the genotyping was funded by the Genetic Association Information Network (GAIN) of the Foundation for the National Institutes of Health and Grand Opportunity grants 1RC2 MH089951). We acknowledge support from VU Amsterdam and the Amsterdam Reproduction and Development (AR&D) research institute.

**NFBC1966**
We thank all cohort members and researchers who participated in the 31y and 46y NFBC1966 study. We also wish to acknowledge the work of the NFBC project centre. NFBC1966 received financial support from University of Oulu Grant no. 65354, Oulu University Hospital Grant no. 2/97, 8/97, Ministry of Health and Social Affairs Grant no. 23/251/97, 160/97, 190/97, National Institute for Health and Welfare, Helsinki Grant no. 54121, Regional Institute of Occupational Health, Oulu, Finland Grant no. 50621, 54231, University of Oulu Grant no. 24000692, Oulu University Hospital Grant no. 24301140, ERDF European Regional Development Fund Grant no. 539/2010 A31592, Academy of Finland grant numbers 24300796, 24302031, 285547 (EGEA) (MRJ); the Medical Research Council (MRC) UK (grant number G0601653) (MRJ); Medical Research Council Biotechnology and Biological Sciences Research Council PREcisE (Nutrition & Epigenome, The Joint Programming Initiative a Healthy Diet for a Healthy Life (JPI HDHL/EU-H2020)) (MRJ); Yrjö Jahnsson Foundation (SP), Päivikki and Sakari Sohlberg Foundation sr (SP); the European Union's Horizon 2020 programmes, iHealth-T2D (grant number 643774) and EDCMET (grant number 825762) (SP).

**The Orkney Complex Disease Study**
The Orkney Complex Disease Study (ORCADES) was supported by the Chief Scientist Office of the Scottish Government (CZB/4/276, CZB/4/710), a Royal Society URF to J.F.W., the MRC

Human Genetics Unit quinquennial programme “QTL in Health and Disease”, Arthritis Research UK and the European Union framework program 6 EUROSPAN project (contract no. LSHG-CT-2006-018947). DNA extractions were performed at the Edinburgh Clinical Research Facility, University of Edinburgh. We would like to acknowledge the invaluable contributions of the research nurses in Orkney, the administrative team in Edinburgh and the people of Orkney. For the purpose of open access, the author has applied a Creative Commons Attribution (CC BY) licence to any Author Accepted Manuscript version arising from this submission.

**QIMR**
We thank the participants and their families for contributing to this research. We also thank A Henders, B Usher, E Souzeau, A Kuot, A McMellon, MJ Wright, MJ Campbell, A Caracella, L Bowdler, S Smith, B Haddon, A Conciatore, D Smyth, H Beeby, O Zheng and B Chapman for their input into project management, databases, phenotype collection, and sample collection, processing and genotyping. The QIMR cohort was supported by National Institutes of Health (NIH) Grants AA07535, AA07728, AA13320, AA13321, AA14041, AA11998, AA17688, DA012854, DA019951, AA010249, AA013320, AA013321, AA011998, AA017688, and DA027995; by Grants from the Australian National Health and Medical Research Council (NHMRC) (241944, 339462, 389927, 389875, 389891, 389892, 389938, 442915, 442981, 496739, 552485, 552498 and 1075175); by Grants from the Australian Research Council (ARC) (A7960034, A79906588, A79801419, DP0770096, DP0212016, and DP0343921).

**SardiNIA**
This research was supported by the Intramural Research Program of the National Institute on Aging, National Institutes of Health (NIH) (contract 75N95021C00012-SardiNIA5).

**SHIP**
SHIP is part of the Community Medicine Research Network of the University Medicine Greifswald, which is supported by the German Federal State of Mecklenburg- West Pomerania.

**TwinGene**
TwinGene is part of the Swedish Twin Registry which is managed by Karolinska Institutet and receives funding through the Swedish Research Council under the grant no 2017-00641 and 2021-00180. Previous funding for data collection and build-up of TwinGene has been received through The Ministry for Higher Education; The Swedish Research Council (M-2005-1112); GenomEUtwin (EU/QLRT-2001-01254; QLG2-CT-2002-01254); NIH DK U01-066134; The Swedish Foundation for Strategic Research (SSF); Heart and Lung foundation no. 20070481.

**TWINS UK**
TwinsUK is funded by the Wellcome Trust, Medical Research Council, Versus Arthritis, European Union Horizon 2020, Chronic Disease Research Foundation (CDRF), Zoe Ltd, the National Institute for Health and Care Research (NIHR) Clinical Research Network (CRN) and Biomedical Research Centre based at Guy’s and St Thomas’ NHS Foundation Trust in partnership with King’s College London.

**VB**
We thank all the participants to the project, the San Raffaele Hospital MDs who contributed to clinical data collection, prof. Clara Camaschella who coordinated the data collection, Corrado Masciullo and Massimiliano Cocca for the database informatics. The research was supported by funds from Compagnia di San Paolo, Torino, Italy; Fondazione Cariplo, Italy; Telethon Italy; Ministry of Health, Ricerca Finalizzata 2008 and 2011-2012 and Public Health Genomics Project 2010.

**AGES**
This study has been funded by NIH contracts N01-AG-1-2100 and 271201200022C, the NIA Intramural Research Program, Hjartavernd (the Icelandic Heart Association), and the Althingi (the Icelandic Parliament). The study is approved by the Icelandic National Bioethics Committee, VSN: 00-063. The researchers are indebted to the participants for their willingness to participate in the study.

**FHS**
The Framingham Heart Study phenotype-genotype analyses were supported by NIA R01AG29451 JMM, KLL). The Framingham Heart Study of the National Heart Lung and Blood Institute of the National Institutes of Health and Boston University School of Medicine was supported by the National Heart, Lung and Blood Institute's Framingham Heart Study Contract No. N01-HC-25195 and its contract with Affymetrix, Inc for genotyping services (Contract No. N02-HL-6-4278). Analyses reflect intellectual input and resource development from the Framingham Heart Study investigators participating in the SNP Health Association Resource (SHARe) project. A portion of this research was conducted using the Linux Cluster for Genetic Analysis (LinGA-II) funded by the Robert Dawson Evans Endowment of the Department of Medicine at Boston University School of Medicine and Boston Medical Center.

**Rotterdam**
The generation and management of GWAS genotype data for the Rotterdam Study are supported by the Netherlands Organisation of Scientific Research NWO Investments (nr. 175.010.2005.011, 911- 03-012). This study is funded by the Research Institute for Diseases in the Elderly (014-93-015; RIDE2), the Netherlands Genomics Initiative (NGI)/Netherlands Organisation for Scientific Research (NWO) project nr. 050-060-810, and funding from the European Commision (HEALTH-F2-2008-201865, GEFOS; HEALTHF2-2008-35627, TREAT-OA) . The Rotterdam Study is funded by Erasmus Medical Center and Erasmus University, Rotterdam, Netherlands Organization for the Health Research and Development (ZonMw), the Research Institute for Diseases in the Elderly (RIDE), the Ministry of Education, Culture and Science, the Ministry for Health, Welfare and Sports, the European Commission (DG XII), and the Municipality of Rotterdam. We thank Pascal Arp, Mila Jhamai, Dr Michael Moorhouse, Marijn Verkerk, and Sander Bervoets for their help in creating the GWAS database. The authors are grateful to the study participants, the staff from the Rotterdam Study and the participating general practioners and pharmacists. We would like to thank Dr. Tobias A. Knoch, Anis Abuseiris, Karol Estrada, and Rob de Graaf as well as their institutions Biophysical Genomics, Erasmus MC Rotterdam, The Netherlands, and especially the national German MediGRID and Services@MediGRID part of the German D-Grid, both funded by the German Bundesministerium fuer Forschung und Technology under grants #01 AK 803 A-H and # 01 IG 07015 G for access to their grid resources.

**The Women's genome Health study**
The WGHS is supported by the National Heart, Lung, and Blood Institute (HL043851 and HL080467) and the National Cancer Institute (CA047988 and UM1CA182913), with funding for genotyping provided by Amgen.

**BMDCS**
We appreciate the dedication of the BMDCS study participants and their families, and the support of Dr. Karen Winer, Scientific Director of this effort. This study was funded by R01 HD58886, R01 HD100406, the Eunice Kennedy Shriver National Institute of Child Health and Human Development (NICHD) contracts (N01-HD-1-3228, -3329, -3330, -3331, -3332, -3333), and the CTSA program Grant 8 UL1 TR000077. S.F.A.G. is supported by the Daniel B. Burke Endowed Chair for Diabetes Research, R01 HD056465 and R01 HG010067.

**CHOP**
This research was financially supported by an Institute Development Award from the Children's Hospital of Philadelphia, a Research Development Award from the Cotswold Foundation, the Children´s Hospital of Philadelphia Endowed Chair in Genomic Research, the Daniel B. Burke Endowed Chair for Diabetes Research and NIH grant R01 HD056465. The authors thank the network of primary care clinicians and the patients and families for their contribution to this project and to clinical research facilitated by the Pediatric Research Consortium (PeRC) at The Children's Hospital of Philadelphia. R. Chiavacci, E. Dabaghyan, A. (Hope) Thomas, K. Harden, A. Hill, C. Johnson-Honesty, C. Drummond, S. Harrison, F. Salley, C. Gibbons, K. Lilliston, C. Kim, E. Frackelton, F. Mentch, G. Otieno, K. Thomas, C. Hou, K. Thomas and M.L. Garris provided expert assistance with genotyping and/or data collection and management. The authors would also like to thank S. Kristinsson, L.A. Hermannsson and A. Krisbjörnsson of Raförninn ehf for extensive software design and contributions.

**RAINE**
**Funding:** The Raine Study was supported by the National Health and Medical Research Council of Australia [grant numbers 572613, 403981 and 1059711] and the Canadian Institutes of Health Research [grant number MOP-82893].
**Acknowledgement:** The authors are grateful to the Raine Study participants and their families, and to the Raine Study research staff for cohort coordination and data collection. The authors gratefully acknowledge the NH&MRC for their long term funding to the study over the last 30 years and also the following institutes for providing funding for Core Management of the Raine Study: The University of Western Australia (UWA) , Curtin University, the Raine Medical Research Foundation, the UWA Faculty of Medicine, Dentistry and Health Sciences, the Telethon Kids Institute, the Women's and Infant's Research Foundation (King Edward Memorial Hospital), Murdoch University, The University of Notre Dame (Australia), and Edith Cowan University. The authors gratefully acknowledge the assistance of the Western Australian DNA Bank (National Health and Medical Research Council of Australia National Enabling Facility). We would also like to acknowledge the Raine Study participants for their ongoing participation in the study, and the Raine Study Team for study co-ordination and data collection. This work was supported by resources provided by the Pawsey Supercomputing Centre with funding from the Australian Government and Government of Western Australia.

**deCODE**
We thank the study subjects for their valuable participation. All deCODE collaborators in this study are employees of deCODE Genetics/Amgen, Inc.

**VIKING**
The Viking Health Study – Shetland (VIKING) was supported by the MRC Human Genetics Unit quinquennial programme grant "QTL in Health and Disease". DNA extractions and genotyping were performed at the Edinburgh Clinical Research Facility, University of Edinburgh. We would like to acknowledge the invaluable contributions of the research nurses in Shetland, the administrative team in Edinburgh and the people of Shetland. For the purpose of open access, the author has applied a Creative Commons Attribution (CC BY) licence to any Author Accepted Manuscript version arising from this submission.

**TRAILS**
TRAILS (TRacking Adolescents' Individual Lives Survey): TRAILS is a collaborative project involving various departments of the University Medical Center and University of Groningen, the University of Utrecht, the Radboud University Medical Center, and the Parnassia Psychiatric Institute, all in the Netherlands. TRAILS has been financially supported by grants from the Netherlands Organization for Scientific Research NWO (Medical Research Council program grant GB-MW 940-38-011; ZonMW Brainpower grant 100-001-004; ZonMw Risk

Behavior and Dependence grant 60-60600-97-118; ZonMw Culture and Health grant 261-98-710; Social Sciences Council medium-sized investment grants GB-MaGW 480-01-006 and GB-MaGW 480-07-001; Social Sciences Council project grants GB-MaGW 452-04-314 and GB-MaGW 452-06-004; ZonMw Longitudinal Cohort Research on Early Detection and Treatment in Mental Health Care grant 636340002, NWO large-sized investment grant 175.010.2003.005; NWO Longitudinal Survey and Panel Funding 481-08-013 and 481-11-001; NWO Vici 016.130.002, 453-16-007/2735, and Vi.C 191.021; NWO Gravitation 024.001.003), the Dutch Ministry of Justice (WODC), the European Science Foundation (EuroSTRESS project FP-006), the European Research Council (ERC-2017-STG-757364 en ERC-CoG-2015-681466), Biobanking and Biomolecular Resources Research Infrastructure BBMRI-NL (CP 32), The Gratema foundation, the Jan Dekker foundation, the participating universities, and Accare Centre for Child and Adolescent Psychiatry. Statistical analyses were carried out on the Genetic Cluster Computer (http://www.geneticcluster.org), which is financially supported by the Netherlands Scientific Organization (NWO 480-05-003) along with a supplement from the Dutch Brain Foundation. We are grateful to all adolescents who participated in this research and to everyone who worked on this project and made it possible.

**The Breast Cancer Association Consortium**
The BCAC data management and breast cancer genome-wide association analyses were supported by Cancer Research UK (C1287/A10118, C1287/A16563, C1287/A10710), the PERSPECTIVE project via the Government of Canada through Genome Canada and the Canadian Institutes of Health Research (grant GPH-129344), the 'Ministère de l'Économie, de la Science et de l'Innovation du Québec' through Genome Québec and grant PSR-SIIRI701, the Quebec Breast Cancer Foundation, the National Institutes of Health (U19 CA148065, CA128978, X01HG007492) and Post-Cancer GWAS initiative (1U19 CA148537, 1U19 CA148065 and 1U19 CA148112 - the GAME-ON initiative), the Department of Defence (W81XWH-10-1-0341), the Canadian Institutes of Health Research (CIHR) for the CIHR Team in Familial Risks of Breast Cancer, Komen Foundation for the Cure, the Breast Cancer Research Foundation, the European Community´s Seventh Framework Programme (HEALTH-F2-2009-223175), and European Union's Horizon 2020 Research and Innovation Programme (grants 634935 and 633784 for BRIDGES and B-CAST respectively). All studies and funders are listed Michailidou et al (2017) [PMID 29059683].

**OCAC**
The Ovarian Cancer Association Consortium is supported by a grant from the Ovarian Cancer Research Fund thanks to donations by the family and friends of Kathryn Sladek Smith (PPD/RPCI.07). The scientific development and funding for this project were in part supported by the US National Cancer Institute GAME-ON Post-GWAS Initiative (U19-CA148112). This study made use of data generated by the Wellcome Trust Case Control consortium that was funded by the Wellcome Trust under award 076113. The results published here are in part based upon data generated by The Cancer Genome Atlas Pilot Project established by the National Cancer Institute and National Human Genome Research Institute (dbGap accession number phs000178.v8.p7).

The OCAC OncoArray genotyping project was funded through grants from the U.S. National Institutes of Health (CA1X01HG007491-01 (C.I.A.), U19-CA148112 (T.A.S.), R01-CA149429 (C.M.P.) and R01-CA058598 (M.T.G.); Canadian Institutes of Health Research (MOP-86727 (L.E.K.) and the Ovarian Cancer Research Fund (A.B.). The COGS project was funded through a European Commission's Seventh Framework Programme grant (agreement number 223175 - HEALTH-F2-2009-223175).

**23andMe**
We thank the research participants and employees of 23andMe for making this work possible.

**The Biobank Japan**
We would express our sincere gratitude to the study participants, the research and medical staffs in the BBJ. This study was supported by the BioBank Japan project, which was supported by the Ministry of Education, Culture, Sports, Sciences and Technology of the Japanese Government and AMED under grant numbers 17km0305002 and 18km0605001. This study was supported by JSPS KAKENHI (JP20H00462) from the Ministry of Education, Culture, Sports, Science and Technology of Japan and Japan Agency for Medical Research and Development (AMED) (JP21kk0305013, JP21tm0424220 and JP21ck0106642).

**China Kadoorie Biobank**
CKB gratefully acknowledges the participants in the study and the members of the survey teams in each of the 10 regional centres, and to the project development and management teams based at Beijing, Oxford and the 10 regional centres. China's National Health Insurance provides electronic linkage to all hospital treatments. The CKB baseline survey and the first re-survey were supported by the Kadoorie Charitable Foundation in Hong Kong. Long-term follow-up was supported by the Wellcome Trust (212946/Z/18/Z, 202922/Z/16/Z, 104085/Z/14/Z, 088158/Z/09/Z), the National Key Research and Development Program of China (2016YFC0900500, 2016YFC0900501, 2016YFC0900504, 2016YFC1303904), and the National Natural Science Foundation of China (81941018, 82192900, 91843302, 91846303). DNA extraction and genotyping was funded by GlaxoSmithKline, and the UK Medical Research Council (MC-PC-13049, MC-PC-14135). The project is supported by core funding from the UK Medical Research Council (MC_UU_00017/1, MC_UU_12026/2, MC_U137686851), Cancer Research UK (C16077/A29186; C500/A16896), and the British Heart Foundation (CH/1996001/9454) to the Clinical Trial Service Unit and Epidemiological Studies Unit and to the MRC Population Health Research Unit at Oxford University. Computation used the Oxford Biomedical Research Computing (BMRC) facility, a joint development between the Wellcome Centre for Human Genetics and the Big Data Institute supported by Health Data Research UK and the NIHR Oxford Biomedical Research Centre.

**Danish Blood Donor Study**
**Funding:** The initiation of the Danish Blood Donor Study was supported by the Danish Administrative Regions (02/2611) and the Danish Council for Independent Research (09–069412). The study is currently funded by the Danish Administrative Regions and Bio- and Genome Bank Denmark.
**Acknowledgements:** We thank the Danish blood donors for their valuable participation in the Danish Blood Donor Study as well as the staff at the blood centers making this study possible.

**MoBa**
The Norwegian Mother, Father and Child Cohort Study (MoBa) is a population-based pregnancy cohort study conducted by the Norwegian Institute of Public Health. Participants were recruited from all over Norway from 1999-2008. The women consented to participation in 41% of the pregnancies. The cohort includes approximately 114.500 children, 95.200 mothers and 75.200 fathers. The current study is based on version 10 of the quality-assured data files released for research in Novel Tools for Early Childhood Predisposition to Obesity. The establishment of MoBa and initial data collection was based on a license from the Norwegian Data Protection Agency and approval from The Regional Committees for Medical and Health Research Ethics. The MoBa cohort is currently regulated by the Norwegian Health Registry Act. The current study was approved by The Regional Committees for Medical and Health Research Ethics (no. 2012/67). The Medical Birth Registry (MBRN) is a national health registry containing information about all births in Norway.

We thank the Norwegian Institute of Public Health (NIPH) for generating high-quality genomic data. This research is part of the HARVEST collaboration, supported by the Research Council of Norway (#229624). We also thank the NORMENT Centre for providing genotype data, funded by the Research Council of Norway (#223273), South East Norway Health Authorities and Stiftelsen Kristian Gerhard Jebsen. We further thank the Center for Diabetes Research, the University of Bergen for providing genotype data and performing quality control and imputation of the data funded by the ERC AdG project SELECTionPREDISPOSED, Stiftelsen Kristian Gerhard Jebsen, Trond Mohn Foundation, the Research Council of Norway, the Novo Nordisk Foundation, the University of Bergen, and the Western Norway Health Authorities.

Supported by grants (to P.R.N.) from the European Research Council (AdG #293574), the Bergen Research Foundation ("Utilizing the Mother and Child Cohort and the Medical Birth Registry for Better Health"), Stiftelsen Kristian Gerhard Jebsen (Translational Medical Center), the University of Bergen, the Research Council of Norway (FRIPRO grant #240413), the Western Norway Regional Health Authority (Strategic Fund "Personalized Medicine for Children and Adults"), the Novo Nordisk Foundation (grant #54741), and the Norwegian Diabetes Association; and (to S.J.) Helse Vest's Open Research Grant (grant #912250), the Research Council of Norway (FRIPRO grant #315599), and Novo Nordisk Foundation (grant #NNF21OC0070349 . This work was partly supported by the Research Council of Norway through its Centres of Excellence funding scheme (#262700), Better Health by Harvesting Biobanks (#229624) and The Swedish Research Council, Stockholm, Sweden (2015-02559), The Research Council of Norway, Oslo, Norway (FRIMEDBIO #547711, March of Dimes (#21-FY16-121). The Norwegian Mother, Father, and Child Cohort Study is supported by the Norwegian Ministry of Health and Care Services and the Ministry of Education and Research, NIH/NIEHS (contract no N01-ES-75558), NIH/NINDS (grant no.1 UO1 NS 047537-01 and grant no.2 UO1 NS 047537-06A1).

Data from the Norwegian Mother, Father and Child Cohort Study and the Medical Birth Registry of Norway used in this study are managed by the national health register holders in Norway (Norwegian Institute of public health) and can be made available to researchers, provided approval from the Regional Committees for Medical and Health Research Ethics (REC), compliance with the EU General Data Protection Regulation (GDPR) and approval from the data owners. The consent given by the participants does not open for storage of data on an individual level in repositories or journals. Researchers who want access to data sets for replication should apply through helsedata.no. Access to data sets requires approval from The Regional Committee for Medical and Health Research Ethics in Norway and an agreement with MoBa.

Analyses were performed using digital laboratories in HUNT Cloud at the Norwegian University of Science and Technology, Trondheim, Norway. We are grateful for outstanding support from the HUNT Cloud community. The participating families who contributed with data and biological material are gratefully acknowledged.

#### Additional acknowledgements

This research has been conducted using the UK Biobank Resource under Application Number 9905. We thank the KoGES study for providing data.

#### Conflicts of interest

JRBP and EJG are employed by Adrestia Therapeutics. DJT is employed by Genomics PLC. DLC and JPB are employed by GSK. ET is employed by Pfizer. DAL has received support from Roche Disgnostics and Medtroic Ltd for work unrelated to the research in this paper. TDS is co-founder and stakeholder of Zoe Global Ltd. PAF conducts research funded by Amgen, Novartis and Pfizer, he received Honoraria from Roche, Novartis and Pfizer. MWB conducts research funded by Amgen, Novartis and Pfizer.

#### Additional funding information

ASB is funded by the Deutsche Forschungsgemeinschaft (DFG, German Research Foundation) – 464240267. NF Is funded by NIH DK117445, MD012765, HL163972. RM was supported by the Estonian Research Council grant PRG1911. SEM is supported by an Australian National Health and Medical Research Council Investigator Grant (APP1172917). CT is funded by JSPS KAKENHI (JP20H00462) from the Ministry of Education, Culture, Sports, Science and Technology of Japan and Japan Agency for Medical Research and Development (AMED) (JP21kk0305013, JP21tm0424220 and JP21ck0106642). VV and CH are funded by MRC University Unit Programme Grant “QTL in Health and Disease” (U. MC_UU_00007/10). GWM is supported by an Australian National Health and Medical Research Council Investigator Grant (APP1177194). DIB is funded by Royal Netherlands Academy of Arts and Sciences (KNAW PAH/6635). TE was supported by the Estonian Research Council grant PRG1291. DAL is supported by the British Heart Foundation (AA/18/1/34219 and CH/F/20/90003), the European Research Council under the European Union’s Horizon 2020 research and innovation program (grant agreements No 101021566) and UK Medical Research Council (MC_UU_00011/6). NGM is supported by an Australian National Health and Medical Research Council Investigator Grant (APP1172990). NJT is a Wellcome Trust Investigator (202802/Z/16/Z), is the PI of the Avon Longitudinal Study of Parents and Children (MRC & WT 217065/Z/19/Z), is supported by the University of Bristol NIHR Biomedical Research Centre (BRC-1215-2001), the MRC Integrative Epidemiology Unit (MC_UU_00011/1) and works within the CRUK Integrative Cancer Epidemiology Programme (C18281/A29019). SFAG is supported by the Daniel B. Burke Endowed Chair for Diabetes Research, R01 HD056465 and R01 HG010067. MV is supported by the Research Council of Norway (project #301178) and the University of Bergen.
